# Psychotherapies for obsessive-compulsive disorder have distinct effects on brain activity during emotional processing

**DOI:** 10.64898/2026.02.10.26345974

**Authors:** Chris Vriend, Aniek Broekhuizen, Nadja Wolf, Patricia van Oppen, Odile A. van den Heuvel, Henny A. D. Visser

**Author notes:** = shared last author (Odile A. van den Heuvel, Henny A. D. Visser). **Corresponding author**: Dr. Chris Vriend, Department of Anatomy & Neurosciences, O|2 Building 13W09. Amsterdam UMC location VUmc. PO Box 7057, 1007 MB, Amsterdam, The Netherlands. | E.

## Abstract

**Background:** To clarify the working mechanisms of psychotherapy for obsessive-compulsive disorder (OCD), we studied the neural effects of two psychotherapies: cognitive behavioral therapy with exposure and response prevention (CBT-ERP) and inference-based cognitive behavioral therapy (I-CBT).

**Methods:** Fifty-five individuals with OCD completed an emotional processing task during fMRI before and after 20 weekly psychotherapy sessions, using general fear and OCD-related visual stimuli. Forty-two healthy controls performed the task once. We used Bayesian region-of-interest analyses to assess changes in brain activation in prefrontal, limbic, sensory, subcortical, and visual areas, and their association with symptom improvement.

**Results:** After treatment, the CBT-ERP group (N=28) showed strong credible evidence for decreased activation across all brain regions during fear (but not OCD) versus neutral stimuli, especially in treatment responders. Conversely, the I-CBT group (N=27) showed increased activation during fear versus neutral stimuli in the precentral gyrus and lateral occipital cortex (LOC), which correlated with symptom improvement. A similar but weaker pattern was observed for OCD-related stimuli. Across all ROIs, baseline fear-related activity was associated with symptom improvement in CBT-ERP, while lower baseline activity was associated with improvement in I-CBT in, amongst others, the precentral gyrus and dorsolateral prefrontal cortex. Lower baseline LOC activation during OCD-related stimuli was linked to symptom improvement after both psychotherapies.

**Conclusions:** The results support CBT-ERP’s mechanism of fear reduction and I-CBT’s mechanism of sensory engagement. Visual brain activity during emotional processing may predict treatment response across psychotherapies.

## Introduction

The gold standard treatments for obsessive-compulsive disorder (OCD) include cognitive behavioral therapy with exposure and response prevention (CBT-ERP) and pharmacological treatment with serotonin reuptake inhibitors (SRIs) that are administered alone or in combination. CBT-ERP aims to help people with OCD to confront anxiety-provoking situations without performing compulsions, breaking the cycle of avoidance and negative thinking (McKay et al., 2015). Although CBT-ERP is an effective treatment, up to 50% of individuals do not show sufficient benefit (Atmaca, 2016; Ost et al., 2015; Pallanti and Quercioli, 2006; Skapinakis et al., 2016). This has spurred the development of new forms of psychotherapy, such as inference based cognitive behavioral therapy (I-CBT) (Aardema et al., 2022). The premise of I-CBT is that individuals with OCD experience obsessional doubt because they confuse imaginable with actual scenarios and show an overreliance on the imagination (Aardema et al., 2022). I-CBT targets the underlying dysfunctional reasoning and mistrust of the senses and, crucially, does not include an exposure component (Wolf et al., 2024).

Using a randomized controlled non-inferiority trial in N=197 individuals with OCD, the arrIBA study (Alternative treatment to Reduce chronicity in OCD: Research Into Brain response and Adequacy of treatment; ClinicalTrials.gov NCT03929081) showed that both CBT-ERP and I-CBT significantly reduced OCD symptom severity, improved insight, reduced comorbid depression and anxiety up to one year after treatment and there were no significant differences between the groups in clinical improvement, response, remission or drop-out rate (Wolf et al., 2024). These findings coincide with other previous studies on CBT-ERP and I-CBT (Aardema et al., 2022; O’Connor et al., 2005; Visser et al., 2015).

Emotional processing tasks during functional MRI are frequently used in OCD to understand the disorder and treatments effects. Compared to healthy controls, previous meta-analyses showed hyperactivity in frontolimbic areas such as the amygdala, OFC, subgenual anterior cingulate cortex (sgACC), insula and ventromedial cortex (vmPFC) in individuals with OCD (Thorsen et al., 2018) and hypoactivity in occipital areas (Yu et al., 2022). A recent mega-analysis conducted by the ENIGMA-OCD workgroup in N=1086 individuals also showed credible evidence for higher activity in the OFC, sgACC and vmPFC and lower activation of the occipital cortex during emotional processing, particularly the lateral occipital cortex (LOC) (Dzinalija et al., 2024). Studies investigating the effects of CBT-ERP on brain activation during emotional processing (Baioui et al., 2013; Morgieve et al., 2014; Nakao et al., 2005; Schiepek et al., 2013; van der Straten et al., 2024) generally showed decreased activation of frontolimbic brain areas, such as the orbitofrontal cortex (OFC), anterior cingulate cortex (ACC), insula, striatum and thalamus after treatment (see (Poli et al., 2022; Thorsen et al., 2015) for systematic reviews).

Studies have also investigated associations between baseline task fMRI during emotional processing and treatment outcome. One study found no association between pre-treatment activity and treatment outcome (Morgieve et al., 2014), another found that pre-treatment task-related activation of the amygdala was associated with treatment outcome after CBT-ERP (Olatunji et al., 2014) or with the outcome of CBT-ERP in combination with repetitive transcranial magnetic stimulation (Houben et al., 2025). According to a meta-analysis across multiple anxiety disorders, successful CBT was associated with higher pre-treatment task-related activity in the inferior frontal gyrus (IFG), anterior insula, dorsomedial prefrontal cortex (dmPFC) and dACC during emotional tasks (Picó-Pérez et al., 2022). In sum, while multiple studies have shown evidence for treatment-induced changes in brain activity related to emotional processing after CBT-ERP, and that pre-treatment activation patterns are associated with treatment outcome, no study has yet investigated the neural effects of I-CBT.

Using data from the arrIBA trial, we investigated the effects of CBT-ERP and I-CBT on emotional processing-related brain activation. As preregistered (https://osf.io/27wrh), we hypothesized that regardless of condition, successful treatment would be associated with decreased task-related activity during emotional processing in the OFC, dorsal ACC, thalamus, caudate nucleus, and limbic regions. Relative to I-CBT, we expected the CBT-ERP group to show a higher increase in activity in cognitive control areas (targeted in CBT-ERP), while the I-CBT group would show a higher increase in activation of areas involved in sensory processing (targeted in I-CBT) (Aardema et al., 2022). We additionally associated baseline task-related activity to treatment response to identify treatment predictors, expecting that, across conditions, higher pre-treatment activation in limbic areas and lower activation in cognitive control areas would be associated with symptom improvement. We also expected lower pre-treatment activation in cognitive control areas to be associated with clinical improvement in CBT-ERP, and lower pre-treatment activation in sensory processing areas with clinical improvement in I-CBT.

## Methods

### Participants

A subset (n=86) of participants from the arrIBA trial underwent MRI before and after treatment. Details on trial design, recruitment and complete in– and exclusion criteria are described elsewhere (Wolf et al., 2024). Briefly, participants with DSM-5 established OCD were eligible if they had a Yale-Brown Obsessive-Compulsive Scale (YBOCS; (Goodman et al., 1989)) score ≥16, were not on medication or on a stable dose for at least three months, did not receive cognitive behavioral therapy in the last six months and met MRI safety criteria. MRI sessions took place within three weeks before and after treatment. An additional n=42 healthy controls underwent one MRI session and did not have any mental illness or first-degree relatives with OCD or tic disorders, used no medication, and met MRI safety criteria. Study procedures were in accordance with the declaration of Helsinki and written informed consent was obtained from all participants. The study was approved by the ethics committee of VU Medical Center.

### Treatment and clinical assessment

Full treatment protocol and main clinical results on the full sample of N=197 are described elsewhere (Wolf et al., 2024). Briefly, patients were randomized over two forms of psychotherapy: CBT-ERP and I-CBT. Both treatments were protocolized into 20 weekly sessions of 45 minutes, adapted from a previous randomized-controlled trial (Visser et al., 2015). Treatment was delivered by trained psychotherapists and bi-monthly supervision meetings were provided. Participants were included in the per-protocol analyses if they attended at least 12 sessions, or if treatment ended earlier due to all treatment goals being achieved. Directly before and after treatment, participants were assessed by blinded research assistants on: OCD symptom severity with the YBOCS (Goodman et al., 1989); comorbidity with the Structured Clinical Interview for DSM-5 (First et al., 2016); insight with the Overvalued Ideas Scale (OVIS) (Neziroglu et al., 1999), and severity of depressive symptoms with the Beck Depression Inventory (BDI) (Beck et al., 1961). We calculated the change in absolute YBOCS scores and classified individuals as responders or non-responders based on the clinically established cut-off of 35% improvement (Mataix-Cols et al., 2016).

### Imaging acquisition and task paradigm

MR images were obtained before and after treatment on a GE discovery MR750 3T with a 32-channel head coil (General Electric, Milwaukee, U.S.). We acquired T1-weighted structural MRI and gradient echo-planar imaging for the task functional MRI with fieldmaps to correct for geometric distortions. Scan parameters are provided in the supplements. The complete task design has been described previously (Broekhuizen et al., 2023). An emotional processing task was performed during fMRI in which neutral, general fearful and OCD-related pictures were presented in blocks of 20 seconds (nine blocks per condition; five images per block). Pictures were previously validated and originated from the International Affective Picture Set (Lang et al., 1999), popular media and previous databases (de Wit et al., 2015; Mataix-Cols et al., 2009; van den Heuvel et al., 2004). OCD-related stimuli consisted of washing, checking and symmetry subtypes, while neutral stimuli were scrambled versions of the OCD-related and generally fearful images. At each timepoint, participants performed one of two (counterbalanced) versions of the task. Before and after the task, participants rated their distress level on a scale of 1-100. After each scan session, participants also rated the unpleasantness of a subset of the visual stimuli on a 5-point Likert scale.

### MRI preprocessing and task contrast

Task fMRI data were preprocessed using fMRIprep (v21.0.1), including corrections for susceptibility-induced distortions, realignment, slice timing and warping to Montreal Neurological Institute (MNI) space (boilerplate in the supplements). Images were visually assessed for artifacts and registration errors. Functional scans with a framewise displacement > 0.5mm were excluded. We used Statistical Parametric Mapping 12 (SPM) to smooth the normalized images with an 8mm Gaussian kernel and perform first and second level analyses. We used a block design to model the OCD-related, general fearful and scrambled pictures for each participant (first level) and to define a fear, OCD and emotional (OCD+fearful pictures) > scrambled contrast. While we pre-registered the emotional contrast as the primary contrast, we observed that the fear and OCD contrast showed opposite directions of effect and we therefore decided to report and interpret the results of the fear and OCD contrasts in the main text. For transparency we still report the emotional contrast in the supplements. We used the default (128s) high-pass filter and added motion parameters (Friston-24) as covariates of no interest. Participant-specific activation contrast estimates were extracted using MarsBar (v0.44, Brett et al., 2002) for region-of-interest analyses. We used different ROIs for pre-post and prediction analysis, based on the relevant meta-analyses (Picó-Pérez et al., 2022; Thorsen et al., 2018; Yu et al., 2022). Cortical ROIs were defined as 5mm spheres around the MNI coordinates reported in the meta-analyses (see supplementary Table S1). Subcortical ROIs were based on the automated anatomical labelling (AAL) atlas.

### Statistical analysis

Analyses were preregistered at https://osf.io/27wrh. Demographic and clinical characteristics were analyzed using R (4.1.3) and group comparisons were performed using t-tests or χ^2^ tests where appropriate. We performed mixed model analyses with a treatment condition*time interaction term and a random intercept for participant to test for differences between the treatments on YBOCS, OVIS and BDI scores.

For pre-to-post imaging analysis, we modelled the difference in task-related activation between T0 (pre-treatment) and T1 (post-treatment) at first level. The extracted ROI-specific contrast estimates were (1) analyzed for each treatment group (CBT-ERP and I-CBT), (2) compared between the treatment groups, (3) associated with change in YBOCS and (4) compared between responders and non-responders. For prediction analysis, we related pre-treatment task activation with the pre-to-post treatment change in YBOCS scores and compared baseline activity between responders and non-responders per treatment group. Age and sex were included as covariates in all analyses alongside pre-treatment YBOCS in prediction analyses and the pre-post change in YBOCS associations. Medication status was added as additional covariate in sensitivity analyses. We additionally compared baseline activity between the two treatment groups and healthy controls.

We used a Bayesian multilevel modelling approach implemented in AFNI: Region-Based Analysis Program through Bayesian Multilevel Modeling (v1.0.10) (Chen et al., 2019). The main advantage of this approach is that multiple ROIs can be incorporated into one model, thereby incorporating shared information between ROIs and eliminating the need for multiple comparison correction (Chen et al., 2019). For interpretation of the results, the full posterior distributions should be considered. To guide the reader, results are summarized in the text using the positive posterior probability (P+) that provides a point estimate of the credibility of evidence for an effect. We classify effect credibility as *moderate* (P+ between [1-]0.05 and [1-]0.10), *strong* (P+ between [1-]0.01 and [1-]0.05), and *very strong* (P+ <0.01 or >0.99) in the main text. We used noninformative Gaussian priors or students’ t distribution in case of small subgroups (N<20) with 1,000 iterations and 6 chains.

Lastly, we performed exploratory whole-brain analyses in SPM using repeated measures ANOVA for the pre-post analyses and multiple regression for the prediction analyses with an extent thresholds of k=5 and *p*<0.001, uncorrected.

## Results

### Demographic and clinical characteristics

Out of 86 participants who underwent MRI at T0, four were excluded (two for not meeting inclusion criteria, two for incomplete task data). Post-treatment YBOCS scores were available for 73 patients (intention-to-treat [ITT] sample), with 65 completing treatment (per-protocol [PP] sample); These samples had similar demographic and clinical characteristics (supplementary table 2). Dropouts (N=9) were similar to the ITT sample except for higher pre-task distress. No differences were found between CBT-ERP and I-CBT groups in either ITT (Table 1) or PP samples (supplementary table 3). Fifty-five participants (27 CBT-ERP, 28 I-CBT) completed post-treatment MRI; baseline characteristics were comparable between groups (Table 1; see supplementary table 4 for a comparison with the drop-out sample). The CBT-ERP group showed a greater reduction in YBOCS than I-CBT (B[SE]=4.43[1.7], P=0.01), but groups did not differ in responder rates, BDI, or OVIS scores (supplementary table 5). Analyses on distress and picture ratings are reported in supplementary table 6 and supplementary figures 2-3.

**Table 1.**
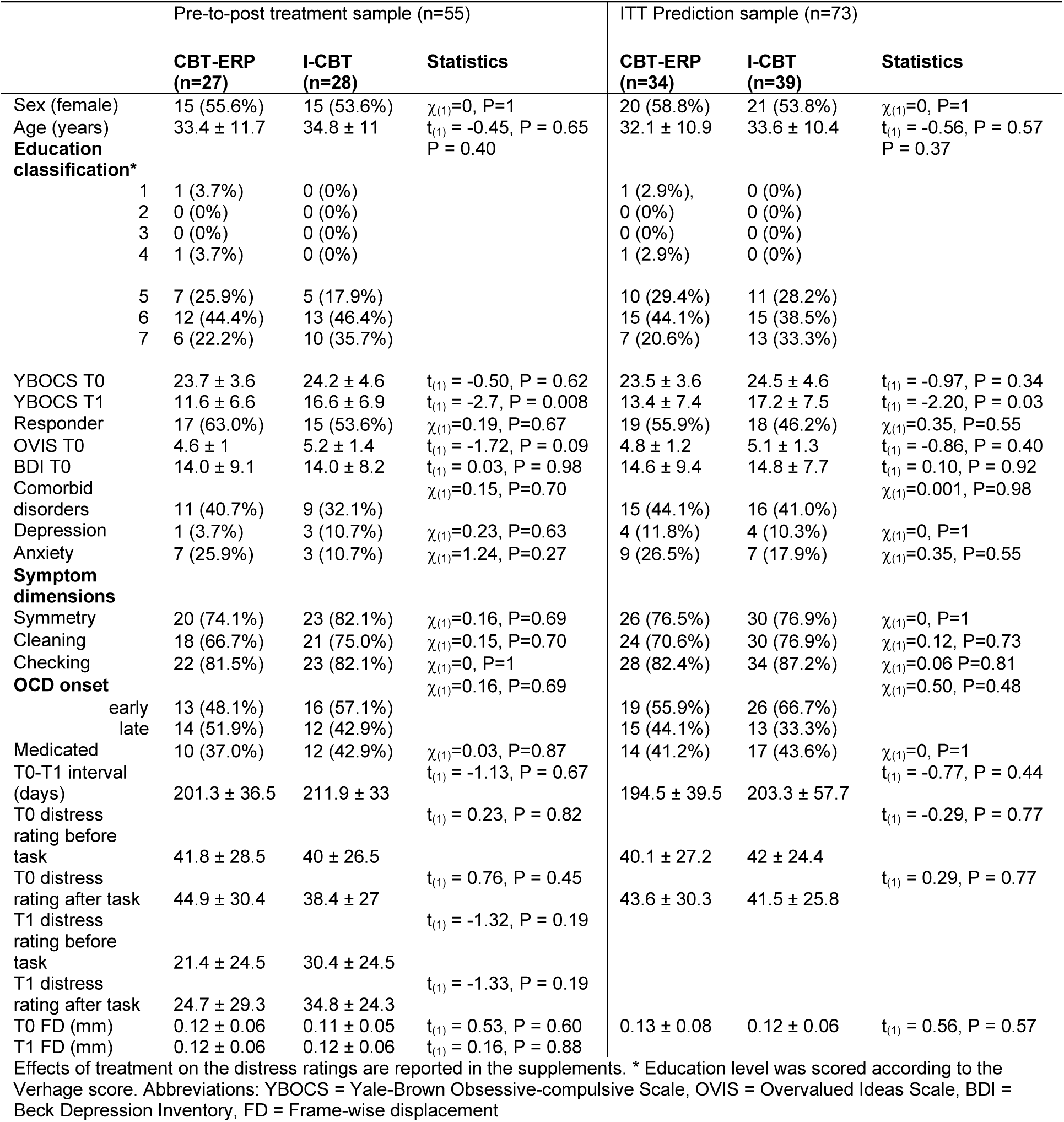
– Demographic and clinical characteristics of the pre-post treatment and ITT sample.

### Pre-treatment activation differences

Region-of-interest analyses showed strong credible evidence for higher baseline task-related activation in the CBT-ERP than the I-CBT group in most brain areas across fear, OCD, and emotion contrasts (supplementary figures 4-6). Compared to healthy controls (N=42), the CBT-ERP group showed no baseline differences except for higher activation in the fear contrast in regions like the amygdala, LOC, and middle prefrontal cortex (mPFC). The I-CBT group generally showed evidence for lower activation than healthy controls across all regions and contrasts, except for the LOC. These findings remained after adjusting for medication status.

### Pre-to-post treatment ROI analyses

#### Fear contrast

There was evidence for opposite pre-to-post treatment changes in activation between groups (Figure 1a). The CBT-ERP group showed moderate to strong evidence for decreased task-related activation after treatment across almost all ROIs (P+ = 0.00-0.10), while the I-CBT group showed moderate evidence for increased activation in the left (P+ = 0.94) and right (P+ = 0.93) precentral gyrus, right LOC (P+ = 0.94), and right middle temporal/angular gyrus (P+ = 0.91). Direct comparison showed credible evidence for larger decreases in activation in the CBT-ERP group across all ROIs (P+ = 0.00-0.06). Adjusting for medication only affected the decrease in LOC activity in the CBT-ERP group.

**Figure 1.**
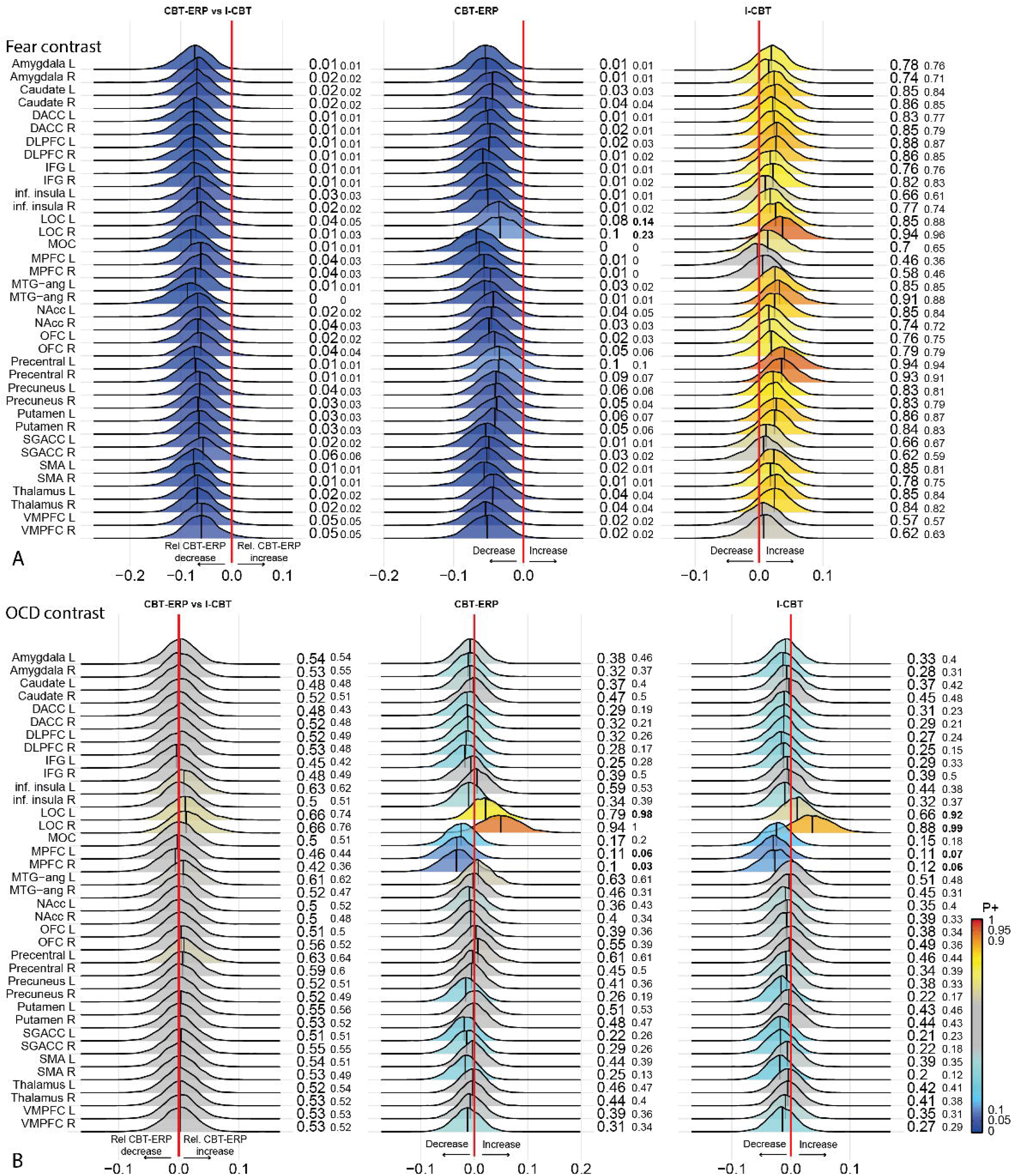
– Bayesian posterior distribution plots of the pre-to-post treatment differences in task activation during the Fear (a) and OCD (b) contrast. The first column shows the relative differences the CBT-ERP group and I-CBT show in the changes in task related activation after treatment. Column two and three show the changes from pre-to-post treatment in the CBT-ERP and I-CBT group, respectively. The posterior distribution communicates the credibility of an effect. Positive posterior probabilities (*P+*) are shown next to each distribution and color coded. *P+* values ≥0.90 indicate moderate to very high credibility for a positive effect, P+ ≤0.10 indicate moderate to very high credibility for a negative effect. The smaller P+ values represent the posterior probabilities when adjusting for medication status. The meaning of the direction of effects is shown next to the red zero-effect line. See supplementary Table 1 for the definition of the abbreviated regions of interest. All analyses were adjusted for age and sex.

The CBT-ERP group showed credible associations between symptom reduction and decreased brain activation across all ROIs, with the strongest association in bilateral LOC (P+ < 0.1; Figure 2a). In contrast, the I-CBT group showed higher brain activation with symptom improvement, especially in the left (P+ = 0.97) and right precentral gyrus (P+ = 1.0), right mPFC (P+ = 0.97), right DLPFC (P+ = 0.91), right precuneus (P+ = 0.91), and middle occipital cortex (P+ = 0.91). Adjusting for medication status had little effect on these associations.

**Figure 2.**
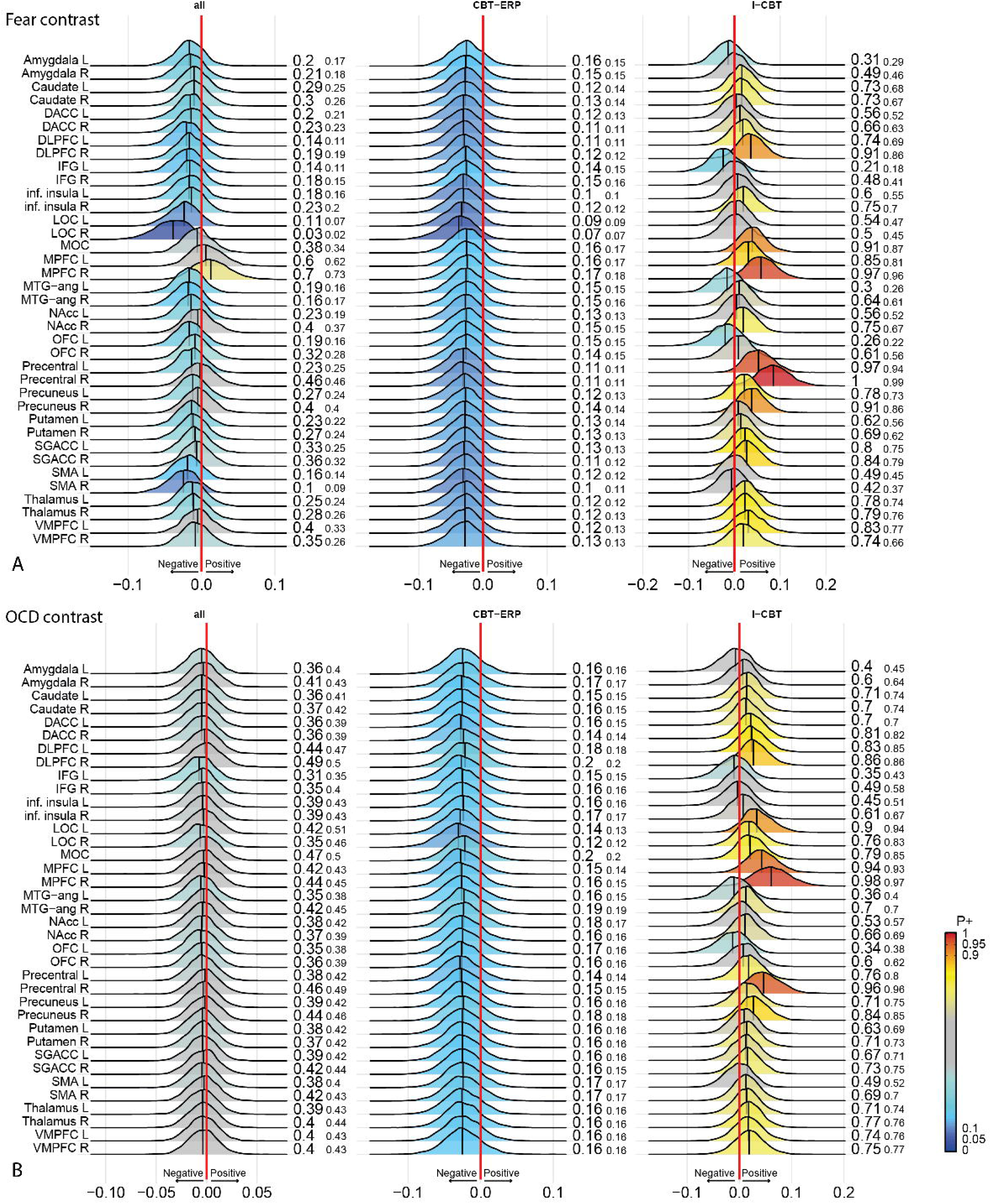
– Bayesian posterior distribution plots of the association between pre-to-post treatment differences in task activation during the fear (a) and OCD (b) contrast and change in symptoms. The first column shows the association across both treatment groups. Column two and three show the associations in the CBT-ERP and I-CBT group, respectively. See legend of Figure 1 for an explanation of the color codes. The smaller P+ values represent the posterior probabilities when adjusting for medication status. The meaning of the direction of effects are shown next to the red zero-effect line. See supplementary Table 1 for the definition of the abbreviated regions of interest. All analyses were adjusted for age and sex and baseline YBOCS score.

CBT-ERP responders showed evidence for decreased activation across all ROIs, while non-responders did not (supplementary figure 7). I-CBT responders showed increased activity after treatment in the bilateral precentral gyrus (P+ = 0.95-0.96), bilateral DLPFC (P+ = 0.92-0.93), and right middle temporal/angular gyrus (P+ = 0.93; supplementary figure 8). For both groups, there was low credibility for larger changes in responders compared to non-responders. Adjusting for medication status had little effect on these associations.

#### OCD contrast

Both groups showed evidence for increased activation after treatment in the bilateral LOC, particularly after adjusting for medication status (P+ > 0.92; Figure 1b). There was also evidence for decreased activation in the bilateral mPFC, but only after adjusting for medication status (P+ range 0.03-0.07). There was no credible evidence for a between-group difference in the change in activation. There was also little evidence for an association between the change in symptoms and change in activation in the CBT-ERP group before and after adjusting for medication status (Figure 2b). Conversely, the I-CBT group showed – whether or not adjusting for medication status – credible evidence for positive associations between change in symptoms and change in activity in the bilateral mPFC (P+ = 0.94-0.98), right precentral gyrus ((P+ = 0.96) and left LOC (P+ = 0.9).

CBT-ERP responders showed moderate evidence for a decrease in right mPFC activity (P+ = 0.09), but no credible evidence was found for a difference in change in activation between responders and non-responders to CBT-ERP with or without adjusting for medication status (supplementary figure 9). Non-responders to I-CBT showed credible evidence for a decrease in right mPFC activity, particularly when adjusting for medication status (P+ = 0.04) (supplementary figure 10). Results on the emotional contrast are reported in the supplements (supplementary figures 11-14).

#### Pre-to-post treatment whole-brain analyses

Whole-brain results were overall consistent with the ROI analyses and are reported in the supplements: the CBT-ERP showed mainly decreased task-related activation, while the I-CBT group showed increased activation after treatment (supplementary tables 7-12).

### Prediction analyses

#### Fear contrast

Both the ITT and PP samples receiving CBT-ERP showed credible evidence for a positive association between baseline task-related activity during the fear contrast and treatment-induced symptom improvement across all ROIs (Figure 3a, supplementary figure 15). In the ITT sample, the I-CBT group showed a negative association between baseline activation in several regions (including bilateral DLPFC, precentral gyrus, superior insula, supramarginal gyrus, middle frontal gyrus, middle temporal gyrus, and right superior temporal gyrus) and symptom improvement, but most of these associations were sensitive to adjusting for medication status. In the PP sample, the I-CBT group showed a positive association between symptom improvement and baseline activity in the bilateral LOC (P+ = 0.95) and left anterior prefrontal cortex (P+ = 0.94), regardless of medication status (supplementary figure 15).

**Figure 3.**
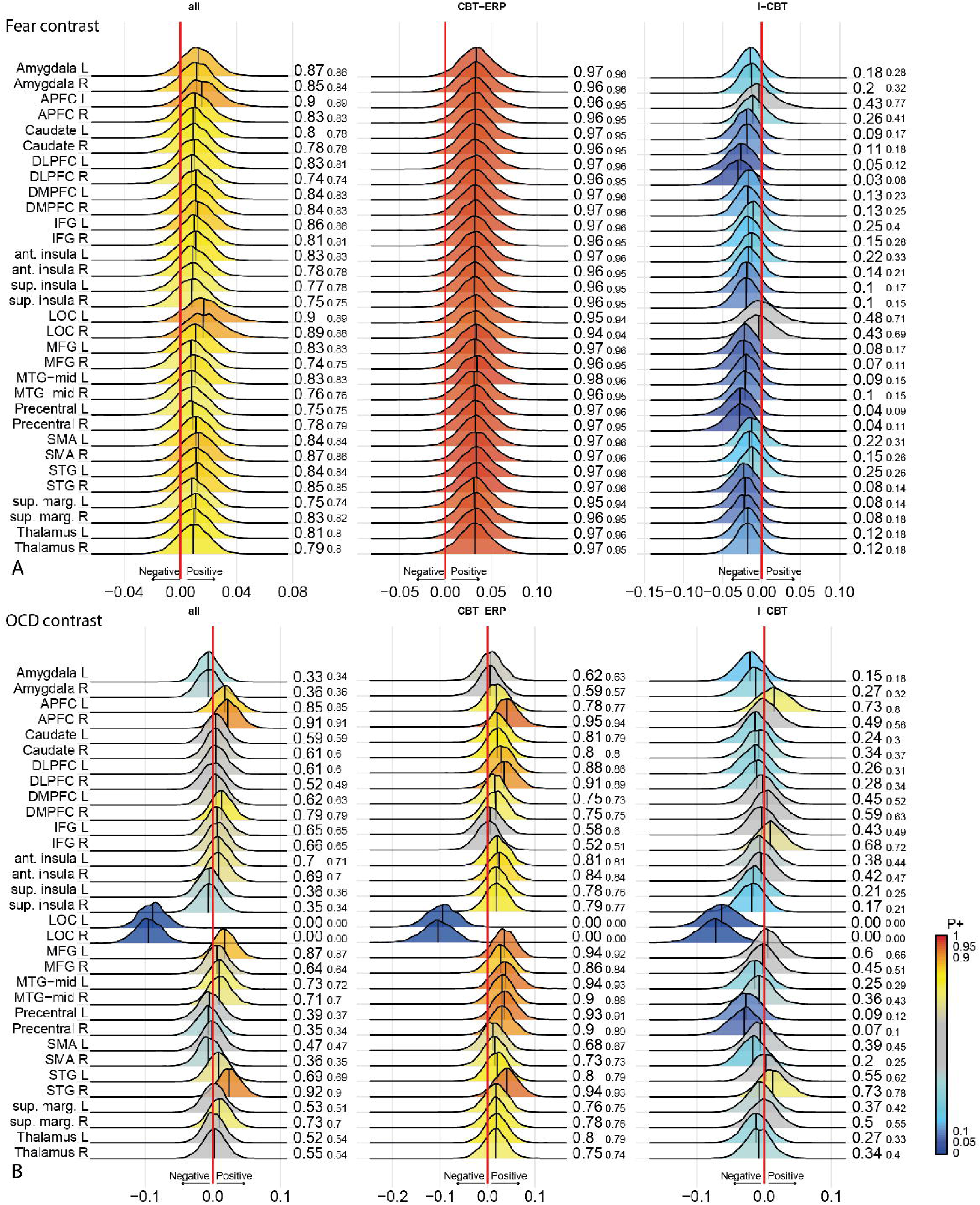
– Bayesian posterior distribution plots of the association between baseline task activation during the fear (a) and OCD (b) contrast and change in OCD symptoms after treatment in the intention-to-treat (ITT) sample. The first column shows the association across both treatment groups. Column two and three show the associations in the CBT-ERP and I-CBT group, respectively. See legend of Figure 1 for an explanation of the color codes. The smaller P+ values represent the posterior probabilities when adjusting for medication status. The meaning of the direction of effects is shown next to the red zero-effect line. See supplementary Table 1 for the definition of the abbreviated regions of interest. All analyses were adjusted for age and sex and baseline YBOCS score.

CBT-ERP responders had higher baseline activity across all ROIs compared to non-responders, with higher credibility in the ITT (supplementary figure 16) than the PP sample (supplementary figure 17), and little influence of medication status. Conversely, I-CBT responders in the ITT sample showed lower baseline activity than non-responders in the bilateral DLPFC (P+ = 0.01), MFG (P+ = 0.04-0.07), precentral gyrus (P+ = 0.03-0.06) and supramarginal gyrus (P+ = 0.05-0.06), with similar results in the PP sample and after medication status adjustment. Across samples and treatment groups, the bilateral LOC showed higher baseline activity in responders, regardless of medication status (P+ > 0.9).

#### OCD contrast

The OCD contrast showed similar patterns to the fear contrast in both ITT and PP samples (Figure 3b, supplementary figure 18), except for the bilateral LOC, which showed strong evidence (P+ < 0.05) for a negative association between baseline task-related activity and symptom improvement in both treatment groups. In the ITT and, to a lesser extent, PP sample, CBT-ERP responders had higher baseline activity than non-responders in the right anterior PFC, bilateral DLPFC, right anterior insula, bilateral precentral gyrus, right superior temporal gyrus, and right supramarginal gyrus (supplementary figures 19-20). Conversely, CBT-ERP responders had lower baseline bilateral LOC activity (P+ = 0.01). I-CBT responders showed credible evidence for lower baseline activity in the bilateral precentral gyrus (P+ = 0.06-0.08) compared to non-responders in the ITT sample. Results for the emotional contrast are presented in the supplements (supplementary figures 21-24).

#### Whole-brain analyses

Whole-brain results are reported in the supplements: the CBT-ERP group showed predominately positive associations between baseline task-related activity and symptom improvement, while the I-CBT group showed relatively few significant associations (supplementary tables 13-15).

## Discussion

The aim of this study was to investigate the effects of two forms of psychotherapy for OCD on neural activation during an emotional processing task and determine whether baseline activity patterns are associated with treatment outcome. Figure 4 gives a schematic summary of the results on the pre-post sample and prediction sample. We found that CBT-ERP and I-CBT had opposite effects on task-related activity. The CBT-ERP group showed evidence for *decreased* activation after treatment across all ROIs, while the I-CBT group showed *increased* activation, with – as hypothesized – the strongest credible evidence for the precentral gyrus. These results were particularly driven by the fear-related pictures. The OCD contrast showed less credible evidence for an effect of treatment. Adjusting for medication status had little impact on these results.

**Figure 4.**
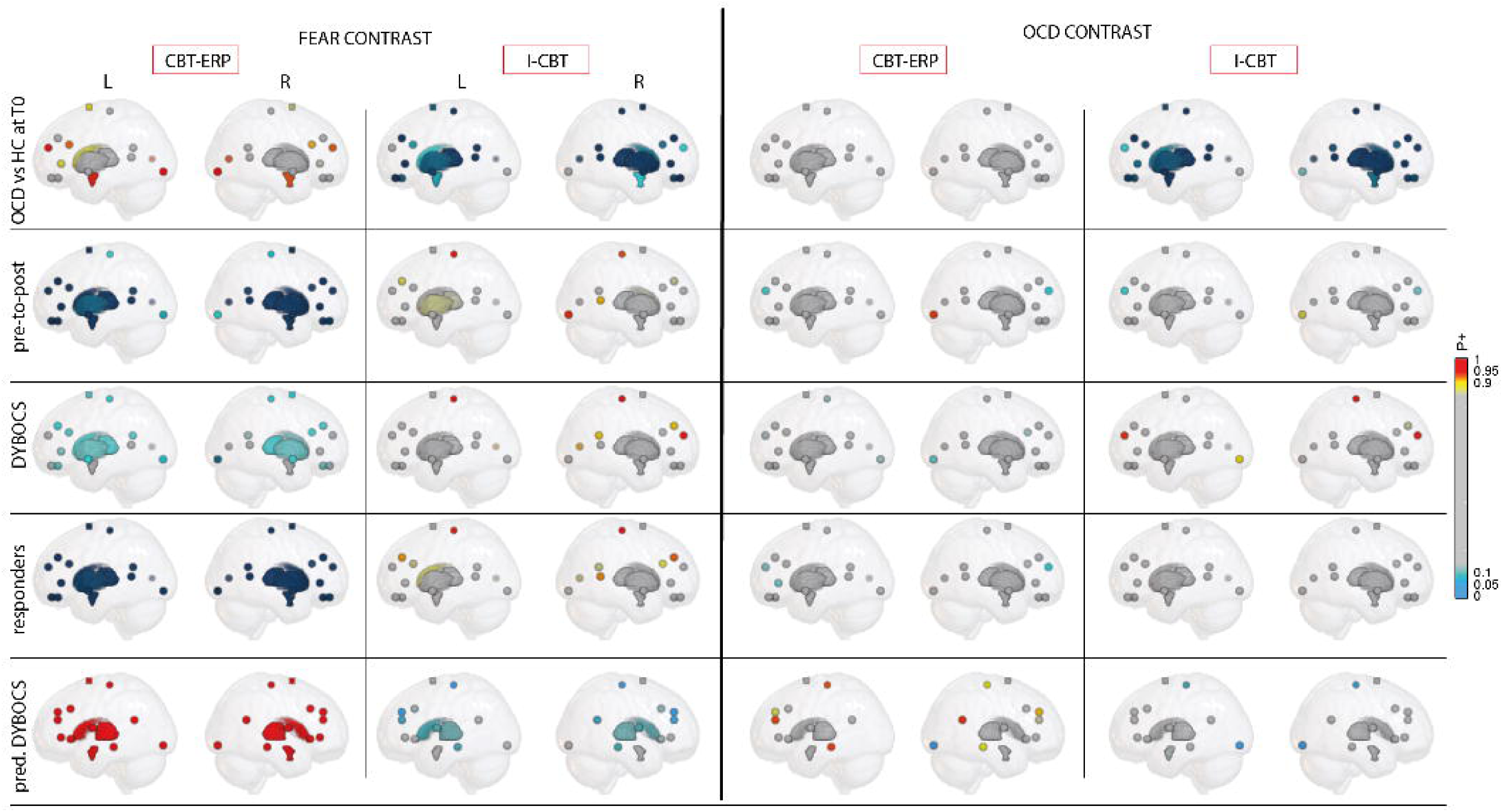
– Schematic overview of the results. Overview of evidence credibility (P+ values) for fear and OCD contrasts across regions of interest, shown separately for CBT-ERP and I-CBT treatments. Blue colors indicate lower activity/negative associations; red-yellow colors indicate higher activity/positive associations; grey indicates no credible evidence. Row 1: Baseline differences in contrast-related activity between each treatment group and healthy controls. Row 2: Pre-to-post treatment changes in activity. Row 3 (ΔYBOCS): Associations between changes in task-related activity and symptom improvement. Row 4 (responders): Activity changes in treatment responders. Row 5 (pred. ΔYBOCS): Associations between baseline activity and symptom change post-treatment.

Higher baseline activation to fear-related pictures was associated with better CBT-ERP response, while baseline activation showed no robust association with I-CBT response. The latter was more sensitive to adjustment for medication status and to whether the PP or ITT sample was considered. Unexpectedly, the two treatment groups, that showed no baseline differences in clinical characteristics, showed differences in task-related activity prior to treatment in the fear contrast, with the CBT-ERP group showing hyperactivity and the I-CBT group showing hypoactivity, relative to healthy controls. It should be stressed that due to randomization *after* the baseline scan, these differences are purely due to chance. While testing for baseline differences is actively discouraged in clinical trials because it can lead to misleading conclusions (Moher et al., 2012), we thought it was important to report differences in activation relative to healthy controls. Nevertheless, the reported results can not merely be ascribed to a regression to the mean as there was evidence for an association between the change in task-related activity and change in symptoms after treatment in both treatment groups and in the CBT-ERP group the effects where stronger in responders than non-responders. Moreover, in each treatment group separately, baseline activity was associated with symptom improvement, albeit in opposite directions. Together, these findings suggest a treatment-induced normalization of activation, supported by the observation that after treatment both groups showed a smaller difference in fear-related activity compared with the healthy controls (supplementary figures 25-26).

In the I-CBT group, particularly the precentral gyrus showed evidence for an association between symptom improvement and an increase in fear-related task activation. While the precentral gyrus is best known for its role in motor control, recent evidence suggests that the precentral gyrus and the broader somatosensory network are involved in emotional processing and implicated in the pathophysiology of OCD (Stein et al., 2019). Viewing of emotional stimuli has been shown to increase excitability of the precentral gyrus (Hajcak et al., 2007) and emotion regulation – particularly reappraisal – is associated with precentral gyrus activation (Morawetz et al., 2017). Furthermore, the precentral gyrus is activated under conditions of doubt during decision making (Naaz et al., 2021) and plays an important role in counterfactual thinking (Suo et al., 2024), imagining how things *could* have happened, which has been shown to be excessive in individuals with OCD (Gillan et al., 2014). I-CBT addresses these cognitive constructs by learning individuals to resolve obsessional doubt and develop greater trust in their sensory perceptions (Aardema et al., 2022). The increase in precentral gyrus activity in association with I-CBT treatment outcome therefore fits with the presumed neurobiological working mechanism. Nevertheless, little research has so far been conducted on the neurobiological mechanisms of action of I-CBT and warrants further scrutiny. Although there was also credible evidence for an association between symptom improvement and change in OCD-related activation of the right precentral gyrus, this association was less robust. We speculate that this might be because participants experienced less distress from these pictures as evidenced by the lower valence rating compared with the fear pictures (see supplementary figure 3).

Effects of CBT-ERP on emotional processing have been studied more elaborately and our results expand upon previous findings of decreased activation after (successful) CBT-ERP in, amongst others, the OFC, thalamus, DLPFC, dACC and caudate nucleus (Poli et al., 2022; Stephenson et al., 2025; Thorsen et al., 2015). Our whole brain analyses confirm a general decrease in fronto-limbic activity after treatment, including in the amygdala and DMPFC. Consistent with the meta-analysis of (Picó-Pérez et al., 2022), our results also showed that successful CBT-ERP was associated with higher pre-treatment activity in fronto-limbic brain areas during the fear contrast, including the IFG, DMPFC and anterior insula. Contrary to our hypothesis, however, higher instead of lower pre-treatment activation in cognitive control areas was associated with CBT-ERP outcome. We speculate that this is explained by our task design that required natural appraisal rather than cognitive reappraisal which relies more heavily on recruitment of these brain areas.

While most regions showed opposite direction of effects of treatment, one striking finding was the highly credible evidence for an association across both treatment groups between lower baseline LOC activation during the OCD contrast and better symptom improvement after treatment. Although less robust in the I-CBT group, there was also credible evidence for an opposite pattern for the fear-related pictures where higher baseline LOC activity was associated with symptom improvement. As extrastriate visual area, the LOC has classically been implicated in object recognition (Grill-Spector, 2003). More recently, studies suggest that the LOC also plays a role in the processing of negative stimuli (Garcia-Garcia et al., 2016; Kuniecki et al., 2018) and may have a driving role in conscious visual processing, where the LOC may be the first node in the circuitry needed to become consciously aware of visual stimuli (Colombari et al., 2024). Interestingly, the study by Colombari and colleagues also showed that LOC activity triggered downstream activity in the precentral gyrus. The LOC and other visual areas are increasingly being implicated in the pathophysiology of OCD with previous studies showing – relative to healthy controls – alterations in functional activity during emotional processing (Dzinalija et al., 2024) and resting-state functional homogeneity (Bruin et al., 2023). A recent study also found that baseline resting-state functional connectivity of the LOC was associated with remission after CBT for OCD (Ikemizu et al., 2025). Together these studies suggest a more important role of the visual system in the pathophysiology of OCD (Gonçalves et al., 2010) than suggested by the prevailing models of OCD (Shephard et al., 2021; Stein et al., 2019). In contrast to the results observed for the LOC, we found that both treatments had overall greater effects on task-related activity during fear stimuli compared to OCD-related stimuli. This may be explained by the fact that the OCD-related visual stimuli reflected a mix of contamination, symmetry, and checking obsessions, and were not personalized to the specific symptom dimensions of our participants. Notably, the opposite associations between symptom improvement and baseline activation in response to fear and OCD-related images in the LOC and other areas highlight the importance of considering these contrasts separately.

Strengths of this study include the direct comparison of two treatment conditions in a randomized controlled trial, the relatively large sample size, and use of Bayesian analysis method allowing integration of shared information across ROIs into one unified model that minimizes multiple comparison corrections and more faithfully reflects the continuous nature of brain activity (Chen et al., 2019). Some limitations may have impacted the results. First, the baseline differences between the groups means that results of the group comparison could still partially reflect regression to the mean and hampers clear-cut comparison between the two treatments. Nevertheless, we argue that this is likely not a substantial effect, given the associations of change in brain activation with symptom reduction in each treatment group. Secondly, the relatively high drop-out rate for the post-treatment MRI (in part due to COVID-restrictions) decreased the statistical power of the pre-post analyses.

The present findings have potential clinical implications for treatment selection in OCD. The differential associations between baseline neural activity and treatment response suggest that individuals with heightened fear-related reactivity may particularly benefit from CBT-ERP, whereas individuals with lower baseline engagement of fear-related and sensory processing networks may respond more favorably to I-CBT. Importantly, both treatments were associated with a normalization of neural activation, albeit from opposite baseline profiles, indicating that distinct therapeutic mechanisms may converge on similar neural outcomes. Although these findings require replication and are not intended to guide clinical decision-making at the individual level, they highlight the potential value of neurobiological markers in advancing more personalized approaches to psychotherapy for OCD when multiple evidence-based interventions are available.

In conclusion, using an emotional task paradigm we show that two forms of psychotherapy for OCD have distinct neurobiological effects that fit with their hypothesized working mechanisms and differentially relate to symptom improvement. During fearful picture viewing, lower baseline activity—particularly in the precentral gyrus and DLPFC—presaged symptom improvement after I-CBT, whereas higher overall baseline activation was associated with improvement following CBT-ERP. Lower LOC activity during processing of OCD-related stimuli was shown to be a robust marker for better treatment outcome across both psychotherapies.

## Data Availability

According to European law (GDPR) data containing potentially identifying or sensitive patient information are restricted; our data involving clinical participants are therefore not freely available but are available from the corresponding author upon reasonable request.

## Acknowledgements

This research was supported by a grant from ZonMW (Grant No. 636310004). We hereby like to thank all the participating trial sites for their help with recruitment and all participants that were willing to participate in the MRI study. We also thank all dedicated therapists and research assessors who contributed to this study.

## Notes

### Competing Interest Statement

The authors have declared no competing interest.

### Clinical Trial

NCT03929081

### Funding Statement

This study was funded by ZonMW (Grant No. 636310004).

### Author Declarations

The ethics committee of VU Medical Center gave ethical approval for this work.

