## Supplementary for "Psychotherapies for obsessive-compulsive disorder have distinct effects on brain activity during emotional processing"

### Supplementary Materials

### Imaging acquisition parameters

On a GE discovery MR750 3T with a 32-channel head coil (General Electric, Milwaukee, U.S.) we acquired T1-weighted structural magnetization-prepared rapid acquisition gradient-echo (MPRAGE; TR = 6.9 ms; TI = 900 ms; TE = 3.0 ms; 256x256 matrix; 1 mm3 isotropic resolution; 168 sections). Task functional MRI (fMRI) consisted of a gradient echo-planar imaging (TR = 2.2 s; TE = 26 ms; 64x64 matrix; field of view 21.1cm; flip angle = 80°) with 42 ascending slices per volume (3.3 x 3.3 mm in-plane resolution; slice thickness = 3.0 mm; interslice gap = 0.3 mm). We also acquired fieldmaps with opposite phase-encoding directions. Scans were acquired within three weeks before and after treatment.

**Supplementary Table 1 - Regions-of-interest and MNI coordinates.**

| | Region of interest | MNI coordinates |
|---|---|---|
| **Pre-post analyses** | | |
| | Amygdala | According to AAL atlas |
| | Caudate nucleus | According to AAL atlas |
| | Dorsal ACC (dACC) | +- 4 / 28 / 24 |
| | Dorsolateral prefrontal cortex (dlPFC) | +- 36 / 42 / 32 |
| | Inferior frontal gyrus (IFG) | +- 48 / 38 / 0 |
| | Inferior insula (inf. insula) | +- 40 / 4 / -10 |
| | Lateral occipital cortex (LOC) | +- 32 / -90 / -10 |
| | Middle occipital cortex (MOC) | 0 / -76 / 6 |
| | Medial prefrontal cortex (mPFC) | +- 4 / 54 / 20 |
| | Middle temporal gyrus / angular gyrus (MTG-ang) | +- 58 / -50 / 8 |
| | Nucleus accumbens (NAcc) | According to AAL atlas |
| | Orbitofrontal cortex (OFC) | +- 14 / 50 / -18 |
| | Precentral gyrus | +- 37 / -23 / 66 |
| | Precuneus | +- 16 / -52 / 20 |
| | Putamen | According to AAL atlas |
| | Subgenual anterior cingulate cortex (sgACC) | +- 4* / 34 / -8 |
| | Supplementary motor area (SMA) | +- 10 / 3 / 71 |
| | Thalamus | According to AAL atlas |
| | Ventromedial prefrontal cortex (vmPFC) | +- 4* / 42 / -18 |
| **Prediction analyses** | | |
| | Amygdala | According to AAL atlas |
| | Anterior prefrontal cortex (aPFC) | +-12 / 68 / 2 |
| | Caudate nucleus | According to AAL atlas |
| | Dorsolateral prefrontal cortex (dlPFC) | +- 36 / 42 / 32 |
| | Dorsomedial prefrontal cortex (dmPFC) | +- 8 / 26 / 36 |
| | Inferior frontal gyrus (IFG) | +- 48 / 38 / 0 |
| | Anterior insula | +- 28 / 26 / -2 |
| | Superior insula | +- 42 / -2 / 12 |
| | Lateral occipital cortex (LOC) | +- 32 / -90 / -10 |
| | Medial frontal gyrus (MFG) | +- 40 / 42 / 22 |
| | Middle temporal gyrus (MTG-mid) | +- 62 / -28 / -12 |
| | Precentral gyrus | +- 37 / -23 / 66 |
| | Supplementary motor area (SMA) | +- 10 / 3 / 71 |
| | Superior temporal gyrus (STG) | +- 58 / -54 / 22 |
| | Supramarginal gyrus | +- 66 / -28 / 28 |
| | Thalamus | According to AAL atlas |

### Fmriprep boilerplate

Results included in this manuscript come from preprocessing performed using fMRIPrep 21.0.1 (Esteban, Markiewicz, et al. (2018); Esteban, Blair, et al. (2018); RRID:SCR_016216), which is based on Nipype 1.6.1 (K. Gorgolewski et al. (2011); K. J. Gorgolewski et al. (2018); RRID:SCR_002502).

Preprocessing of B0 inhomogeneity mappings

A total of 2 fieldmaps were found available within the input BIDS structure for this particular subject. A B0-nonuniformity map (or fieldmap) was estimated based on two (or more) echo-planar imaging (EPI) references with topup (Andersson, Skare, and Ashburner (2003); FSL 6.0.5.1:57b01774).

Anatomical data preprocessing

A total of 2 T1-weighted (T1w) images were found within the input BIDS dataset. All of them were corrected for intensity non-uniformity (INU) with N4BiasFieldCorrection (Tustison et al. 2010), distributed with ANTs 2.3.3 (Avants et al. 2008, RRID:SCR_004757). The T1w-reference was then skull-stripped with a Nipype implementation of the antsBrainExtraction.sh workflow (from ANTs), using OASIS30ANTs as target template. Brain tissue segmentation of cerebrospinal fluid (CSF), white-matter (WM) and gray-matter (GM) was performed on the brain-extracted T1w using fast (FSL 6.0.5.1:57b01774, RRID:SCR_002823, Zhang, Brady, and Smith 2001). A T1w-reference map was computed after registration of 2 T1w images (after INU-correction) using mri_robust_template (FreeSurfer 6.0.1, Reuter, Rosas, and Fischl 2010). Brain surfaces were reconstructed using recon-all (FreeSurfer 6.0.1, RRID:SCR_001847, Dale, Fischl, and Sereno 1999), and the brain mask estimated previously was refined with a custom variation of the method to reconcile ANTs-derived and FreeSurfer-derived segmentations of the cortical gray-matter of Mindboggle (RRID:SCR_002438, Klein et al. 2017). Volume-based spatial normalization to two standard spaces (MNI152NLin6Asym, MNI152NLin2009cAsym) was performed through nonlinear registration with antsRegistration (ANTs 2.3.3), using brain-extracted versions of both T1w reference and the T1w template. The following templates were selected for spatial normalization: FSL's MNI ICBM 152 non-linear 6th Generation Asymmetric Average Brain Stereotaxic Registration Model [Evans et al. (2012), RRID:SCR_002823; TemplateFlow ID: MNI152NLin6Asym], ICBM 152 Nonlinear Asymmetrical template version 2009c [Fonov et al. (2009), RRID:SCR_008796; TemplateFlow ID: MNI152NLin2009cAsym].

Functional data preprocessing

For each of the 2 BOLD runs found per subject (across all tasks and sessions), the following preprocessing was performed. First, a reference volume and its skull-stripped version were generated using a custom methodology of fMRIPrep. Head-motion parameters with respect to the BOLD reference (transformation matrices, and six corresponding rotation and translation parameters) are estimated before any spatiotemporal filtering using mcflirt (FSL 6.0.5.1:57b01774, Jenkinson et al. 2002). The estimated fieldmap was then aligned with rigid-registration to the target EPI (echo-planar imaging) reference run. The field coefficients were mapped on to the reference EPI using the transform. BOLD runs were slice-time corrected to 1.07s (0.5 of slice acquisition range 0s-2.15s) using 3dTshift from AFNI (Cox and Hyde 1997, RRID:SCR_005927). The BOLD reference was then co-registered to the T1w reference using bbregister (FreeSurfer) which implements boundary-based registration (Greve and Fischl 2009). Co-registration was configured with six degrees of freedom. Several confounding time-series were calculated based on the preprocessed BOLD: framewise displacement (FD), DVARS and three region-wise global signals. FD was computed using two formulations following Power (absolute sum of relative motions, Power et al. (2014)) and Jenkinson (relative root mean square displacement between affines, Jenkinson et al. (2002)). FD and DVARS are calculated for each functional run, both using their implementations in Nipype (following the definitions by Power et al. 2014). The three global signals are extracted within the CSF, the WM, and the whole-brain masks. Additionally, a set of physiological regressors were extracted to allow for component-based noise correction (CompCor, Behzadi et al. 2007). Principal components are estimated after high-pass filtering the preprocessed BOLD time-series (using a discrete cosine filter with 128s cut-off) for the two CompCor variants: temporal (tCompCor) and anatomical (aCompCor). tCompCor components are then calculated from the top 2% variable voxels within the brain mask. For aCompCor, three probabilistic masks (CSF, WM and combined CSF+WM) are generated in anatomical space. The implementation differs from that of Behzadi et al. in that instead of eroding the masks by 2 pixels on BOLD space, the aCompCor masks are subtracted a mask of pixels that likely contain a volume fraction of GM.

This mask is obtained by dilating a GM mask extracted from the FreeSurfer's aseg segmentation, and it ensures components are not extracted from voxels containing a minimal fraction of GM. Finally, these masks are resampled into BOLD space and binarized by thresholding at 0.99 (as in the original implementation). Components are also calculated separately within the WM and CSF masks. For each CompCor decomposition, the k components with the largest singular values are retained, such that the retained components' time series are sufficient to explain 50 percent of variance across the nuisance mask (CSF, WM, combined, or temporal). The remaining components are dropped from consideration. The head-motion estimates calculated in the correction step were also placed within the corresponding confounds file. The confound time series derived from head motion estimates and global signals were expanded with the inclusion of temporal derivatives and quadratic terms for each (Satterthwaite et al. 2013). Frames that exceeded a threshold of 0.5 mm FD or 1.5 standardised DVARS were annotated as motion outliers. The BOLD time-series were resampled into standard space, generating a preprocessed BOLD run in MNI152NLin6Asym space. First, a reference volume and its skull-stripped version were generated using a custom methodology of fMRIPrep. The BOLD time-series were resampled onto the following surfaces (FreeSurfer reconstruction nomenclature): fsnative, fsaverage5. Automatic removal of motion artifacts using independent component analysis (ICA-AROMA, Pruim et al. 2015) was performed on the preprocessed BOLD on MNI space time-series after removal of non-steady state volumes and spatial smoothing with an isotropic, Gaussian kernel of 6mm FWHM (full-width half-maximum). Corresponding "non-aggresively" denoised runs were produced after such smoothing. Additionally, the "aggressive" noise-regressors were collected and placed in the corresponding confounds file. All resamplings can be performed with a single interpolation step by composing all the pertinent transformations (i.e. head-motion transform matrices, susceptibility distortion correction when available, and co-registrations to anatomical and output spaces). Gridded (volumetric) resamplings were performed using antsApplyTransforms (ANTs), configured with Lanczos interpolation to minimize the smoothing effects of other kernels (Lanczos 1964). Non-gridded (surface) resamplings were performed using mri_vol2surf (FreeSurfer).

Many internal operations of fMRIPrep use Nilearn 0.8.1 (Abraham et al. 2014, RRID:SCR_001362), mostly within the functional processing workflow. For more details of the pipeline, see the section corresponding to workflows in fMRIPrep's documentation.

Copyright Waiver

The above boilerplate text was automatically generated by fMRIPrep with the express intention that users should copy and paste this text into their manuscripts unchanged. It is released under the CC0 license.

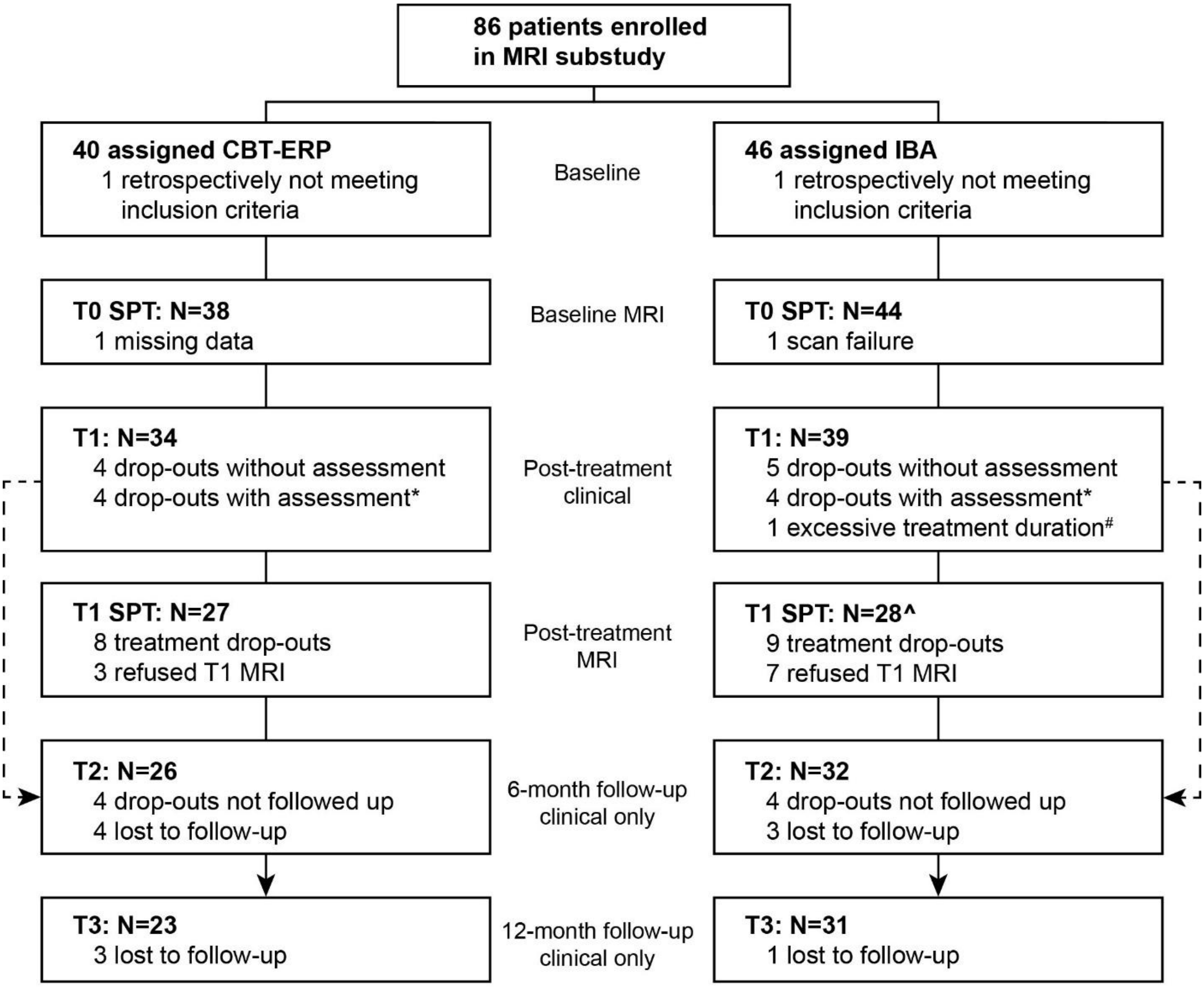

**Supplementary figure 1 – flowchart**. *Treatment drop-outs with clinical data at T1 were included in the intention-to-treat prediction analysis of T1, but not followed up. #One participant with a long treatment duration (>1 year) was included in intention-to-treat prediction analysis only; sensitivity analysis that included this participant revealed no differences in the reported results (data not shown). ^One participant inadvertently received the same version of the task at both timepoints. Sensitivity analyses without this participant had no effect on the reported results (data not shown).

| **Supplementary table 2 – Demographic and clinical characteristics of the intention-to-treatment (ITT), per protocol (PP) and drop-out sample** | | | | |
|---|---|---|---|---|
| | ITT sample (n=73) (n=73) | PP sample (n=65) | Drop-out sample (n=9) | ITT vs Drop-out statistics |
| **Sex (female)** | 41 (56.2%) | 36 (55.4%) | 5 (55.6%) | $\chi_{(1)}$=0, P=1 |
| **Age (years)** | 32.9 ± 10.6 | 33.5 ± 10.9 | 32.6 ± 11.4 | $t_{(1)}$ = -0.09, P = 0.93 |
| **Education classification*** | | | | |
| **1** | 1 (1.4%) | 1 (1.5%) | 0 (0%) | |
| **2** | 0 (0%) | 0 (0%) | 0 (0%) | |
| **3** | 0 (0%) | 0 (0%) | 0 (0%) | |
| **4** | 1 (1.4%) | 1 (1.5%) | 0 (0%) | |
| **5** | 21 (28.8%) | 17 (26.2%) | 3 (33.3%) | |
| **6** | 20 (41.1%) | 28 (43.1%) | 3 (33.3%) | |
| **7** | 20 (27.4%) | 18 (27.7%) | 3 (33.3%) | |
| **YBOCS T0** | 24 ± 4.2 | 24 ± 4.2 | 25.7 ± 2.9 | $t_{(1)}$ = 1.52, P = 0.15 |
| **YBOCS T1** | 15.4 ± 7.6 | 14.8 ± 7.6 | - | - |
| **Responder** | 37 (50.7%) | 34 (52.3%) | - | - |
| **OVIS T0** | 4.9 ± 1.3 | 4.8 ± 1.2 | 5.1 ± 1.4 | $t_{(1)}$ = 0.29, P = 0.78 |
| **BDI T0** | 14.8 ± 8.5 | 14.4 ± 8.5 | 17 ± 7.6 | $t_{(1)}$ = 0.82, P = 0.43 |
| **Comorbid disorders** | 31 (42.5%) | 25 (38.5%) | 6 (66.7%) | $\chi_{(1)}$=1.04, P=0.31 |
| **Depression** | 8 (11.0%) | 7 (10.8%) | 3 (33.3%) | $\chi_{(1)}$=1.8, P=0.18 |
| **Anxiety** | 16 (21.9%) | 14 (21.5%) | 3 (33.3%) | $\chi_{(1)}$=0.12, P=0.73 |
| **Symptom dimensions** | | | | |
| **Symmetry** | 56 (76.7%) | 49 (75.4%) | 7 (77.8%) | $\chi_{(1)}$=0, P=1 |
| **Cleaning** | 54 (74.0%) | 48 (73.8%) | 7 (77.8%) | $\chi_{(1)}$=0, P=1 |
| **Checking** | 62 (84.9%) | 54 (83.1%) | 8 (88.9%) | $\chi_{(1)}$=0, P=1 |
| **OCD onset** | | | | $\chi_{(1)}$=0, P=1 |
| **early** | 45 (61.6%) | 38 (58.5%) | 6 (66.7%) | |
| **late** | 28 (38.4%) | 27 (41.5%) | 3 (33.3%) | |
| **medicated** | 31 (42.5%) | 26 (40.0%) | 4 (44.4%) | $\chi_{(1)}$=0, P=1 |
| **T0-T1 interval (days)** | 199.2 ± 50 | 208 ± 42.9 | - | - |
| **T0 distress rating before task** | 41.1 ± 25.5 | 42.1 ± 25.8 | 53.6 ± 19.3 | $t_{(1)}$ = 1.57, P = 0.15 |
| **T0 distress rating after task** | 42.4 ± 27.6 | 43.3 ± 27.8 | 63.3 ± 10.8 | $t_{(1)}$ = 3.70, P = 0.003 |
| **T1 distress rating before task** | 26.2 ± 24.7 | 26.6 ± 24.7 | | - |
| **T1 distress rating after task** | 30.2 ± 26.9 | 30.8 ± 26.9 | | - |
| **T0 FD (mm)** | 0.13 ± 0.07 | 0.12 ± 0.06 | 0.16 ± 0.14 | $t_{(1)}$ = 0.66, P = 0.53 |
| **T1 FD (mm)** | 0.12 ± 0.06 | 0.12 ± 0.06 | - | - |
| **For the ITT and PP sample post-treatment clinical data was available. The drop-out sample discontinued treatment and did not return for an exit meeting with clinical evaluation.** | | | | |

**Supplementary table 3 – Demographic and clinical characteristics of the per protocol (PP) sample for prediction analyses**

| | PP Prediction sample (N=65) | | |
|---|---|---|---|
| | CBT-ERP (n=30) | I-CBT (n=35) | Statistics |
| **Sex (female)** | 17 (56.7%) | 19 (54.3%) | $\chi_{(1)}$=0, P=1 |
| **Age (years)** | 32.4 ± 11.5 | 34.4 ± 10.4 | $t_{(1)}$ = -0.71, P = 0.48 |
| **Education classification*** | | | P=0.32 |
| **1** | 1 (3.3%) | 0 (0%) | |
| **2** | 0 (0%) | 0 (0%) | |
| **3** | 0 (0%) | 0 (0%) | |
| **4** | 1 (3.3%) | 0 (0%) | |
| **5** | 9 (30.0%) | 8 (22.9%) | |
| **6** | 13 (43.3%) | 15 (42.9%) | |
| **7** | 6 (20.0%) | 12 (34.3%) | |
| **YBOCS T0** | 23.5 ± 3.6 | 24.5 ± 4.7 | $t_{(1)}$ = -1.01, P = 0.31 |
| **YBOCS T1** | 12.5 ± 7.1 | 16.8 ± 7.6 | $t_{(1)}$ = -2.35, P = 0.02 |
| **Responder** | 17 (56.7%) | 17 (48.6%) | $\chi_{(1)}$=0.16, P=0.69 |
| **OVIS T0** | 4.6 ± 1 | 5 ± 1.3 | $t_{(1)}$ = -1.46, P = 0.15 |
| **BDI T0** | 14.7 ± 9.5 | 14.2 ± 7.8 | $t_{(1)}$ = 0.21, P = 0.83 |
| **Comorbid disorders** | 13 (43.3%) | 12 (34.3%) | $\chi_{(1)}$=0.24, P=0.62 |
| **Depression** | 3 (10.0%) | 4 (11.4%) | $\chi_{(1)}$=0, P=1 |
| **Anxiety** | 9 (30.0%) | 5 (14.3%) | $\chi_{(1)}$=1.52, P=0.21 |
| **Symptom dimensions** | | | |
| **Symmetry** | 22 (73.3%) | 27 (77.1%) | $\chi_{(1)}$=0.004, P=0.95 |
| **Cleaning** | 21 (70.0%) | 27 (77.1%) | $\chi_{(1)}$=0.14, P=0.71 |
| **Checking** | 24 (80.0%) | 30 (85.7%) | $\chi_{(1)}$=0.08 P=0.80 |
| **OCD onset** | | | $\chi_{(1)}$=0.28, P=0.6 |
| **early** | 16 (53.3%) | 22 (62.9%) | |
| **late** | 14 (46.7%) | 13 (37.1%) | |
| **medicated** | 12 (40.0%) | 14 (40.0%) | $\chi_{(1)}$=0, P=1 |
| **T0 distress rating before task** | 44.1 ± 27.1 | 40.6 ± 25 | $t_{(1)}$ = 0.50, P = 0.62 |
| **T0 distress rating after task** | 48 ± 29.4 | 39.9 ± 26.5 | $t_{(1)}$ = 1.04, P = 0.31 |
| **T0 FD (mm)** | 0.12 ± 0.06 | 0.12 ± 0.06 | $t_{(1)}$ = -0.23, P = 0.81 |

Abbreviations: YBOCS = Yale-Brown Obsessive-Compulsive Scale, OVIS = Overvalued Ideas Scale, BDI = Beck Depression Inventory. FD = Framewise Displacement

**Supplementary table 4 – demographic and clinical characteristics of the pre-to-post treatment and drop-out sample**

| | Pre-post sample (n=55) | Drop-out sample (n=27) | Statistics |
|---|---|---|---|
| **Sex (female)** | 30 (54.5%) | 15 (55.6%) | $\chi_{(1)}$=0, P=1 |
| **Age (years)** | 34.1 ± 11.2 | 31 ± 8.6 | $t_{(1)}$ = -1.39, P = 0.17 |
| **Education classification*** | | | P=0.16 |
| **1** | 1 (1.8%) | 0 (0%) | |
| **2** | 0 (0%) | 0 (0%) | |
| **3** | 0 (0%) | 0 (0%) | |
| **4** | 1 (1.8%) | 1 (3.7%) | |
| **5** | 12 (21.8%) | 12 (44.4%) | |
| **6** | 25 (45.5%) | 7 (25.9%) | |
| **7** | 16 (29.1%) | 7 (25.9%) | |
| **YBOCS T0** | 23.9 ± 4.1 | 25.6 ± 5.6 | $t_{(1)}$ = -1.37, P = 0.18 |
| **Responder** | 32 (58.2%) | - | - |
| **OVIS T0** | 4.9 ± 1.2 | 5.1 ± 1.5 | $t_{(1)}$ = 0.67, P = 0.51 |
| **BDI T0** | 14 ± 8.6 | 18.1 ± 10.4 | $t_{(1)}$ = 1.76, P = 0.09 |
| **Comorbid disorders** | 20 (36.4%) | 16 (59.3%) | $\chi_{(1)}$=3.0, P=0.08 |
| **Depression** | 4 (7.3%) | 6 (22.2%) | $\chi_{(1)}$=2.5, P=0.11 |
| **Anxiety** | 10 (18.2%) | 9 (33.3%) | $\chi_{(1)}$=1.5, P=0.2 |
| **Symptom dimensions** | | | |
| **Symmetry** | 43 (78.2%) | 21 (77.8%) | |
| **Cleaning** | 39 (70.9%) | 19 (70.4%) | |
| **Checking** | 45 (81.8%) | 24 (88.9%) | |
| **OCD onset** | | | $\chi_{(1)}$=3.8, P=0.05 |
| **early** | 29 (52.7%) | 21 (77.8%) | |
| **late** | 26 (47.3%) | 6 (22.2%) | |
| **medicated** | 22 (40.0%) | 14 (51.9%) | $\chi_{(1)}$=0.61, P=0.44 |
| **T0 distress rating before task** | 44.1 ± 27.1 | 40.6 ± 25 | $t_{(1)}$ = -0.05, P = 0.97 |
| **T0 distress rating after task** | 48 ± 29.4 | 45.9 ± 24.9 | $t_{(1)}$ = 0.62, P = 0.52 |
| **T0 FD (mm)** | 0.12 ± 0.06 | 0.16 ± 0.1 | $t_{(1)}$ = 2.2, P = 0.03 |

The attrition sample refers to participants that did not return for an MRI scan after treatment, either because they discontinued treatment or did not wish to be rescanned. Abbreviations: YBOCS = Yale-Brown Obsessive-Compulsive Scale, OVIS = Overvalued Ideas Scale, BDI = Beck Depression Inventory. FD = Framewise Displacement

| Supplementary Table 5 –change in clinical measures after CBT-ERP and I-CBT treatment | | | | | |
|---|---|---|---|---|---|
| | CBT-ERP | I-CBT | Statistics | | |
| | M ± SD | M ± SD | B [SE] | 95% CI | P |
| **Pre-post sample (N=55)** | | | | | |
| **YBOCS** | | | | | |
| **T0** | 23.67 ± 3.57 | 24.21 ± 4.58 | | | |
| **T1** | 11.59 ± 6.56 | 16.57 ± 6.88 | 4.4 [1.7] | 1.10 \| 7.76 | 0.01 |
| **OVIS** | | | | | |
| **T0** | 4.62 ± 0.98 | 5.17 ± 1.38 | | | |
| **T1** | 2.70 ± 1.41 | 3.26 ± 1.24 | 0.004 [0.37] | -0.71 \| 0.72 | 0.99 |
| **BDI** | | | | | |
| **T0** | 14.04 ± 9.13 | 13.96 ± 8.19 | | | |
| **T1** | 10.42 ± 10.88 | 10.50 ± 7.14 | 0.07 [2.3] | -4.47 \| 4.61 | 0.98 |
| **ITT sample (N=73)** | | | | | |
| **YBOCS** | | | | | |
| **T0** | 23.53 ± 3.59 | 24.46 ± 4.63 | | | |
| **T1** | 13.38 ± 7.37 | 17.21 ± 7.46 | 2.89 [1.66] | 0.09 \| -0.35 | 0.09 |
| **OVIS** | | | | | |
| **T0** | 4.80 ± 1.23 | 5.05 ± 1.30 | | | |
| **T1** | 3.03 ± 1.49 | 3.37 ± 1.31 | 0.09 [0.30] | -0.50 \| 0.68 | 0.76 |
| **BDI** | | | | | |
| **T0** | 14.65 ± 9.42 | 14.85 ± 7.68 | | | |
| **T1** | 11.03 ± 10.35 | 11.17 ± 6.80 | -0.49 [1.97] | -4.34 \| 3.38 | 0.80 |
| **PP sample (N=65)** | | | | | |
| **YBOCS** | | | | | |
| **T0** | 23.47 ± 3.62 | 24.51 ± 4.70 | | | |
| **T1** | 12.53 ± 7.12 | 16.83 ± 7.58 | 3.24 [1.7] | -0.08 \| 6.58 | 0.06 |
| **OVIS** | | | | | |
| **T0** | 4.62 ± 1.01 | 5.05 ± 1.34 | | | |
| **T1** | 2.90 ± 1.46 | 3.26 ± 1.26 | -0.08 [0.33] | -0.72 \| 0.57 | 0.82 |
| **BDI** | | | | | |
| **T0** | 14.67 ± 9.45 | 14.20 ± 7.79 | | | |
| **T1** | 10.52 ± 10.57 | 10.56 ± 6.74 | -0.003 [2.20] | -4.30 \| 4.34 | 0.99 |

Measure are presented as mean ± standard deviation. Abbreviations: CBT-ERP = cognitive behavioral therapy with exposure and relapse prevention therapy, I-CBT = inference-based cognitive behavioral therapy. ITT = Intention-to-treat sample, PP = Per protocol sample, YBOCS = Yale-Brown Obsessive-compulsive Scale, OVIS = Overvalued Ideas Scale, BDI = Beck Depression Inventory,

### Distress and picture ratings

Across treatment groups, distress ratings decreased significantly from pre-to-post treatment, both the rating before (B[SE] = -19.5[5.6], P= 0.001) and after performing the task (B[SE] = -18.8[5.9], P= 0.002). There were no between-group differences in the pre-to-post treatment decrease in distress (distress before task: B[SE] = 10.0[7.6], P= 0.194; distress after task: B[SE] = 15.5[7.8], P= 0.053). Ordinal mixed model analyses of the picture ratings showed no differences across time or between the treatment groups in the rating of the fear or OCD pictures; only a decrease over time in the rating of the scrambled pictures). See supplementary table 6 and supplementary Figures 2-3.

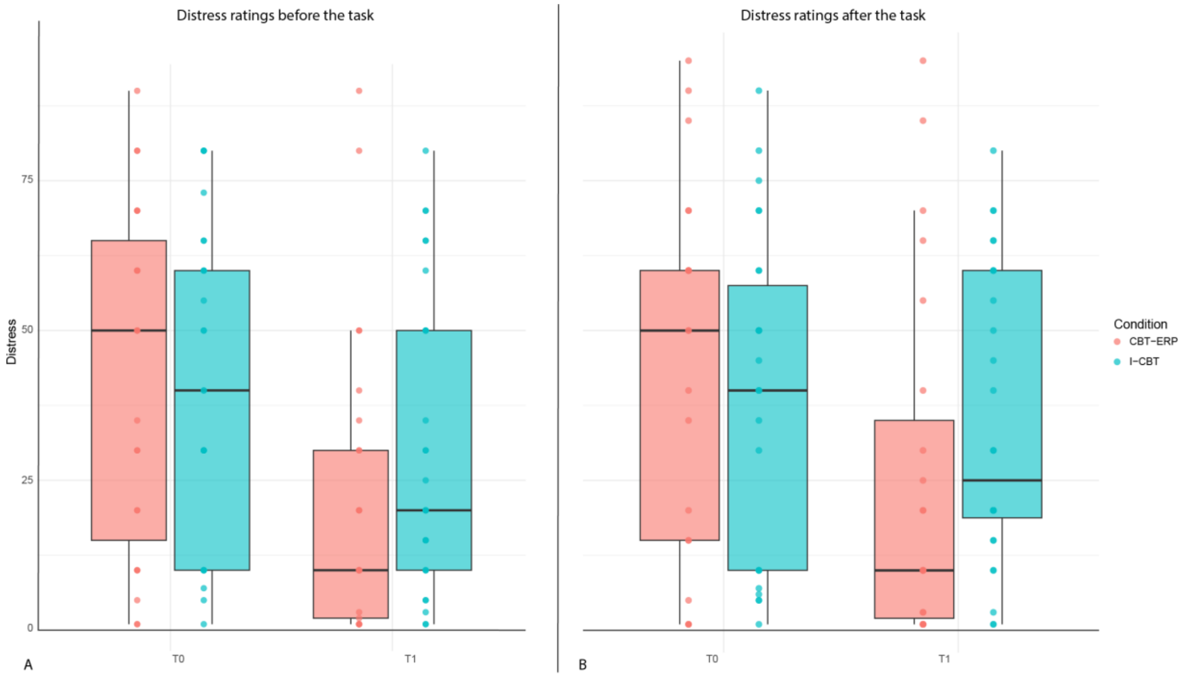

**Supplementary Figure 2 – Distress ratings at pre-treatment and post-treatment MRI before (a) and after (b) the task fMRI.** Ratings were available for N=21 CBT-ERP and N=26 I-CBT participants.

**Supplementary table 6 - ordinal mixed model analyses on picture ratings**

| | B [SE] | 95% CI | P |
|---|---|---|---|
| **Fear** | | | |
| **Time** | 0.01 [0.70] | -1.37 \| 1.41 | 0.97 |
| **Time * Condition** | 0.59 [0.98] | -1.33 \| 2.51 | 0.55 |
| **OCD** | | | |
| **Time** | -0.71 [0.86] | -2.40 \| 0.98 | 0.41 |
| **Time * Condition** | -0.99 [1.20] | -3.35 \| 1.37 | 0.41 |
| **Scrambled** | | | |
| **Time** | -0.55 [0.003] | -0.56 \| -0.55 | <0.001 |
| **Time * Condition** | 0.42 [1.01] | -1.55 \| 2.40 | 0.68 |

Data was available from 40 participants (CBT-ERP n=19, I-CBT n=21)

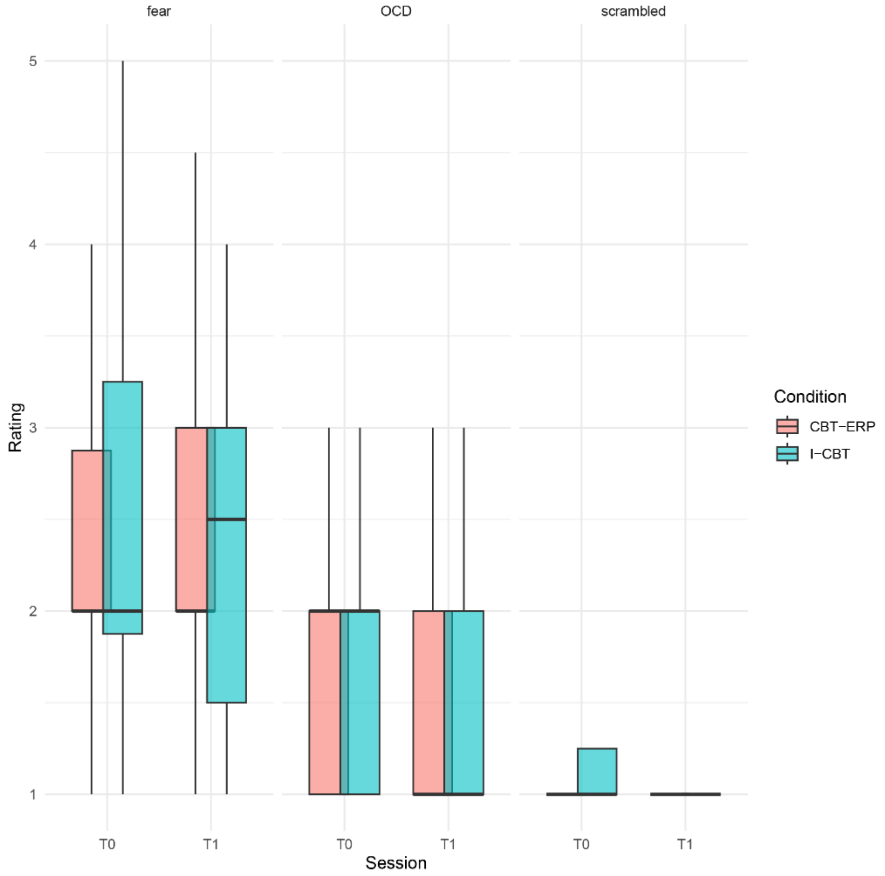

**Supplementary Figure 3 – Picture ratings (outside the scanner) before and after treatment for both treatment conditions**. Data was available from 40 participants (CBT-ERP n=19, I-CBT n=21).

### Pre-treatment group differences in activation

Region-of-interest analyses at baseline (i.e. before treatment commenced) using the pre-to-post sample (N=55) showed very strong evidence for higher activation in the CBT-ERP group compared with the I-CBT group in almost all areas (supplementary figures 4-6). This was mainly observed in the the fear contrast (P+ ranging from 0.84 to 0.99). Relative to healthy controls (N=42), there was no credible evince for differences in activity with the CBT-ERP group in the OCD or emotional contrast. In the fear contrast there were several regions that showed evidence for higher activation in the CBT-ERP group compared with healthy controls, including the bilateral amygdala, lateral occipital cortex (LOC) and middle prefrontal cortex. Across all contrasts and all ROIs except the LOC, the I-CBT group showed strong credible evidence for lower activation relative to healthy controls.

### Pre-treatment group differences in activation

contrast: fear

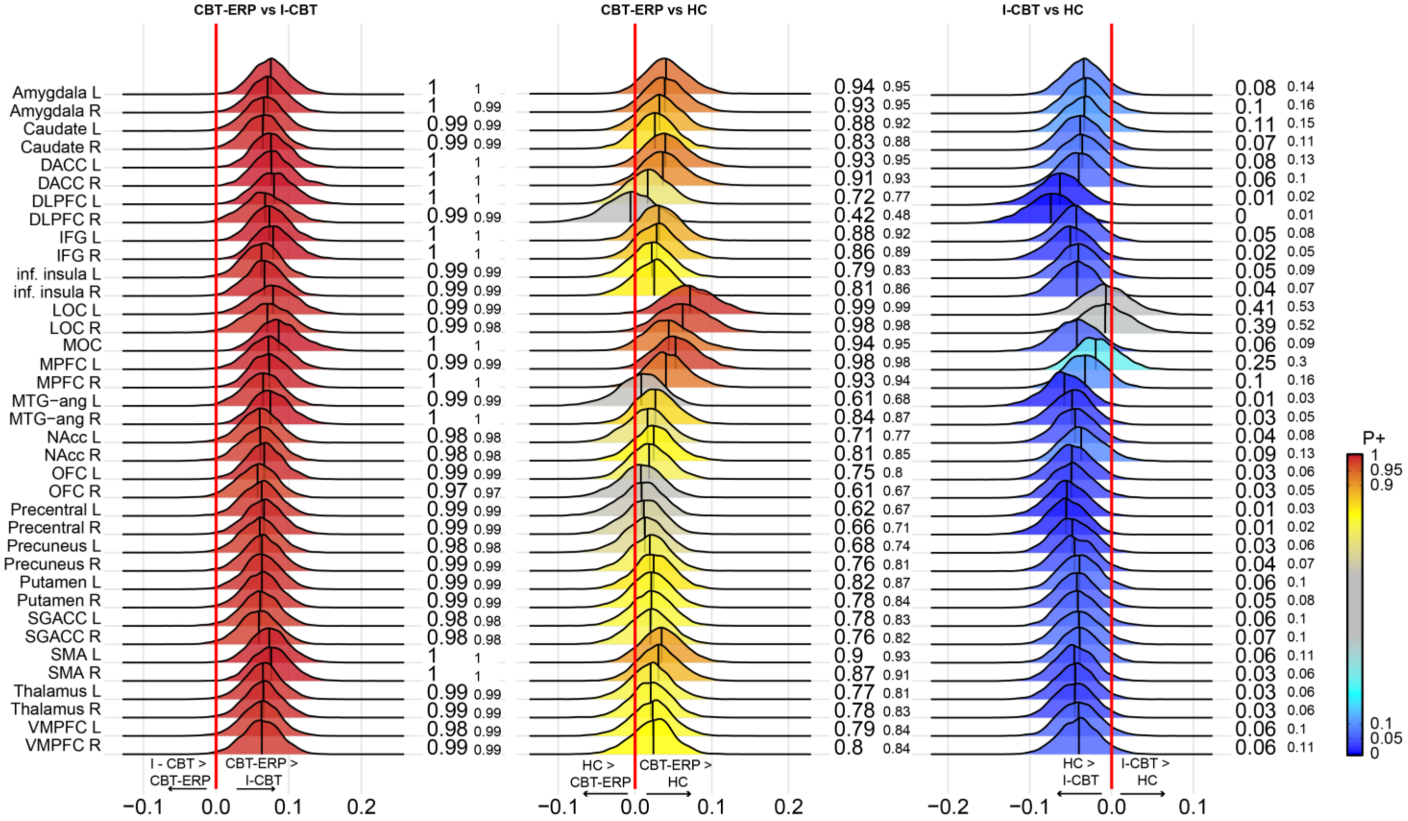

**Supplementary Figure 4 - pre-treatment differences in activation in the fear contrast in the CBT-ERP and I-CBT treatment groups and healthy controls.** The posterior distribution communicates the credibility of an effect. Positive posterior probabilities (*P*+) are shown next to each distribution and color coded. *P*+ values ≥0.90 indicate moderate to very high credibility for a positive effect, P+ ≤0.10 indicate moderate to very high credibility for a negative effect. The meaning of the direction of effects are shown next to the green zero-effect line. See supplementary Table 1 for the definition of the abbreviated regions of interest.

### Pre-treatment group differences in activation

Contrast: OCD

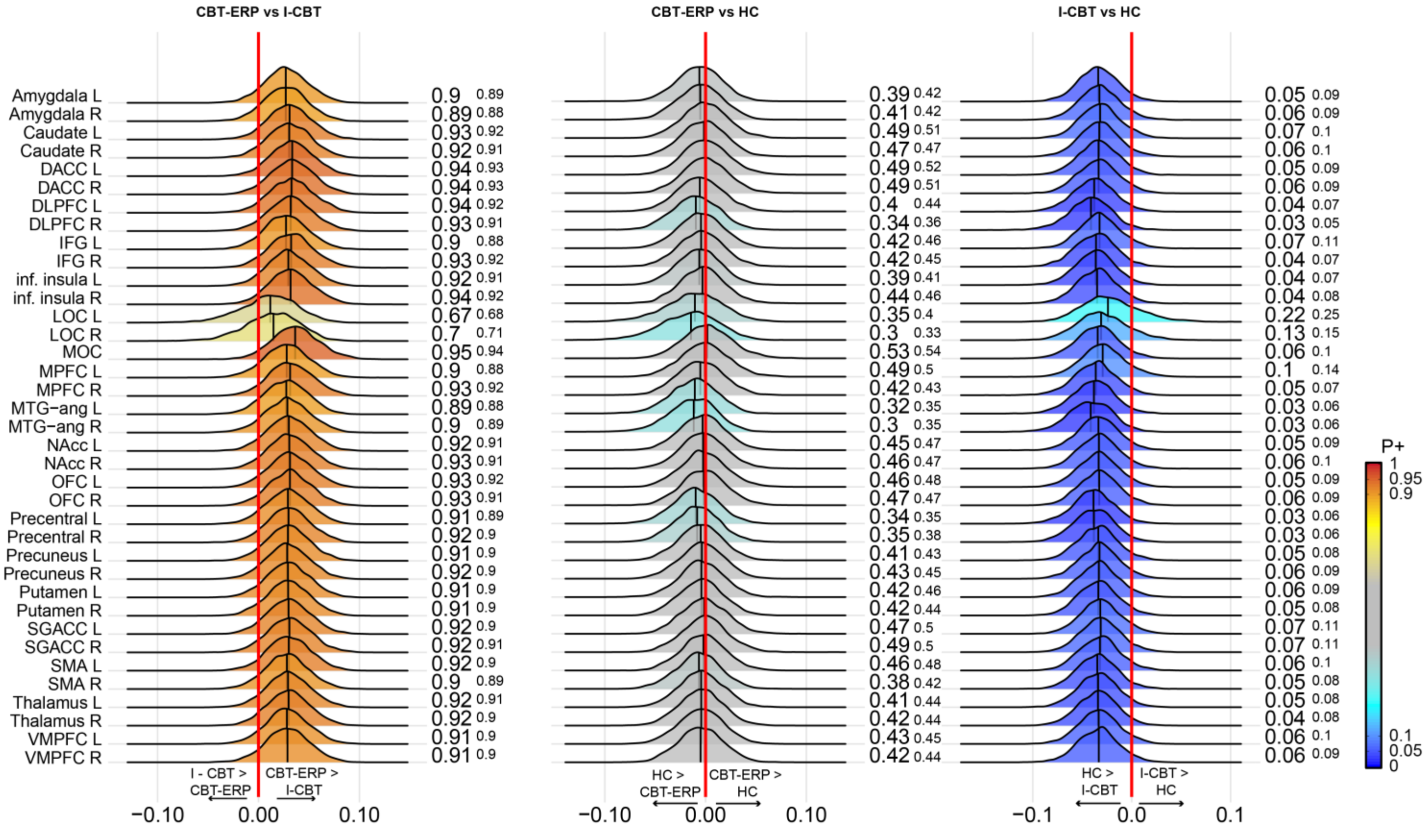

**Supplementary Figure 5 - pre-treatment differences in activation in the OCD contrast in the CBT-ERP and I-CBT treatment groups and healthy controls.** The posterior distribution communicates the credibility of an effect. Positive posterior probabilities (*P*+) are shown next to each distribution and color coded. *P*+ values ≥0.90 indicate moderate to very high credibility for a positive effect, P+ ≤0.10 indicate moderate to very high credibility for a negative effect. The meaning of the direction of effects are shown next to the green zero-effect line. See supplementary Table 1 for the definition of the abbreviated regions of interest.

### Pre-treatment group differences in activation

Contrast: emotion

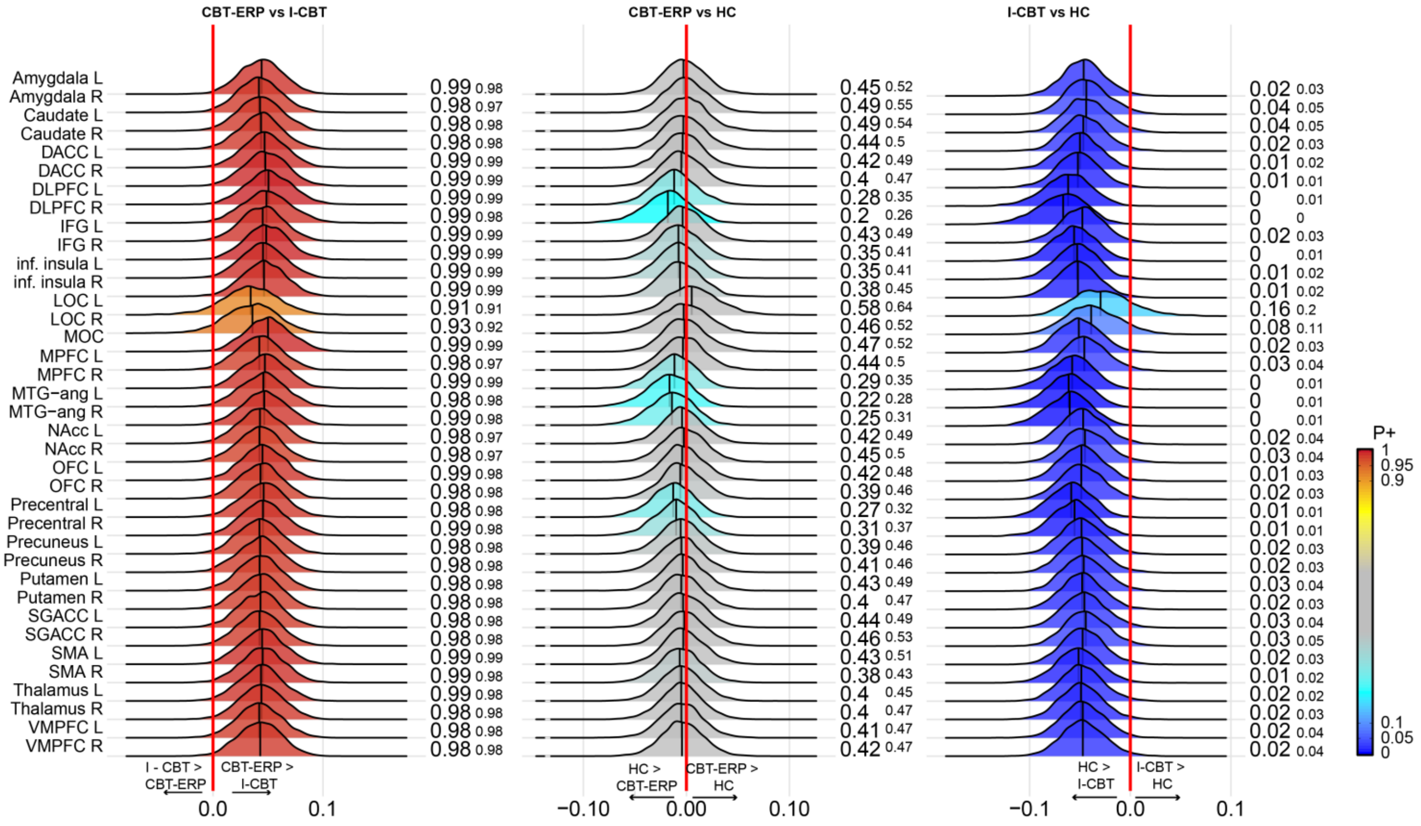

**Supplementary Figure 6 - pre-treatment differences in activation in the emotion contrast in the CBT-ERP and I-CBT treatment groups and healthy controls.** The posterior distribution communicates the credibility of an effect. Positive posterior probabilities (*P*+) are shown next to each distribution and color coded. *P*+ values ≥0.90 indicate moderate to very high credibility for a positive effect, P+ ≤0.10 indicate moderate to very high credibility for a negative effect. The meaning of the direction of effects are shown next to the green zero-effect line. See supplementary Table 1 for the definition of the abbreviated regions of interest.

### Difference in pre-to-post treatment changes between responders and non-responders to CBT-ERP

Contrast: FEAR

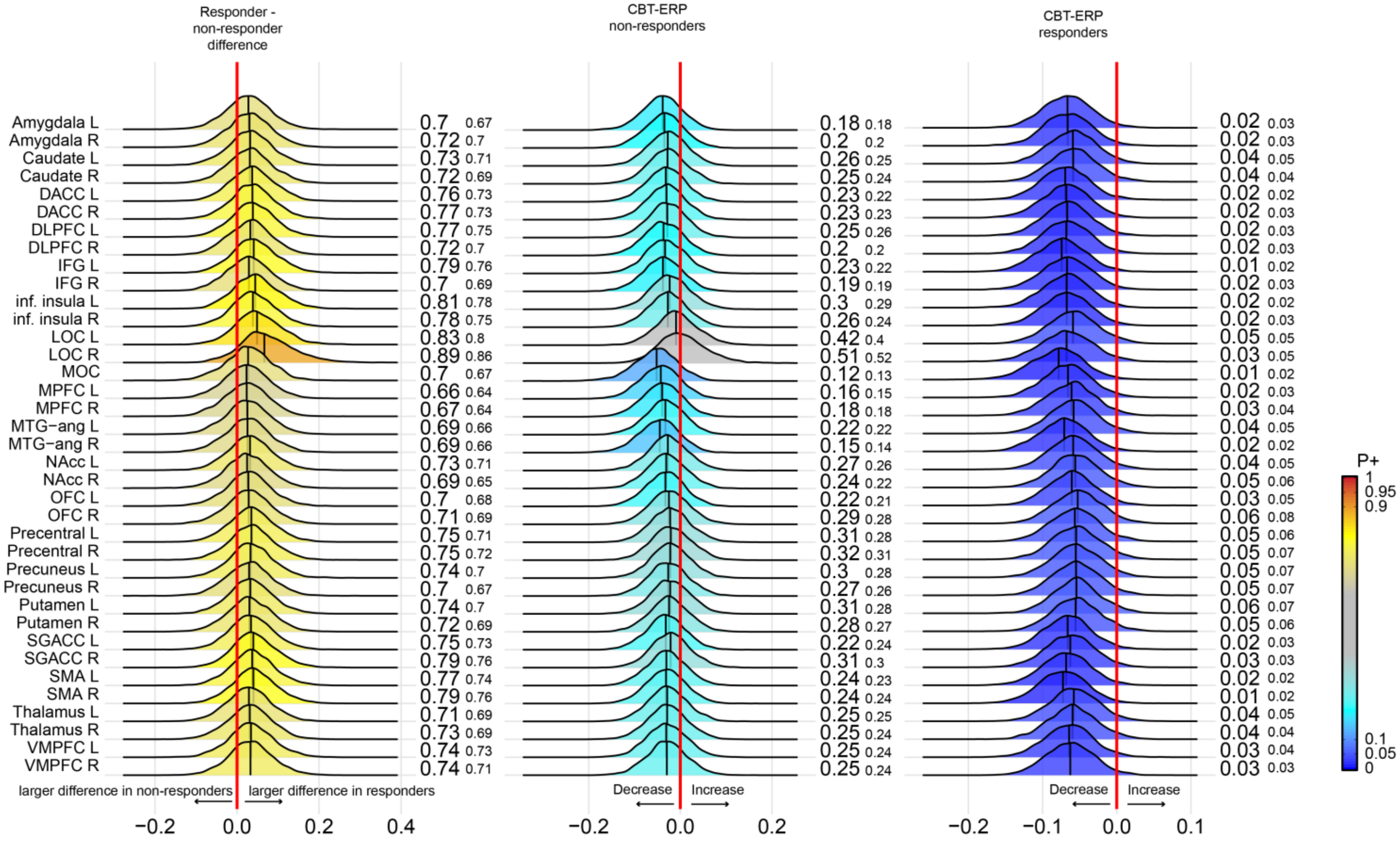

**Supplementary Figure 7 – Bayesian posterior distribution plots of the pre-to-post treatment differences in task activation during the fear contrast between responders and non-responders to CBT-ERP.**
The first column shows the relative differences between responders and non-responders to CBT-ERP in the changes in task related activation after treatment. Column two and three show the changes from pre-to-post treatment in the non-responders and responders, respectively. The posterior distribution communicates the credibility of an effect.
Positive posterior probabilities (*P*+) are shown next to each distribution and color coded. *P*+ values ≥0.90 indicate moderate to very high credibility for a positive effect, P+ ≤0.10 indicate moderate to very high credibility for a negative effect. The smaller P+ values represent the posterior probabilities when adjusting for medication status. The meaning of the direction of effects are shown next to the red zero-effect line. See supplementary Table 1 for the definition of the abbreviated regions of interest. All analyses were adjusted for age and sex.

### Difference in pre-to-post treatment changes between responders and non-responders to I-CBT

Contrast: FEAR

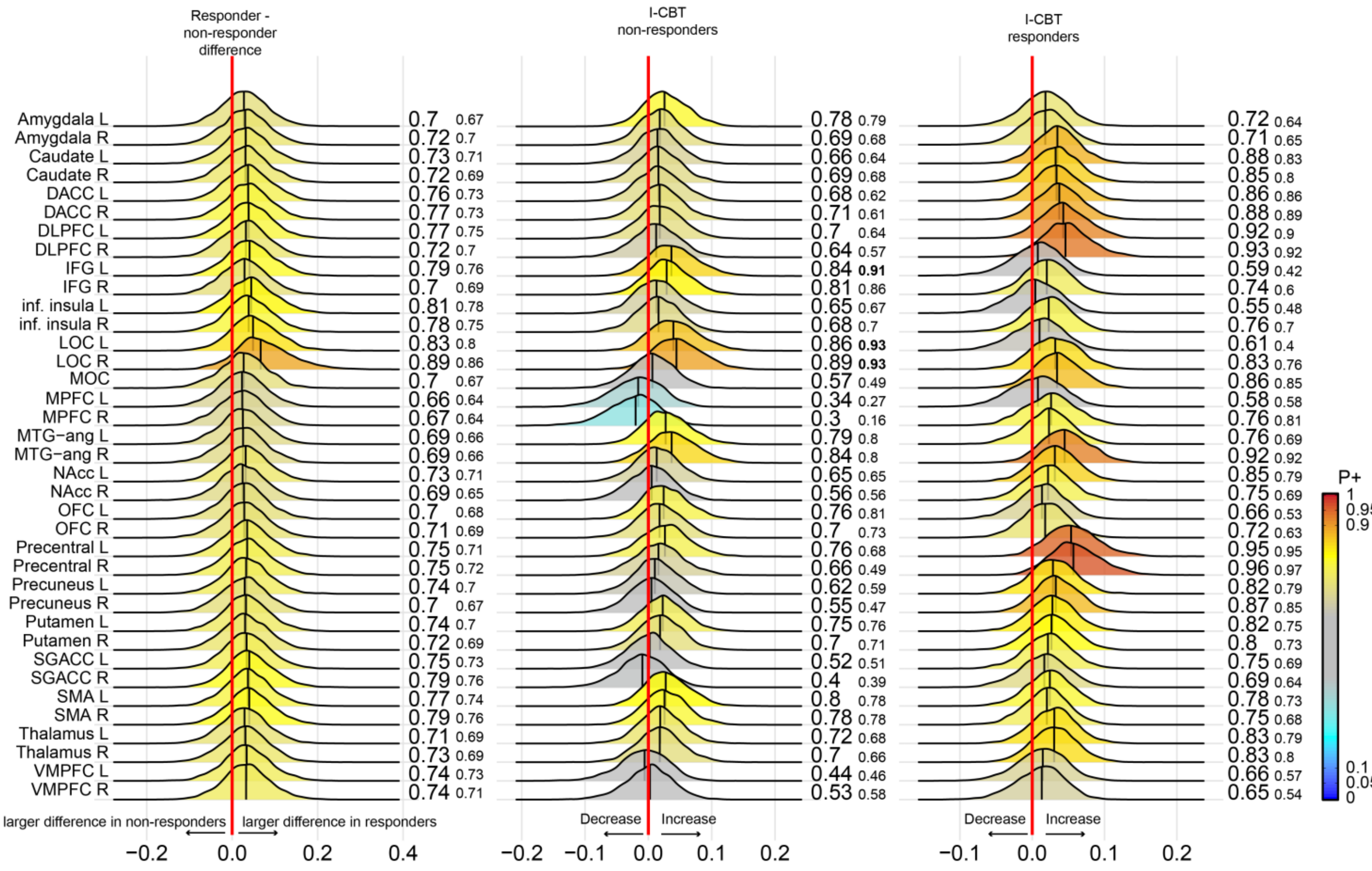

**Supplementary Figure 8 – Bayesian posterior distribution plots of the pre-to-post treatment differences in task activation during the fear contrast between responders and non-responders to I-CBT.**
The first column shows the relative differences between responders and non-responders to I-CBT in the changes in task related activation after treatment. Column two and three show the changes from pre-to-post treatment in the non-responders and responders, respectively. The posterior distribution communicates the credibility of an effect.
Positive posterior probabilities (*P+*) are shown next to each distribution and color coded. *P+* values ≥0.90 indicate moderate to very high credibility for a positive effect, P+ ≤0.10 indicate moderate to very high credibility for a negative effect. The smaller P+ values represent the posterior probabilities when adjusting for medication status. The meaning of the direction of effects are shown next to the red zero-effect line. See supplementary Table 1 for the definition of the abbreviated regions of interest. All analyses were adjusted for age and sex.

#### Whole-brain analyses

| Supplementary Table 7 - CBT-ERP vs. I-CBT comparison in the fear contrast. | | | | | | | |
|---|---|---|---|---|---|---|---|
| **Group, effect** | **Region** | **Hemisphere** | **cluster size (k)** | **MNI x** | **y** | **z** | **Peak Z** |
| **All, time - increase** | | | | | | | |
| | Supramarginal gyrus | L | 9 | -68 | -26 | 32 | 3.54 |
| **All, time - decrease** | | | | | | | |
| | OFC | R | 221 | 26 | 16 | -16 | 5.77 |
| | DLPFC | L | 131 | -20 | 58 | 36 | 4.04 |
| | DLPFC | L | 66 | -14 | 40 | 46 | 3.69 |
| | DLPFC | R | 100 | 18 | 54 | 38 | 3.89 |
| | Pre-SMA | R | 13 | 6 | 18 | 66 | 3.47 |
| | Angular gyrus | L | 60 | -50 | -68 | 44 | 3.55 |
| | Posterior cingulate cortex | L | 7 | -12 | -44 | 32 | 3.18 |
| | MTG | L | 64 | -60 | -14 | -22 | 3.72 |
| | Parahippocampal gyrus / WM | L | 15 | -20 | -30 | -16 | 3.46 |
| | Temporal pole | L | 98 | -44 | 8 | -30 | 3.79 |
| | Temporal pole | R | 81 | 38 | 12 | -36 | 4.27 |
| | Anterior insula | L | 75 | -26 | 6 | -20 | 4.24 |
| | Amygdala | R | 30 | 16 | -4 | -16 | 3.97 |
| | Cerebellum crus II | R | 64 | 30 | -76 | -36 | 3.42 |
| **All, group*time interaction** | | | | | | | |
| | Anterior insula / OFC | L | 8 | -34 | 26 | -4 | 3.17 |
| | dACC | L | 46 | -8 | 30 | 30 | 3.57 |
| | dACC | R | 12 | 8 | 24 | 28 | 3.32 |
| | Pre-SMA | L | 58 | -12 | 2 | 76 | 3.56 |
| | Pre-SMA | L | 8 | -10 | 20 | 56 | 3.6 |
| | Medial cingulate cortex | B | 71 | 6 | 12 | 38 | 3.38 |
| | Precentral gyrus / WM | L | 36 | -32 | -18 | 64 | 4.17 |
| | Postcentral gyrus | L | 12 | -56 | -18 | 34 | 3.41 |
| | Postcentral gyrus | L | 39 | -36 | -26 | 20 | 3.9 |
| | Supramarginal gyrus | L | 11 | -50 | -26 | 16 | 3.37 |
| | Supramarginal gyrus | L | 9 | -60 | -34 | 28 | 3.29 |
| | Angular gyrus | R | 145 | 52 | -46 | 16 | 4.03 |
| | Angular gyrus | R | 9 | 40 | -74 | 36 | 3.39 |
| | Angular gyrus | R | 48 | 52 | -60 | 16 | 3.53 |
| | Precuneus | L | 11 | -14 | -48 | 60 | 3.25 |
| | Posterior cingulate cortex | R | 47 | 10 | -42 | 10 | 4.06 |
| | Posterior cingulate cortex | L | 47 | -8 | -20 | 34 | 4 |
| | Transverse temporal gyrus | R | 14 | 42 | -20 | 14 | 3.56 |
| | Paracentral lobule | R | 11 | 14 | -30 | 48 | 3.31 |
| | Cerebellum lobule VI | R | 11 | 46 | -68 | -26 | 3.37 |
| | Cerebellum lobule VI | L | 20 | -44 | -62 | -28 | 3.36 |
| | Cerebellum lobule VI/crus I | L | 175 | -22 | -72 | -32 | 4.43 |
| | Cerebellum crus I | R | 111 | 50 | -52 | -38 | 3.67 |
| | Cerebellum crus II | L | 16 | -4 | -78 | -30 | 3.3 |
| **CBT-ERP, increase** | | | | | | | |
| | - | | | | | | |

Supplementary Table 7 - CBT-ERP vs. I-CBT comparison in the fear contrast. (continued)

| Group, effect | Region | Hemisphere | cluster size (k) | MNI X | Y | Peak Z z |
|---|---|---|---|---|---|---|
| **CBT-ERP, decrease** | | | | | | |
| | OFC / amygdala | R | 710 | 26 | | |
| | IFG | L | 9 | -46 | | |
| | IFG | R | 17 | 50 | | |
| | IFG | R | 8 | 50 | | |
| | DMPFC | L | 26 | -10 | | |
| | DMPFC | L | 157 | -22 | | |
| | DMPFC | R | 12 | 18 | | |
| | dACC | B | 499 | -8 | | |
| | Pre-SMA | B | 326 | 2 | | |
| | Angular gyrus | L | 44 | -46 | | |
| | Angular gyrus | R | 210 | 56 | | |
| | Cuneus | R | 58 | 10 | | |
| | Posterior cingulate cortex | L | 94 | -10 | | |
| | Posterior cingulate cortex | L | 45 | -6 | | |
| | Posterior cingulate cortex | L | 69 | -4 | | |
| | Posterior cingulate cortex | R | 29 | 10 | | |
| | STG | R | 20 | 48 | | |
| | ITG | R | 18 | 40 | | |
| | Parahippocampal gyrus | L | 10 | -22 | | |
| | Anterior insula | L | 389 | -26 | | |
| | Cerebellum lobule VI | L | 53 | -44 | | |
| | Cerebellum crus I | L | 73 | -22 | | |
| **I-CBT, increase** | | | | | | |
| | Pre-SMA | L | 24 | -28 | | |
| | Precentral gyrus | L | 70 | -32 | | |
| | Precentral gyrus | L | 26 | -46 | | |
| | Postcentral gyrus | L | 107 | -36 | | |
| | Postcentral gyrus | L | 7 | -38 | | |
| | Postcentral gyrus | L | 9 | -8 | | |
| | Supramarginal gyrus | L | 12 | -44 | | |
| | Supramarginal gyrus | L | 296 | -68 | | |
| | Supramarginal gyrus | L | 8 | -52 | | |
| | Supramarginal gyrus | R | 10 | 64 | | |
| | Supramarginal gyrus | R | 14 | 52 | | |
| | Angular gyrus | R | 19 | 40 | | |
| | Angular gyrus / MTG | R | 56 | 52 | | |
| | Medial parietal lobe / WM | R | 19 | 30 | | |
| | Precuneus | L | 52 | -14 | | |
| | Precuneus | R | 5 | 8 | | |
| | Posterior cingulate cortex | L | 33 | -10 | | |
| | Superior occipital gyrus | L | 11 | -38 | | |
| | Globus pallidus | L | 8 | -14 | | |

| Supplementary Table 7 - CBT-ERP vs. I-CBT comparison in the fear contrast. (continued) | | | | | | | |
|---|---|---|---|---|---|---|---|
| **Group, effect** | **Region** | **Hemisphere** | **cluster size (k)** | **MNI X** | **Y** | **z** | **Peak Z** |
| **I-CBT, decrease** | | | | | | | |
| | MTG | L | 66 | -54 | | | |
| | Temporal pole | R | 63 | 38 | | | |

Results are reported at P<0.001, uncorrected for multiple comparisons.
All group*time interaction effects signify a decrease in the CBT-ERP and increase in I-CBT. Abbreviations: OFC = orbitofrontal cortex, DLPFC = dorsolateral prefrontal cortex, pre-SMA = pre-supplementary motor area, MTG = middle temporal gyrus, dACC = dorsal anterior cingulate cortex, IFG = inferior frontal gyrus, DMPFC = dorsomedial prefrontal cortex

**Supplementary Table 8 - Associations between symptom reduction and change in activation in the fear contrast**

| **Group, association** | **Region** | **Hemisphere** | **Cluster size (k)** | **MNI x** | **y** | **z** | **Peak Z** |
|---|---|---|---|---|---|---|---|
| **CBT-ERP, positive** | - | | | | | | |
| **CBT-ERP, negative** | | | | | | | |
| | DLPFC | R | 14 | 38 | 30 | 40 | 3.4 |
| | IFG | L | 18 | -36 | 10 | 14 | 3.64 |
| | SMA | L | 42 | -22 | -6 | 52 | 3.55 |
| | SMA | R | 10 | 16 | 4 | 68 | 3.4 |
| | SMA | R | 9 | 32 | -10 | 54 | 3.37 |
| | Supramarginal / postcentral gyrus | L | 7 | -38 | -28 | 36 | 3.36 |
| | Cerebellum lobule V | R | 10 | 8 | -82 | -36 | 3.37 |
| | Cerebellum lobule VI | L | 8 | -20 | -72 | -48 | 3.58 |
| | Cerebellum lobule IX | R | 10 | 12 | -50 | -46 | 3.26 |
| **I-CBT, positive** | | | | | | | |
| | Anterior PFC | R | 36 | 4 | 66 | 0 | 3.47 |
| | Medial PFC / WM | R | 24 | 26 | 42 | 14 | 3.48 |
| | DMPFC | R | 30 | 20 | 30 | 40 | 3.61 |
| | Precentral gyrus | L | 20 | -32 | -22 | 62 | 3.48 |
| | Precentral gyrus | R | 58 | 36 | -20 | 58 | 4.05 |
| | Precentral gyrus | R | 5 | 68 | -4 | 18 | 3.14 |
| | Postcentral gyrus | L | 21 | -42 | -20 | 44 | 3.65 |
| | Postcentral gyrus | L | 16 | -30 | -34 | 52 | 3.62 |
| | Postcentral gyrus | L | 19 | -20 | -34 | 74 | 3.23 |
| | Postcentral gyrus / superior parietal lobule | R | 5 | 22 | -38 | 56 | 3.22 |
| | Superior parietal lobule | R | 66 | 22 | -54 | 64 | 4.21 |
| | Supramarginal gyrus | L | 40 | -42 | -32 | 22 | 3.6 |
| | Angular gyrus | R | 36 | 58 | -64 | 32 | 3.47 |
| | Precuneus | L | 77 | -8 | -66 | 22 | 3.73 |
| | Precuneus | L | 8 | -14 | -62 | 34 | 3.43 |
| | Lingual gyrus | | 50 | 4 | -76 | -8 | 3.32 |
| | Fusiform gyrus | R | 6 | 18 | -46 | -14 | 3.5 |
| | Hippocampus/WM | L | 18 | -40 | -32 | -8 | 4.26 |
| | STG | L | 32 | 68 | -16 | 10 | 3.54 |
| | STG | L | 5 | -68 | -12 | -2 | 3.37 |
| | Temporal pole | R | 23 | 64 | 4 | -4 | 3.31 |
| **I-CBT, negative** | | | | | | | |
| | OFC | L | 73 | -38 | 26 | -2 | 3.92 |
| | DLPFC | L | 15 | -52 | 32 | 30 | 3.34 |
| | Parahippocampal gyrus | L | 11 | -32 | -6 | -34 | 3.6 |

Results are reported at P<0.001, uncorrected for multiple comparisons. Abbreviations: OFC = orbitofrontal cortex, DLPFC = dorsolateral prefrontal cortex, pre-SMA = pre-supplementary motor area, SMA = supplementary motor area, STG = superior temporal gyrus, IFG = inferior frontal gyrus, DMPFC = dorsomedial prefrontal cortex, PFC = prefrontal cortex, WM = white matter.

### Difference in pre-to-post treatment changes between responders and non-responders to CBT-ERP

Contrast: OCD

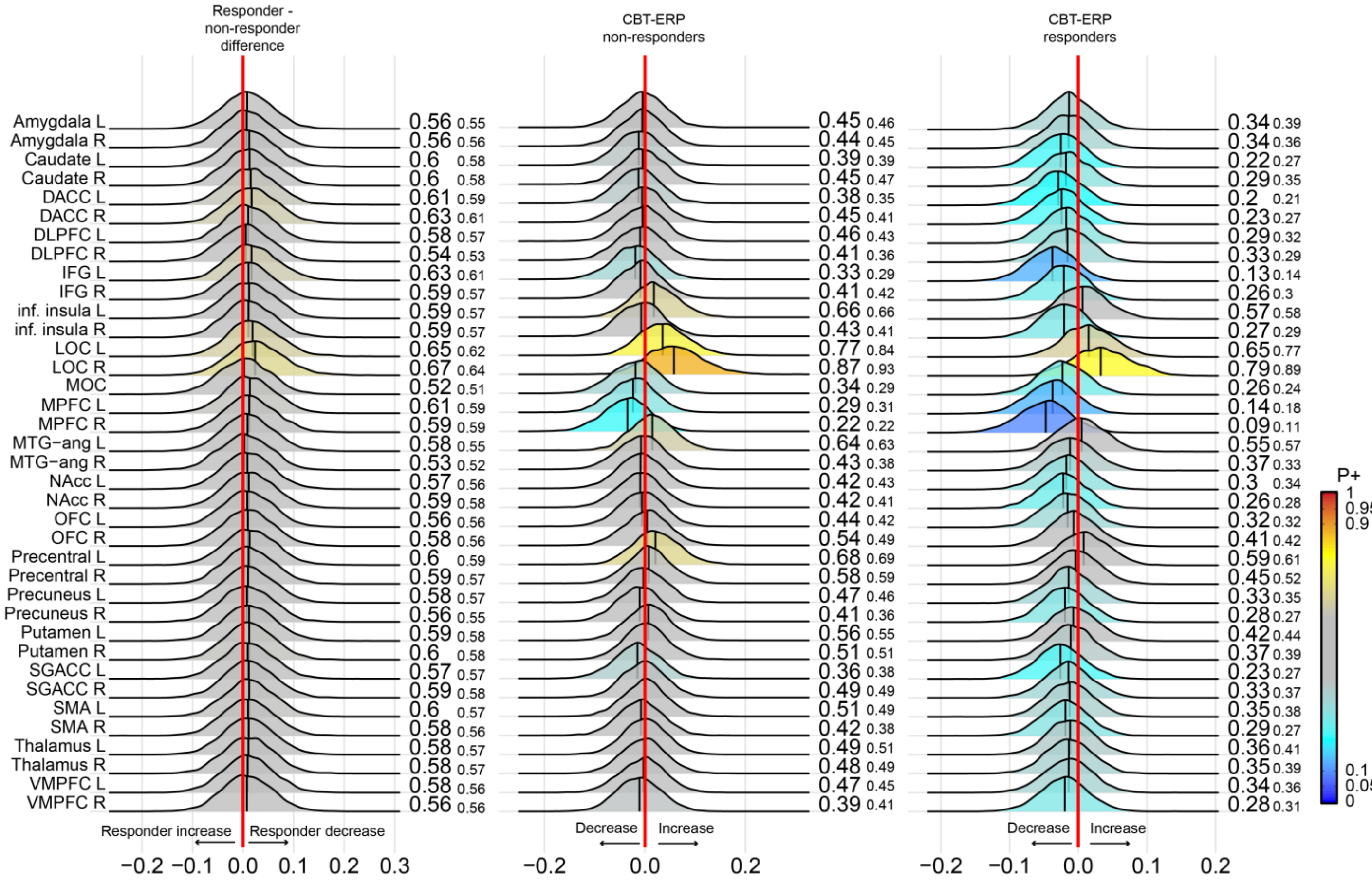

**Supplementary Figure 9 – Bayesian posterior distribution plots of the pre-to-post treatment differences in task activation during the OCD contrast between responders and non-responders to CBT-ERP.**
The first column shows the relative differences between responders and non-responders to CBT-ERP in the changes in task related activation after treatment. Column two and three show the changes from pre-to-post treatment in the non-responders and responders, respectively. The posterior distribution communicates the credibility of an effect.
Positive posterior probabilities (*P*+) are shown next to each distribution and color coded. *P*+ values ≥0.90 indicate moderate to very high credibility for a positive effect, P+ ≤0.10 indicate moderate to very high credibility for a negative effect. The smaller P+ values represent the posterior probabilities when adjusting for medication status. The meaning of the direction of effects are shown next to the red zero-effect line. See supplementary Table 1 for the definition of the abbreviated regions of interest. All analyses were adjusted for age and sex.

### Difference in pre-to-post treatment changes between responders and non-responders to I-CBT

Contrast: OCD

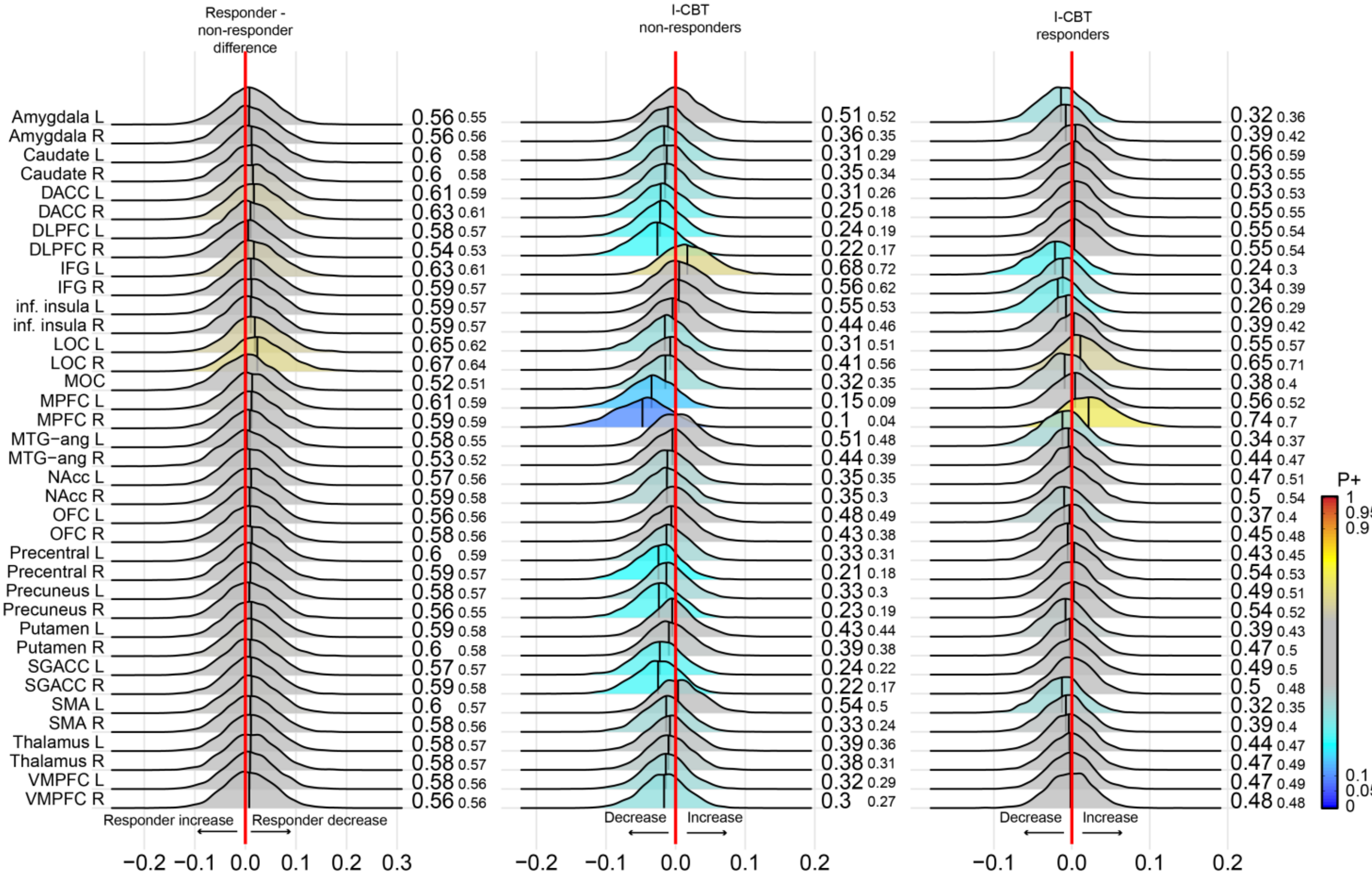

**Supplementary Figure 10 – Bayesian posterior distribution plots of the pre-to-post treatment differences in task activation during the OCD contrast between responders and non-responders to I-CBT.**

The first column shows the relative differences between responders and non-responders to I-CBT in the changes in task related activation after treatment. Column two and three show the changes from pre-to-post treatment in the non-responders and responders, respectively. The posterior distribution communicates the credibility of an effect.

Positive posterior probabilities (*P*+) are shown next to each distribution and color coded. *P*+ values ≥0.90 indicate moderate to very high credibility for a positive effect, P+ ≤0.10 indicate moderate to very high credibility for a negative effect. The smaller P+ values represent the posterior probabilities when adjusting for medication status. The meaning of the direction of effects are shown next to the red zero-effect line. See supplementary Table 1 for the definition of the abbreviated regions of interest. All analyses were adjusted for age and sex.

### Whole-brain analyses

**Supplementary Table 9 - CBT-ERP vs. I-CBT comparison in the OCD contrast**

| Group, effect | Region | Hemisphere | cluster size (k) | MNI X | Y | Z | Peak Z |
|---|---|---|---|---|---|---|---|
| **All, time - increase** | | | | | | | |
| | - | | | | | | |
| **All, time - decrease** | | | | | | | |
| | Angular gyrus | L | 113 | -50 | -70 | 40 | 4.18 |
| | Amygdala | R | 27 | 16 | -4 | -16 | 3.92 |
| | Precuneus | L | 89 | -10 | -58 | 34 | 3.82 |
| | Posterior cingulate cortex | R | 32 | 10 | -52 | 22 | 3.68 |
| | DLPFC | L | 40 | -22 | 54 | 38 | 3.67 |
| | OFC | R | 17 | 26 | 18 | -14 | 3.55 |
| | Angular gyrus | R | 61 | 54 | -62 | 26 | 3.55 |
| | DMPFC | L | 15 | -4 | 34 | 34 | 3.4 |
| | DLPFC | L | 8 | -28 | 60 | 26 | 3.26 |
| | STG | R | 6 | 50 | 16 | -10 | 3.2 |
| | Posterior cingulate cortex | L | 7 | -4 | -44 | 30 | 3.18 |
| **All, group*time interaction** | | | | | | | |
| | - | | | | | | |
| **CBT-ERP, increase** | | | | | | | |
| | Posterior insula | L | 25 | -42 | -10 | 10 | 3.65 |
| **CBT-ERP, decrease** | | | | | | | |
| | DMPFC | L | 28 | -4 | 34 | 34 | 3.47 |
| | DMPFC | R | 13 | 16 | 28 | 48 | 3.55 |
| | Angular gyrus | L | 5 | -46 | -66 | 22 | 3.24 |
| | Angular gyrus | R | 61 | 54 | -62 | 24 | 3.71 |
| | Precuneus | L | 20 | -12 | -58 | 36 | 3.58 |
| | Posterior cingulate cortex | R | 10 | 10 | -50 | 20 | 3.41 |
| | Amygdala | R | 7 | 16 | -2 | -14 | 3.45 |
| **I-CBT, increase** | | | | | | | |
| | Medial parietal lobe | L | 30 | -30 | -66 | 24 | 3.46 |
| | Medial parietal lobe | R | 7 | 28 | -56 | 34 | 3.35 |
| **I-CBT, decrease** | | | | | | | |
| | DLPFC | R | 8 | 20 | 54 | 36 | 3.3 |
| | IFG | L | 35 | -30 | 24 | 14 | 3.75 |
| | IFG | R | 20 | 32 | 24 | 14 | 3.78 |
| | Precentral gyrus | L | 14 | -46 | -4 | 18 | 3.66 |
| | Angular gyrus | L | 63 | -54 | -68 | 38 | 3.78 |
| | Parahippocampal gyrus | L | 15 | -16 | -34 | -10 | 3.62 |
| | STG / MTG | R | 13 | 50 | -28 | -2 | 3.28 |
| | STG | L | 5 | -66 | -6 | -14 | 3.35 |
| | STG / WM | L | 60 | -48 | -20 | -6 | 3.94 |
| | MTG | L | 14 | -52 | -10 | -24 | 3.47 |
| | ITG | R | 11 | 46 | -2 | -34 | 3.3 |
| | Temporal pole | R | 6 | 40 | 24 | -28 | 3.55 |

Results are reported at P<0.001, uncorrected for multiple comparisons. All group*time interaction effects signify a decrease in the CBT-ERP and increase in I-CBT. Abbreviations: OFC = orbitofrontal cortex, DLPFC = dorsolateral prefrontal cortex, STG = superior temporal gyrus, ITG = inferior temporal gyrus, IFG = inferior frontal gyrus, DMPFC = dorsomedial prefrontal cortex, PFC = prefrontal cortex, WM = white matter.

### Whole-brain analyses

**Supplementary Table 10 - Associations between symptom reduction and change in activation in the OCD contrast**

| **Group, association** | Region | Hemisphere | Cluster size (k) | MNI x | y | z | Peak Z |
|---|---|---|---|---|---|---|---|
| **CBT-ERP, positive** | - | | | | | | |
| **CBT-ERP, negative** | | | | | | | |
| | SMA | | 7 | -20 | -2 | 54 | 3.29 |
| **I-CBT, positive** | | | | | | | |
| | DLPFC | R | 5 | 36 | 24 | 36 | 3.31 |
| | Postcentral gyrus / WM | R | 34 | 24 | -44 | 54 | 3.53 |
| | STG | R | 16 | 58 | 2 | -4 | 3.61 |
| | WM | | 6 | -12 | -32 | 52 | 3.72 |
| **I-CBT, negative** | | | | | | | |
| | DLPFC | R | 14 | 38 | 38 | 10 | 3.71 |
| | Angular gyrus | L | 12 | -54 | -56 | 52 | 3.65 |
| | Parahippocampal gyrus | L | 8 | -30 | -20 | -28 | 3.48 |
| | Parahippocampal gyrus | L | 8 | -38 | -12 | -34 | 3.31 |

Results are reported at P<0.001, uncorrected for multiple comparisons. Abbreviations: SMA = supplementary motor area, DLPFC = dorsolateral prefrontal cortex, WM = white matter, STG = superior temporal gyrus.

### EMOTIONAL CONTRAST

The positive posterior distribution plots in supplementary Figure 11 show evidence for opposite pre-to-post treatment changes in task-related activation for the emotional contrast in the CBT-ERP compared to the I-CBT group. There was moderate evidence for decreased activation after CBT-ERP in the left (P+ = 0.07) and right (P+ = 0.09) middle prefrontal cortex (mPFC) and middle occipital cortex (MOC; P+ = 0.10) and strong evidence for increased activation in the right LOC (P+ = 0.95) after I-CBT. Directly comparing the two treatment groups showed evidence for a relatively larger decrease in activation in the CBT-ERP group across all regions of interest, but none with P+ < 0.10. Adding medication status as an additional covariate had little effect on these results, except that both treatment groups showed credible evidence for a treatment-induced increase in LOC (right more than left) activity.

There was no credible evidence for an association between change in OCD symptom severity and change in task-related brain activation when considering both treatment groups together. When considering the two treatment groups separately, the CBT-ERP group showed no credible evidence for associations between change in symptoms and brain activation. Conversely, the I-CBT group showed credible evidence for positive associations, particularly in the left (P+ = 0.99) and right precentral gyrus (P+ = 1.00), bilateral mPFC (P+ = 1.00) , MOC (P+ = 0.98), right precuneus (P+ = 0.95), right dlPFC (P+ = 0.93) and left VMPFC (P+ = 0.9) (supplementary Figure 12). Results were comparable when adjusting for medication status. There was no credible evidence for a change in activity in CBT-ERP responders (compared with non-responders) (supplementary figure 13). Conversely, I-CBT responders showed an increase in activity in the bilateral precentral gyrus (P+ = 0.92), bilateral dorsal ACC (P+ = 0.90-0.92), right DLPFC (P+ = 0.92) and right MPFC (P+ = 0.95) (supplementary figures 14). Non-responders showed evidence for a decrease in right MPFC activity but only after adjusting for medication status (P+ = 0.06).

### Pre-to-post treatment changes

Contrast: EMOTION

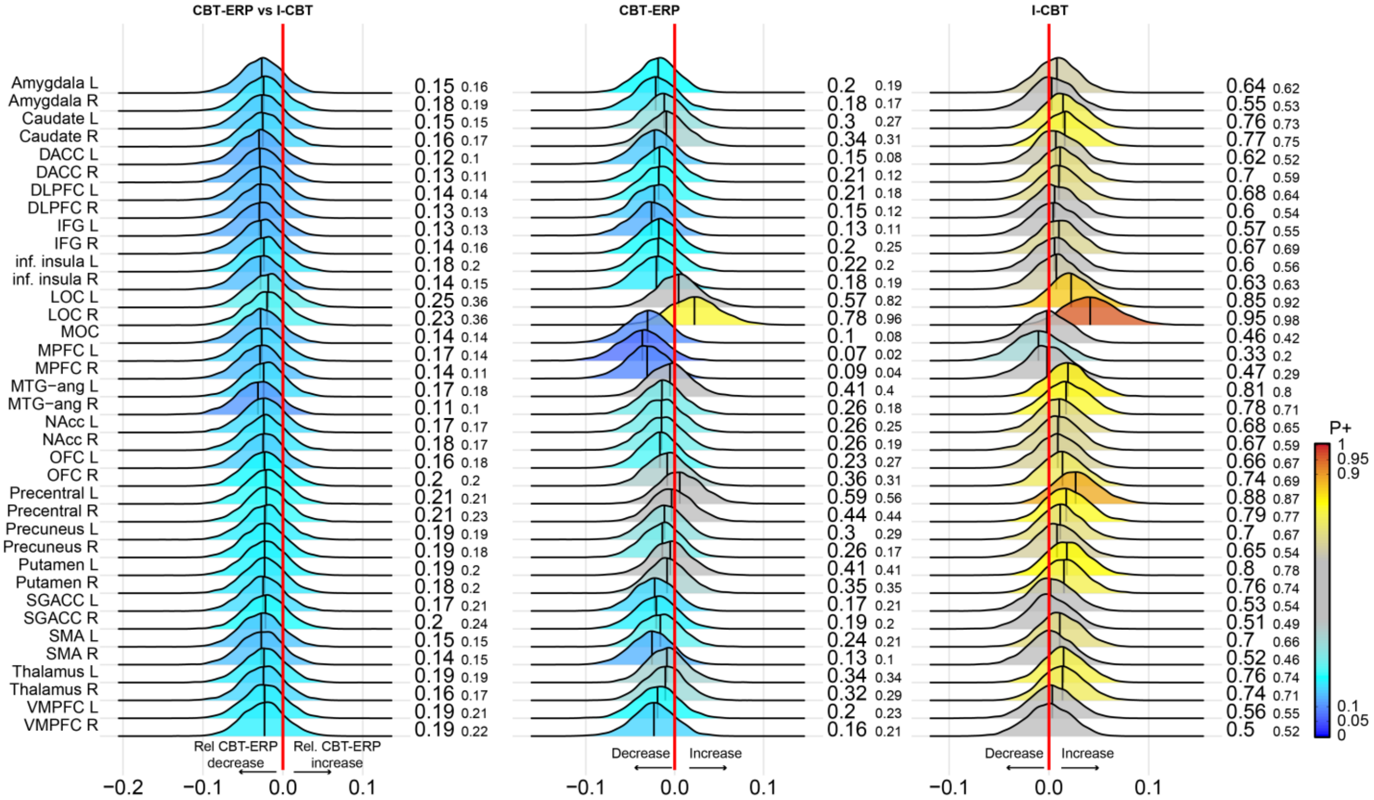

**Supplementary Figure 11 – Bayesian posterior distribution plots of the pre-to-post treatment differences in task activation during the emotion contrast.**

The first column shows the relative differences the CBT-ERP group and I-CBT show in the changes in task related activation after treatment. Column two and three show the changes from pre-to-post treatment in the CBT-ERP and I-CBT group, respectively. The posterior distribution communicates the credibility of an effect. Positive posterior probabilities (*P*+) are shown next to each distribution and color coded. *P*+ values ≥0.90 indicate moderate to very high credibility for a positive effect, P+ ≤0.10 indicate moderate to very high credibility for a negative effect. The smaller P+ values represent the posterior probabilities when adjusting for medication status. The meaning of the direction of effects are shown next to the red zero-effect line. See supplementary Table 1 for the definition of the abbreviated regions of interest. All analyses were adjusted for age and sex.

### Association between pre-to-post treatment changes in activation and symptoms

Contrast: EMOTION

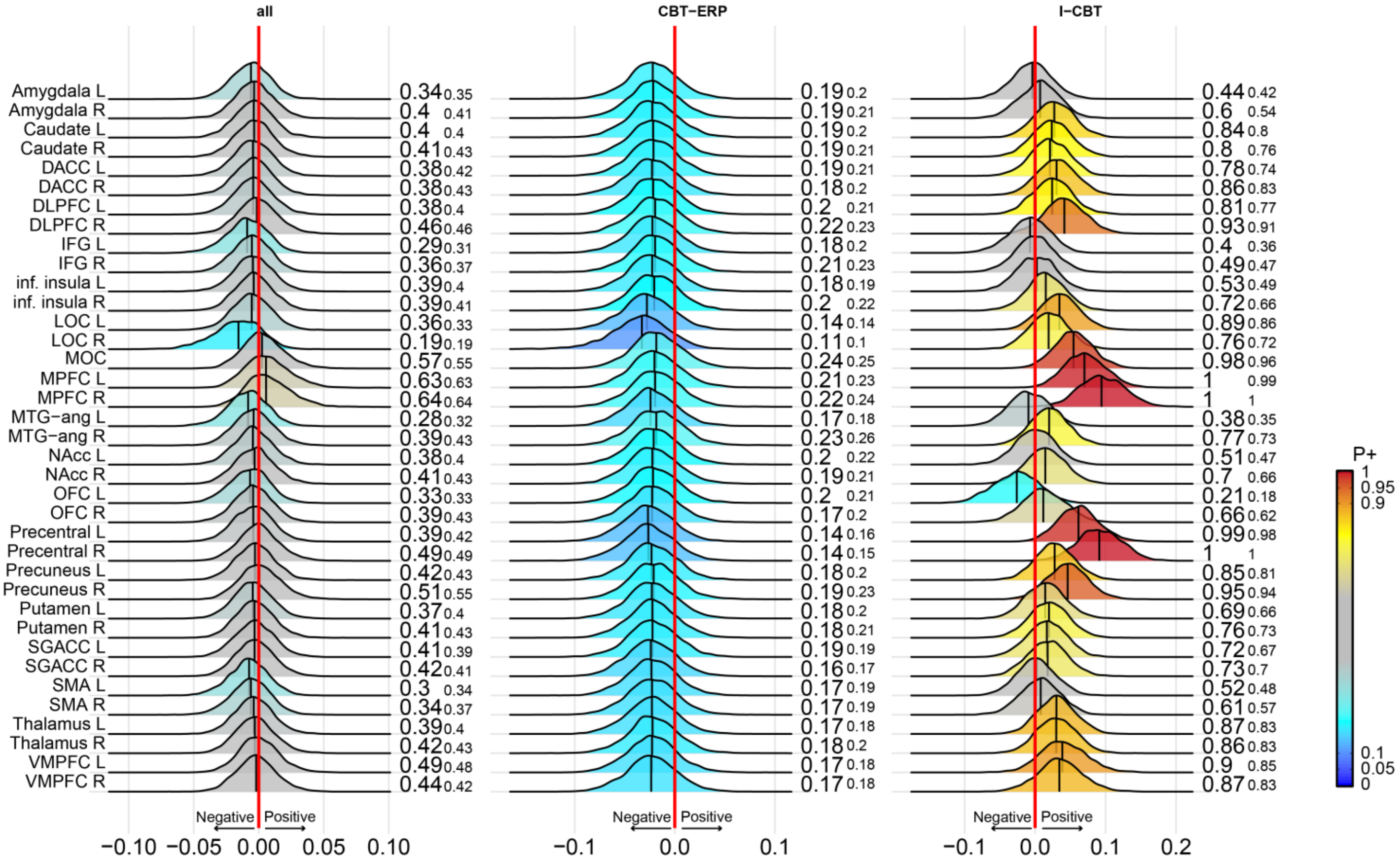

**Supplementary Figure 12 – Bayesian posterior distribution plots of the association between pre-to-post treatment differences in task activation during the emotion contrast and change in symptoms.**
The first column shows the association across both treatment groups. Column two and three show the associations in the CBT-ERP and I-CBT group, respectively. The posterior distribution communicates the credibility of an effect.
Positive posterior probabilities (*P+*) are shown next to each distribution and color coded. *P+* values ≥0.90 indicate moderate to very high credibility for a positive association, P+ ≤0.10 indicate moderate to very high credibility for a negative association. The smaller P+ values represent the posterior probabilities when adjusting for medication status. The meaning of the direction of effects are shown next to the red zero-effect line. See supplementary Table 1 for the definition of the abbreviated regions of interest. All analyses were adjusted for age and sex and baseline YBOCS score.

### Difference in pre-to-post treatment changes between responders and non-responders to CBT-ERP

Contrast: EMOTION

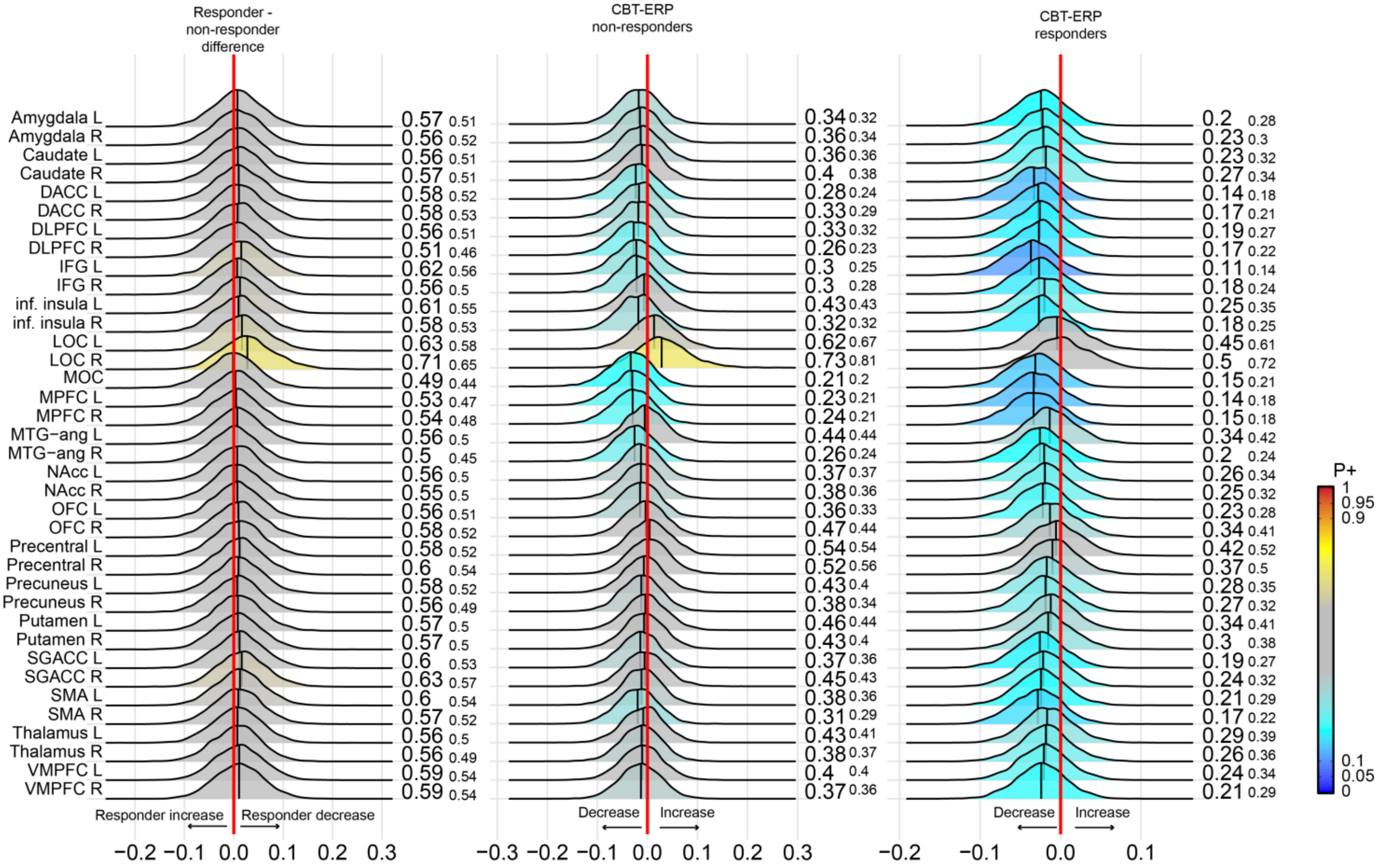

**Supplementary Figure 13 – Bayesian posterior distribution plots of the pre-to-post treatment differences in task activation during the emotion contrast between responders and non-responders to CBT-ERP.**
The first column shows the relative differences between responders and non-responders to CBT-ERP in the changes in task related activation after treatment. Column two and three show the changes from pre-to-post treatment in the non-responders and responders, respectively. The posterior distribution communicates the credibility of an effect.
Positive posterior probabilities (*P+*) are shown next to each distribution and color coded. *P+* values ≥0.90 indicate moderate to very high credibility for a positive effect, P+ ≤0.10 indicate moderate to very high credibility for a negative effect. The smaller P+ values represent the posterior probabilities when adjusting for medication status. The meaning of the direction of effects are shown next to the red zero-effect line. See supplementary Table 1 for the definition of the abbreviated regions of interest. All analyses were adjusted for age and sex.

### Difference in pre-to-post treatment changes between responders and non-responders to I-CBT

Contrast: EMOTION

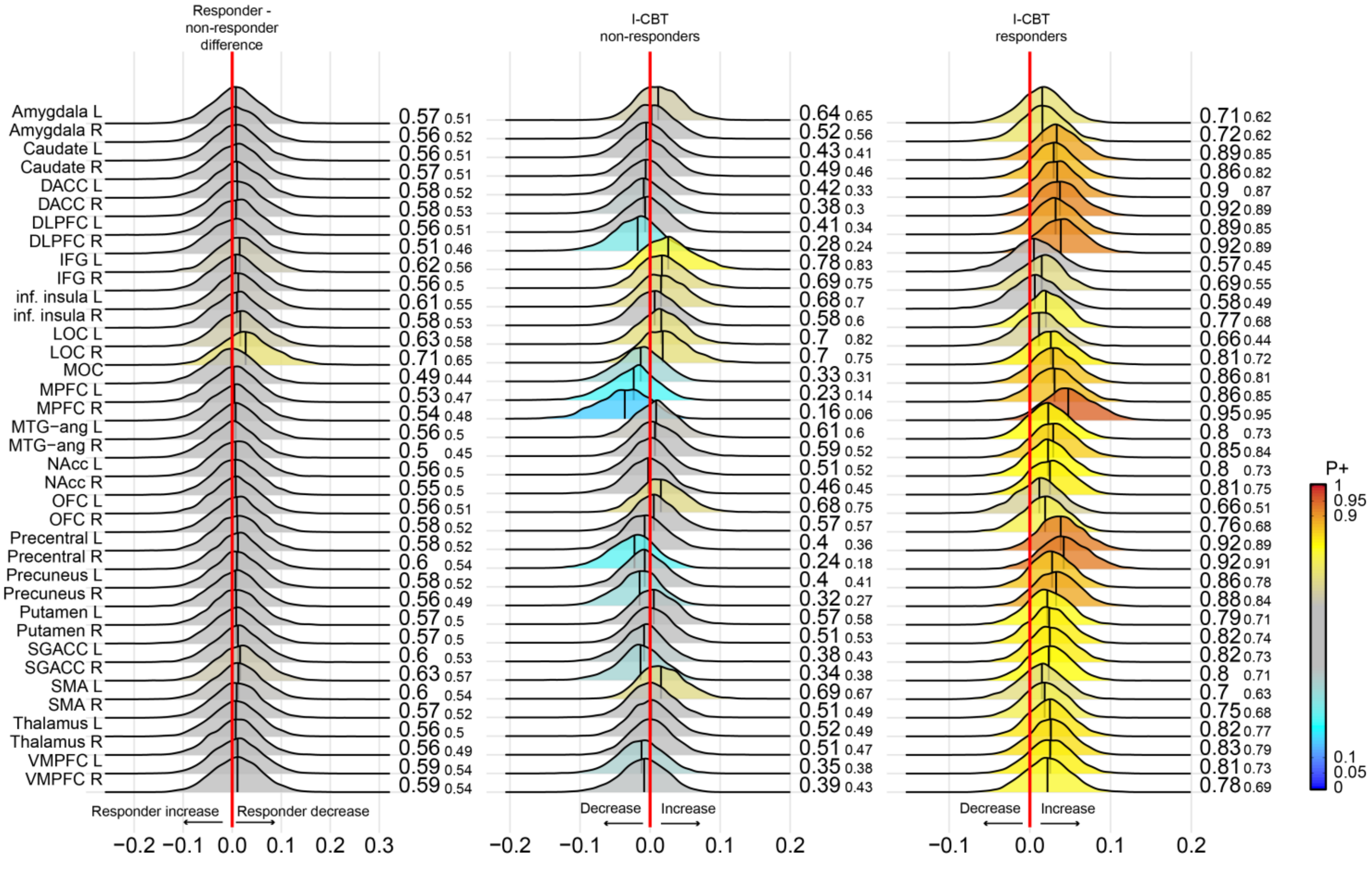

**Supplementary Figure 14 – Bayesian posterior distribution plots of the pre-to-post treatment differences in task activation during the emotion contrast between responders and non-responders to I-CBT.**

The first column shows the relative differences between responders and non-responders to I-CBT in the changes in task related activation after treatment. Column two and three show the changes from pre-to-post treatment in the non-responders and responders, respectively. The posterior distribution communicates the credibility of an effect.

Positive posterior probabilities (*P*+) are shown next to each distribution and color coded. *P*+ values ≥0.90 indicate moderate to very high credibility for a positive effect, P+ ≤0.10 indicate moderate to very high credibility for a negative effect. The smaller P+ values represent the posterior probabilities when adjusting for medication status. The meaning of the direction of effects are shown next to the red zero-effect line. See supplementary Table 1 for the definition of the abbreviated regions of interest. All analyses were adjusted for age and sex.

### Whole-brain analyses

| Supplementary table 11 - CBT-ERP vs. I-CBT comparison in the emotional contrast. | | | | | | | |
|---|---|---|---|---|---|---|---|
| **Group, effect** | Region | Hemisphere | cluster size (k) | MNI X | Y | Z | Peak Z |
| **All, time - increase** | | | | | | | |
| | Postcentral gyrus | R | 14 | 62 | -14 | 30 | 3.33 |
| | Supramarginal gyrus | L | 70 | -68 | -26 | 32 | 4.24 |
| | Supramarginal gyrus | L | 32 | -54 | -28 | 44 | 3.41 |
| | Supramarginal gyrus | L | 9 | -44 | -40 | 40 | 3.32 |
| **All, time - decrease** | | | | | | | |
| | DLPFC | L | 61 | -22 | 56 | 38 | 4.19 |
| | DLPFC | L | 5 | -28 | 58 | 26 | 3.23 |
| | DLPFC | L | 11 | -16 | 38 | 48 | 3.23 |
| | DLPFC | R | 49 | 16 | 58 | 36 | 3.8 |
| | Angular gyrus | L | 61 | -50 | -70 | 42 | 3.9 |
| | Temporal pole | R | 16 | 48 | 16 | -16 | 3.36 |
| | Temporal pole | R | 5 | 38 | 12 | -36 | 3.29 |
| | Temporal pole | R | 7 | 58 | 4 | -28 | 3.22 |
| | Anterior insula | R | 70 | 26 | 16 | -16 | 5 |
| | Amygdala | R | 46 | 16 | -4 | -14 | 4.33 |
| **All, group*time interaction** | | | | | | | |
| | Caudate nucleus | R | 14 | 14 | 24 | 6 | 3.37 |
| **CBT-ERP, increase** | | | | | | | |
| | SMA | R | 37 | 12 | -18 | 60 | 3.89 |
| | Supramarginal gyrus | L | 21 | -52 | -28 | 44 | 3.46 |
| **CBT-ERP, decrease** | | | | | | | |
| | OFC | R | 53 | 26 | 16 | -16 | 4.37 |
| | DMPFC | B | 150 | -6 | 34 | 34 | 3.97 |
| | DLPFC | L | 32 | -22 | 56 | 38 | 3.67 |
| | DLPFC | L | 15 | -28 | 62 | 24 | 3.48 |
| | DLPFC | R | 80 | 22 | 56 | 26 | 4.02 |
| | DLPFC | R | 4 | 18 | 44 | 44 | 3.28 |
| | Angular gyrus | L | 5 | -46 | -68 | 22 | 3.16 |
| | Angular gyrus | R | 44 | 54 | -62 | 24 | 3.47 |
| | Cuneus | R | 6 | 10 | -80 | 30 | 3.17 |
| | Temporal pole | L | 20 | 50 | 18 | -10 | 3.34 |
| | Insula | L | 26 | -34 | 16 | -8 | 3.55 |
| | Amygdala | R | 37 | 16 | -4 | -12 | 3.95 |
| **I-CBT, increase** | | | | | | | |
| | Supramarginal gyrus | L | 117 | -68 | -26 | 32 | 3.84 |
| | Supramarginal gyrus | L | 12 | -44 | -42 | 42 | 3.41 |
| | STG/Supramarginal gyrus | L | 35 | -54 | -28 | 16 | 3.45 |
| | Superior occipital gyrus | L | 51 | -30 | -66 | 24 | 3.71 |
| | Superior occipital gyrus | L | 26 | -38 | -62 | 6 | 3.8 |
| | MTG | L | 24 | -50 | -50 | 0 | 3.55 |
| | Globus pallidus | L | 16 | -14 | 6 | 2 | 3.36 |
| | Caudate nucleus | R | 14 | 16 | 24 | 6 | 3.35 |
| | Caudate nucleus | L | 12 | -16 | 18 | 4 | 3.31 |
| **I-CBT, decrease** | | | | | | | |
| | Angular gyrus | L | 10 | -50 | -70 | 42 | 3.43 |
| | MTG | L | 5 | -54 | -12 | -22 | 3.22 |
| | ITG | R | 6 | 46 | -2 | -36 | 3.48 |
| | Temporal pole | R | 15 | 38 | 10 | -34 | 3.59 |

Results are reported at P<0.001, uncorrected for multiple comparisons. The group*time interaction effect signifies a decrease in CBT-ERP and an increase in I-CBT. Abbreviations: orbitofrontal cortex, DLPFC = dorsolateral prefrontal cortex, MTG = middle temporal gyrus, IFG = inferior frontal gyrus, DMPFC = dorsomedial prefrontal cortex, SMA = supplementary motor area

### Whole-brain analyses

**Supplementary Table 12 - Associations between symptom reduction and change in activation in the emotional contrast**

| Group, association | Region | Hemisphere | Cluster size (k) | MNI x | y | z | Peak Z |
|---|---|---|---|---|---|---|---|
| **CBT-ERP, positive** | - | | | | | | |
| **CBT-ERP, negative** | | | | | | | |
| | sgACC | R | 6 | 4 | 42 | -6 | 3.49 |
| | Cerebellum crus II | R | 6 | 38 | -68 | -44 | 3.2 |
| **I-CBT, positive** | | | | | | | |
| | dACC | R | 24 | 12 | 46 | 16 | 3.3 |
| | DMPFC | R | 10 | 16 | 40 | 36 | 3.43 |
| | DLPFC | L | 19 | -24 | 16 | 42 | 3.4 |
| | DLPFC | R | 18 | 36 | 26 | 34 | 3.67 |
| | Middle frontal gyrus | L | 25 | -58 | -2 | 38 | 3.58 |
| | Precentral gyrus | L | 348 | -40 | -20 | 46 | 4.86 |
| | Precentral gyrus | L | 47 | -32 | -22 | 70 | 3.6 |
| | Precentral gyrus | L | 14 | -6 | -20 | 58 | 3.53 |
| | Precentral gyrus | R | 84 | 36 | -20 | 58 | 3.73 |
| | Precentral gyrus | R | 22 | 66 | 2 | 32 | 3.32 |
| | Postcentral gyrus | R | 6 | 60 | -12 | 40 | 3.26 |
| | Angular gyrus / postcentral gyrus | R | 151 | 24 | -52 | 62 | 4.31 |
| | Supramarginal gyrus | L | 10 | -42 | -32 | 24 | 3.31 |
| | Cuneus | L | 49 | -10 | -88 | 32 | 3.73 |
| | Cuneus | R | 8 | 6 | -86 | 30 | 3.33 |
| | Posterior cingulate cortex | L | 6 | -16 | -56 | 12 | 3.23 |
| | STG | L | 8 | -68 | -14 | -2 | 3.46 |
| | STG | R | 43 | 60 | 2 | -4 | 3.74 |
| | Posterior thalamus | R | 8 | 8 | -22 | 14 | 3.23 |
| **I-CBT, negative** | | | | | | | |
| | DLPFC | R | 5 | 38 | 40 | 8 | 3.43 |
| | Inferior temporal gyrus | L | 19 | -36 | -8 | -36 | 3.66 |

Results are reported at P<0.001, uncorrected for multiple comparisons. Abbreviations: sgACC = subgenual anterior cingulate cortex, dACC = doral anterior cingulate cortex, DMPFC = dorsomedial prefrontal cortex, DLPFC = dorsolateral prefrontal cortex, STG = superior temporal cortex.

### Association between pre-treatment activation and symptom improvement – PP sample (N=65)

Contrast: FEAR

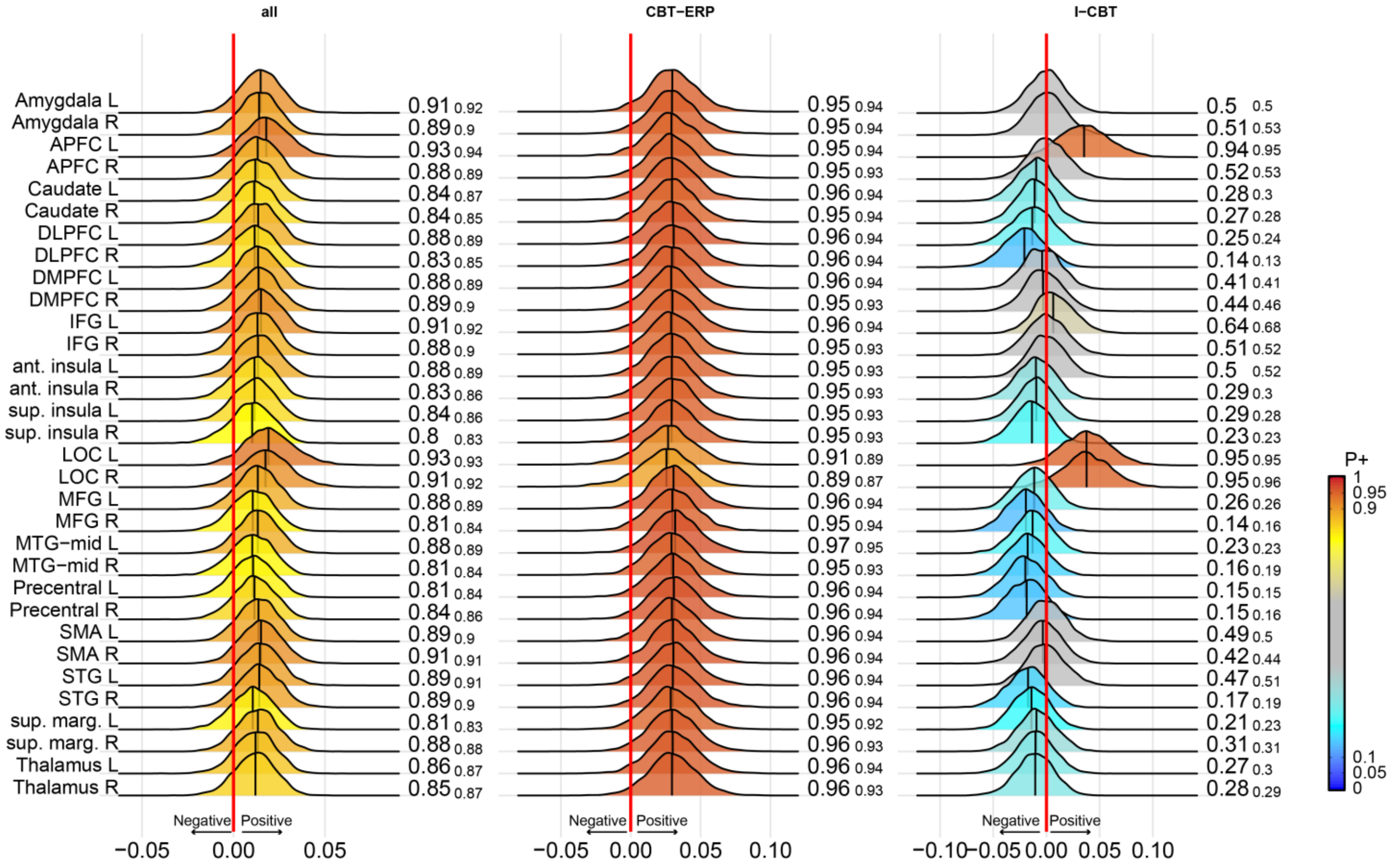

**Supplementary Figure 15 – Bayesian posterior distribution plots of the association between baseline task activation during the fear contrast and change in OCD symptoms after treatment in the Per Protocol (PP) sample.**

The first column shows the association across both treatment groups. Column two and three show the associations in the CBT-ERP and I-CBT group, respectively. The posterior distribution communicates the credibility of an effect. Positive posterior probabilities (*P*+) are shown next to each distribution and color coded. *P*+ values ≥0.90 indicate moderate to very high credibility for a positive effect, P+ ≤0.10 indicate moderate to very high credibility for a negative effect. The smaller P+ values represent the posterior probabilities when adjusting for medication status. The meaning of the direction of effects are shown next to the red zero-effect line. See supplementary Table 1 for the definition of the abbreviated regions of interest. All analyses were adjusted for age and sex and baseline YBOCS score.

### Difference in pre-treatment activation between responders and non-responders – ITT sample (N=73)

Contrast: FEAR

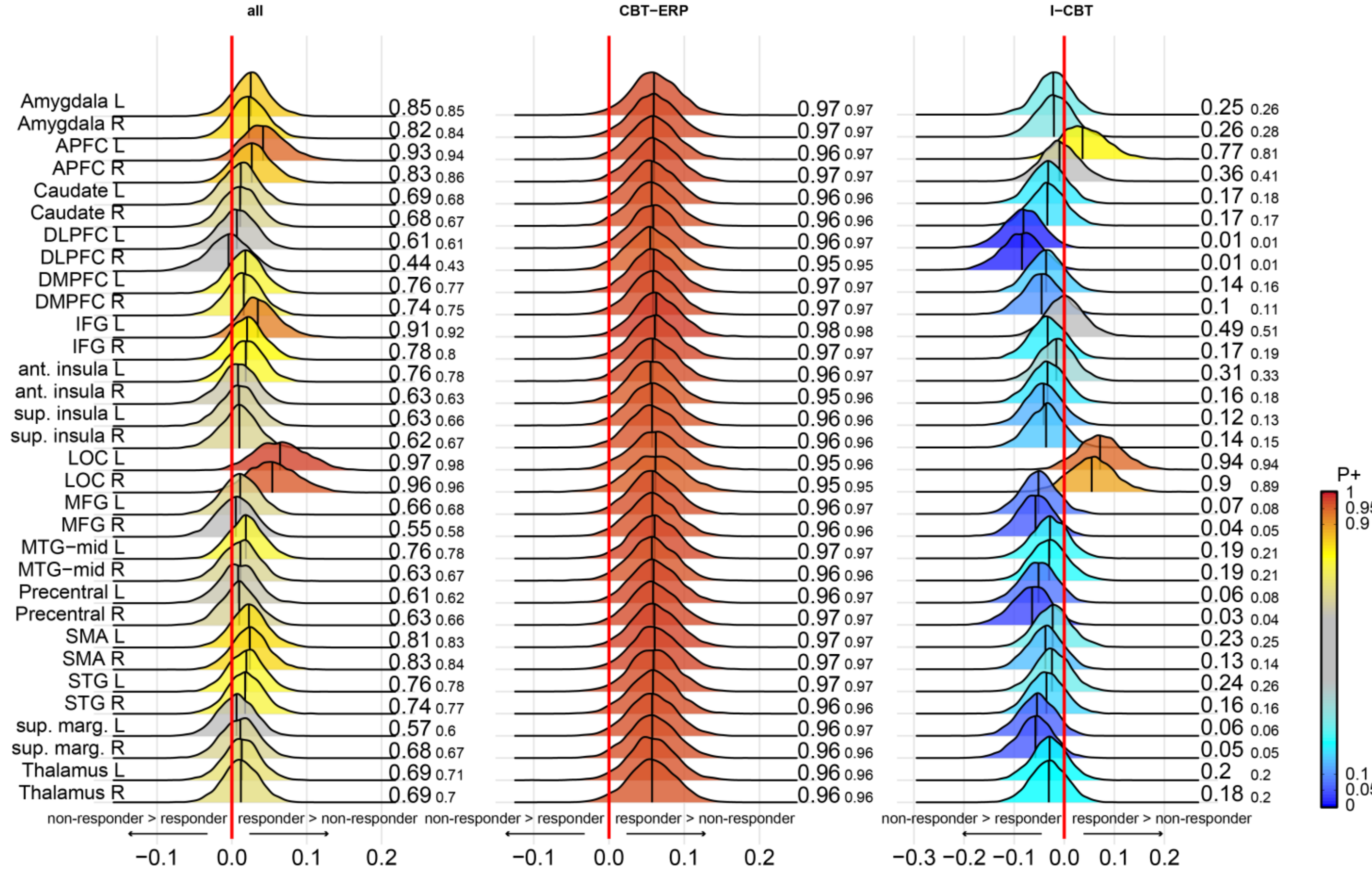

**Supplementary Figure 16 – Bayesian posterior distribution plots of the difference in baseline task activation during the fear contrast between responders and non-responders in the intention-to-treat (ITT) sample.** The first column shows the difference between responders and non-responders across both treatment groups. Column two and three show the between-group difference in the CBT-ERP and I-CBT treatment conditions, respectively. The posterior distribution communicates the credibility of an effect. Positive posterior probabilities (*P*+) are shown next to each distribution and color coded. *P*+ values ≥0.90 indicate moderate to very high credibility for a positive effect, P+ ≤0.10 indicate moderate to very high credibility for a negative effect. The smaller P+ values represent the posterior probabilities when adjusting for medication status. The meaning of the direction of effects are shown next to the red zero-effect line. See supplementary Table 1 for the definition of the abbreviated regions of interest. All analyses were adjusted for age and sex.

### Difference in pre-treatment activation between responders and non-responders – PP sample (N=65)

Contrast: FEAR

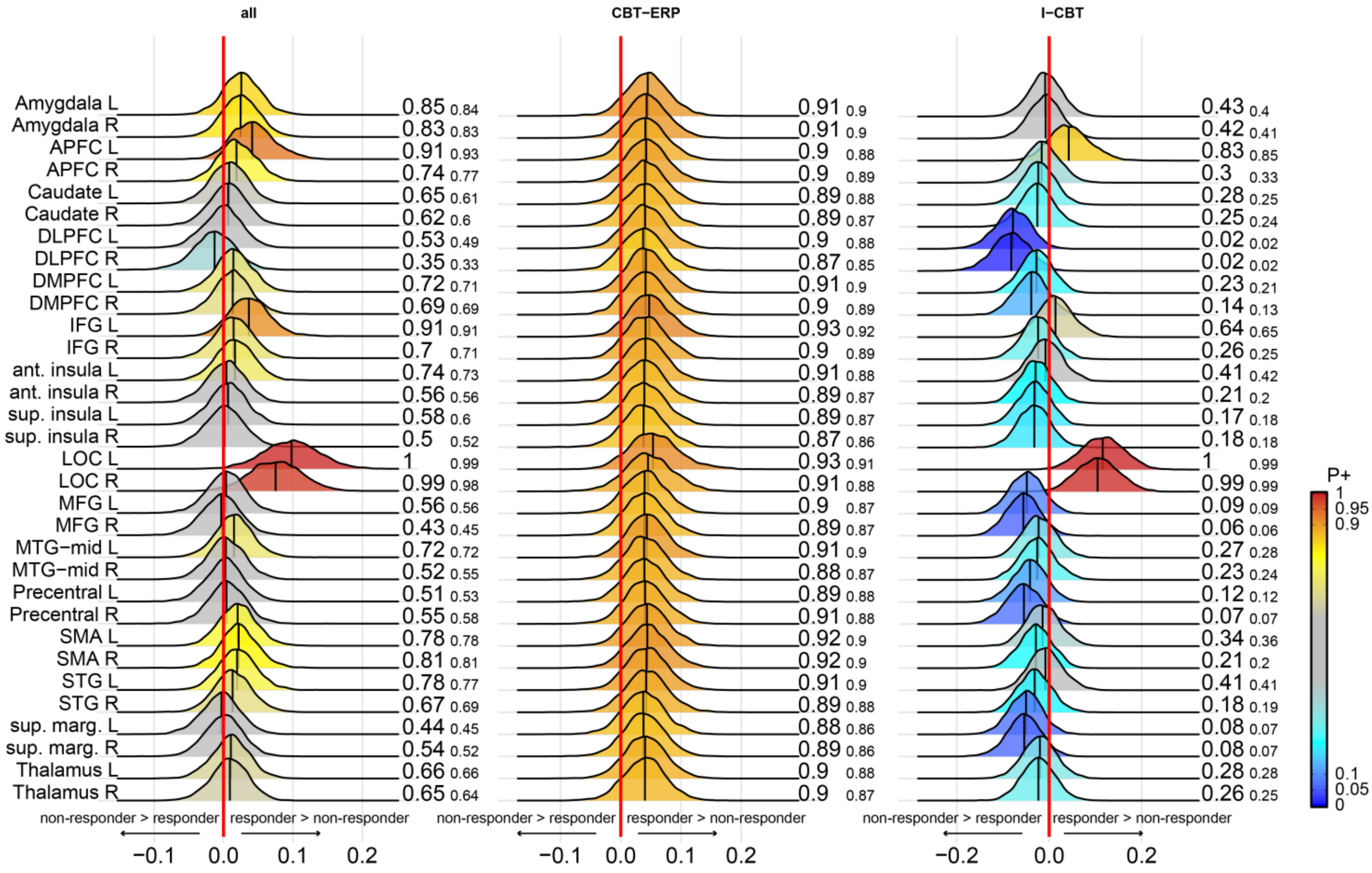

**Supplementary Figure 17 – Bayesian posterior distribution plots of the difference in baseline task activation during the fear contrast between responders and non-responders in the Per Protocol (PP) sample.** The first column shows the difference between responders and non-responders across both treatment groups. Column two and three show the between-group difference in the CBT-ERP and I-CBT treatment conditions, respectively. The posterior distribution communicates the credibility of an effect. Positive posterior probabilities (*P*+) are shown next to each distribution and color coded. *P*+ values ≥0.90 indicate moderate to very high credibility for a positive effect, P+ ≤0.10 indicate moderate to very high credibility for a negative effect. The smaller P+ values represent the posterior probabilities when adjusting for medication status. The meaning of the direction of effects are shown next to the red zero-effect line. See supplementary Table 1 for the definition of the abbreviated regions of interest. All analyses were adjusted for age and sex.

### Association between pre-treatment activation and symptom improvement – PP sample (N=65)

Contrast: OCD

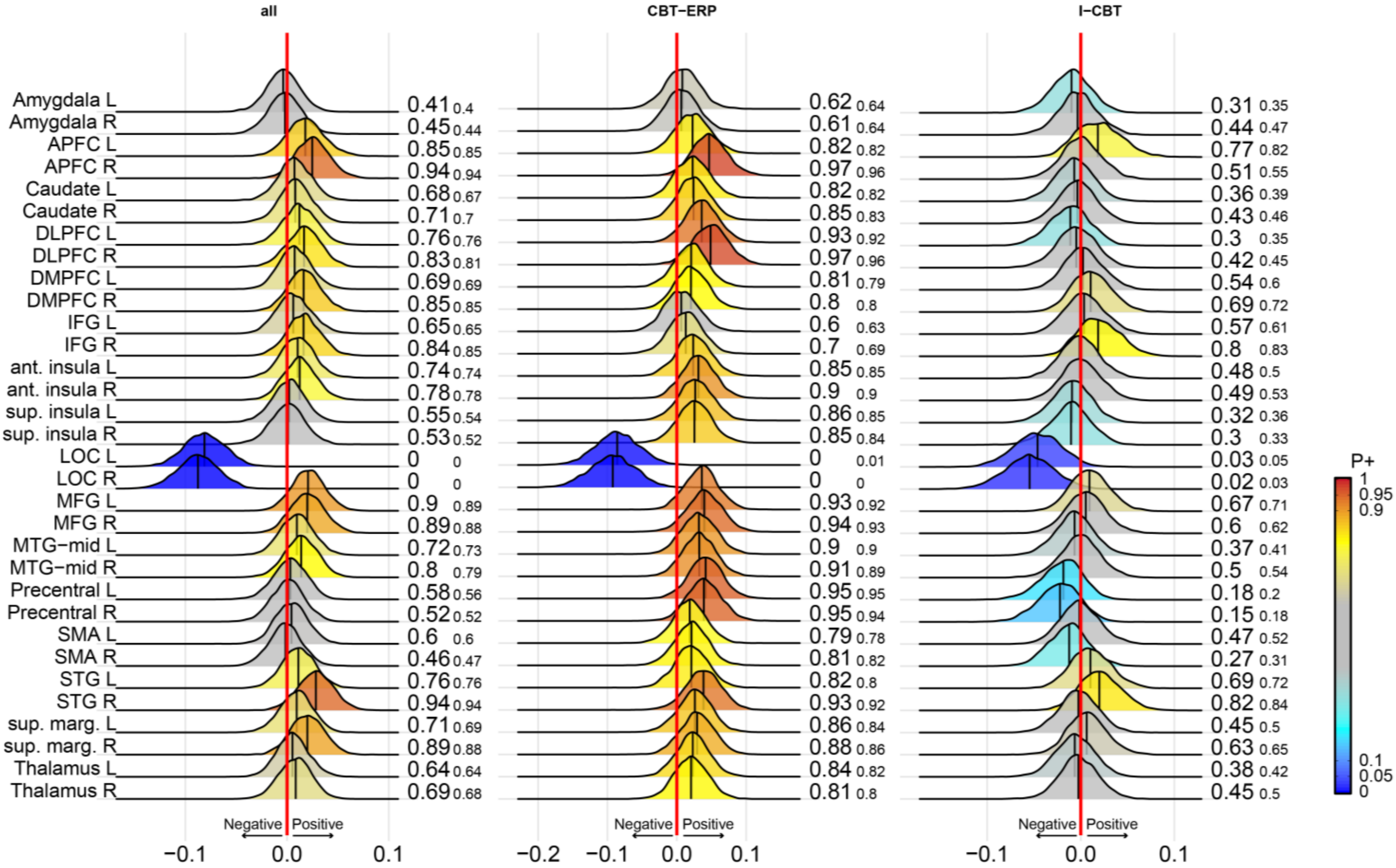

**Supplementary Figure 18 – Bayesian posterior distribution plots of the association between baseline task activation during the OCD contrast and change in OCD symptoms after treatment in the Per Protocol (PP) sample.**

The first column shows the association across both treatment groups. Column two and three show the associations in the CBT-ERP and I-CBT group, respectively. The posterior distribution communicates the credibility of an effect. Positive posterior probabilities (*P*+) are shown next to each distribution and color coded. *P*+ values ≥0.90 indicate moderate to very high credibility for a positive effect, P+ ≤0.10 indicate moderate to very high credibility for a negative effect. The smaller P+ values represent the posterior probabilities when adjusting for medication status. The meaning of the direction of effects are shown next to the red zero-effect line. See supplementary Table 1 for the definition of the abbreviated regions of interest. All analyses were adjusted for age and sex and baseline YBOCS score.

### Difference in pre-treatment activation between responders and non-responders – ITT sample (N=73)

Contrast: OCD

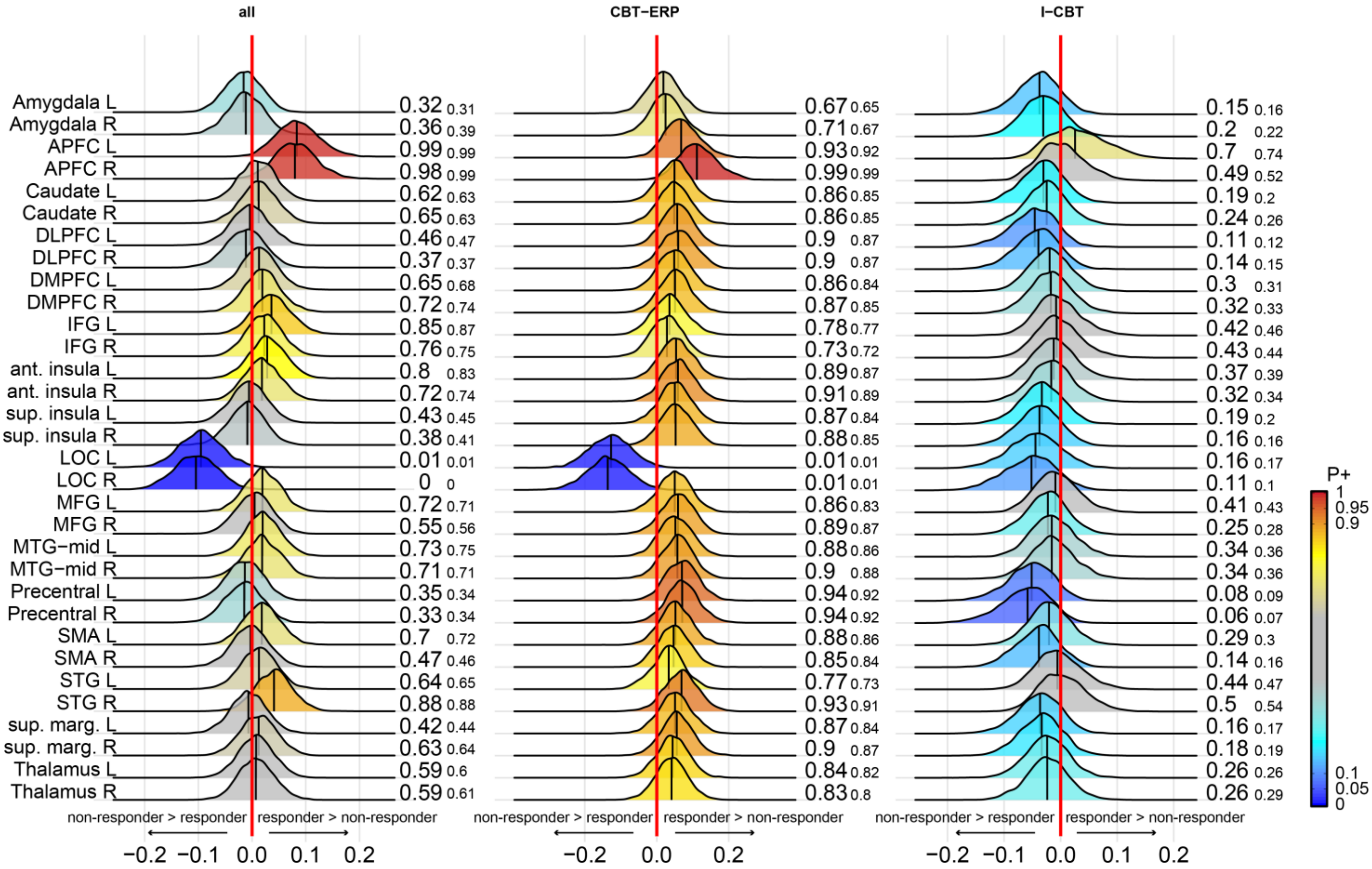

**Supplementary Figure 19 – Bayesian posterior distribution plots of the difference in baseline task activation during the OCD contrast between responders and non-responders in the intention-to-treat (ITT) sample.** The first column shows the difference between responders and non-responders across both treatment groups. Column two and three show the between-group difference in the CBT-ERP and I-CBT treatment conditions, respectively. The posterior distribution communicates the credibility of an effect. Positive posterior probabilities (*P*+) are shown next to each distribution and color coded. *P*+ values ≥0.90 indicate moderate to very high credibility for a positive effect, P+ ≤0.10 indicate moderate to very high credibility for a negative effect. The smaller P+ values represent the posterior probabilities when adjusting for medication status. The meaning of the direction of effects are shown next to the red zero-effect line. See supplementary Table 1 for the definition of the abbreviated regions of interest. All analyses were adjusted for age and sex.

### Difference in pre-treatment activation between responders and non-responders – PP sample (N=65)

Contrast: OCD

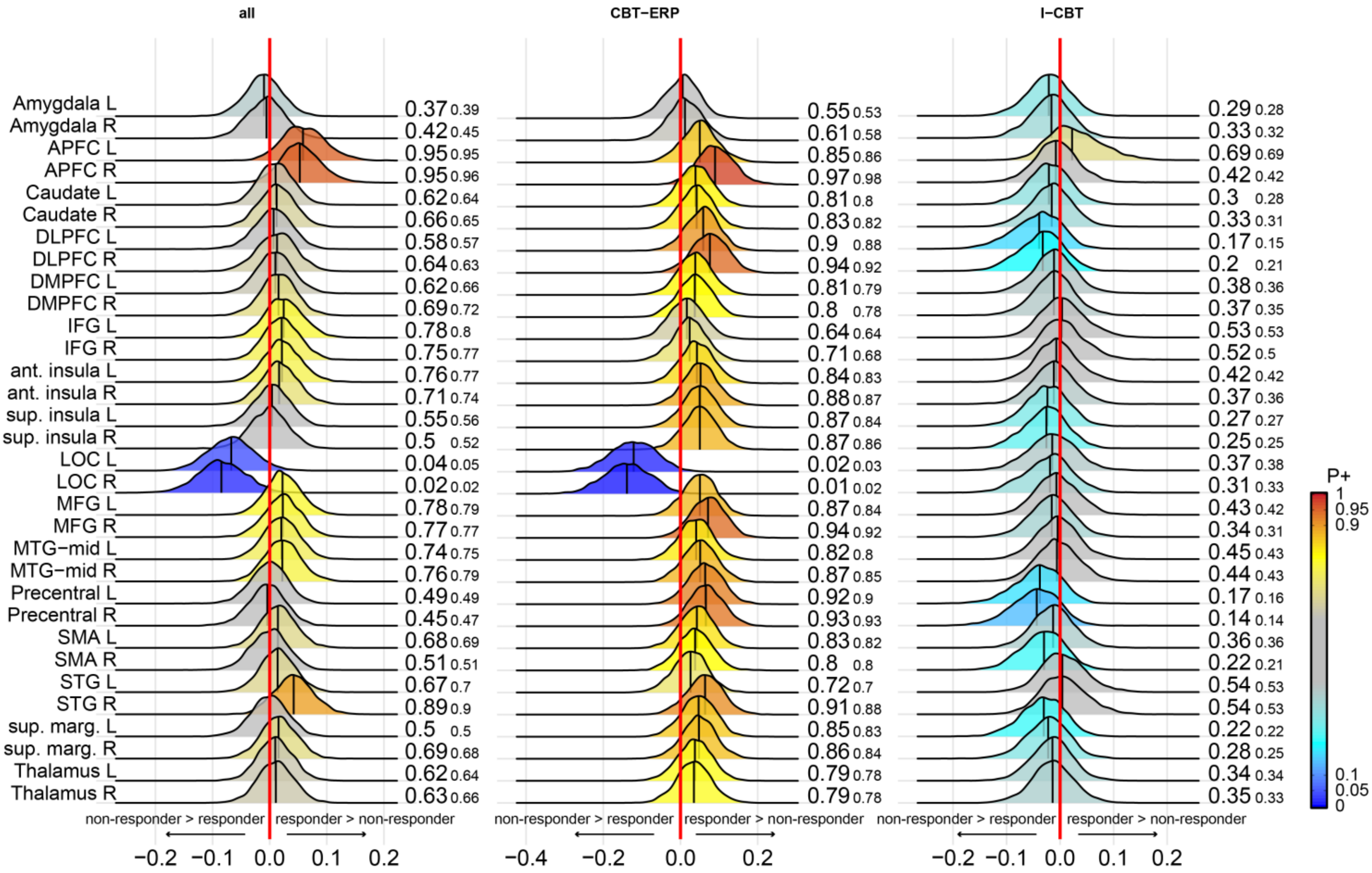

**Supplementary Figure 20 – Bayesian posterior distribution plots of the difference in baseline task activation during the OCD contrast between responders and non-responders in the Per Protocol (PP) sample.**

The first column shows the difference between responders and non-responders across both treatment groups. Column two and three show the between-group difference in the CBT-ERP and I-CBT treatment conditions, respectively. The posterior distribution communicates the credibility of an effect. Positive posterior probabilities (*P+*) are shown next to each distribution and color coded. *P+* values ≥0.90 indicate moderate to very high credibility for a positive effect, P+ ≤0.10 indicate moderate to very high credibility for a negative effect. The smaller P+ values represent the posterior probabilities when adjusting for medication status. The meaning of the direction of effects are shown next to the red zero-effect line. See supplementary Table 1 for the definition of the abbreviated regions of interest. All analyses were adjusted for age and sex.

### Association between pre-treatment activation and symptom improvement – ITT sample (N=73)

Contrast: EMOTION

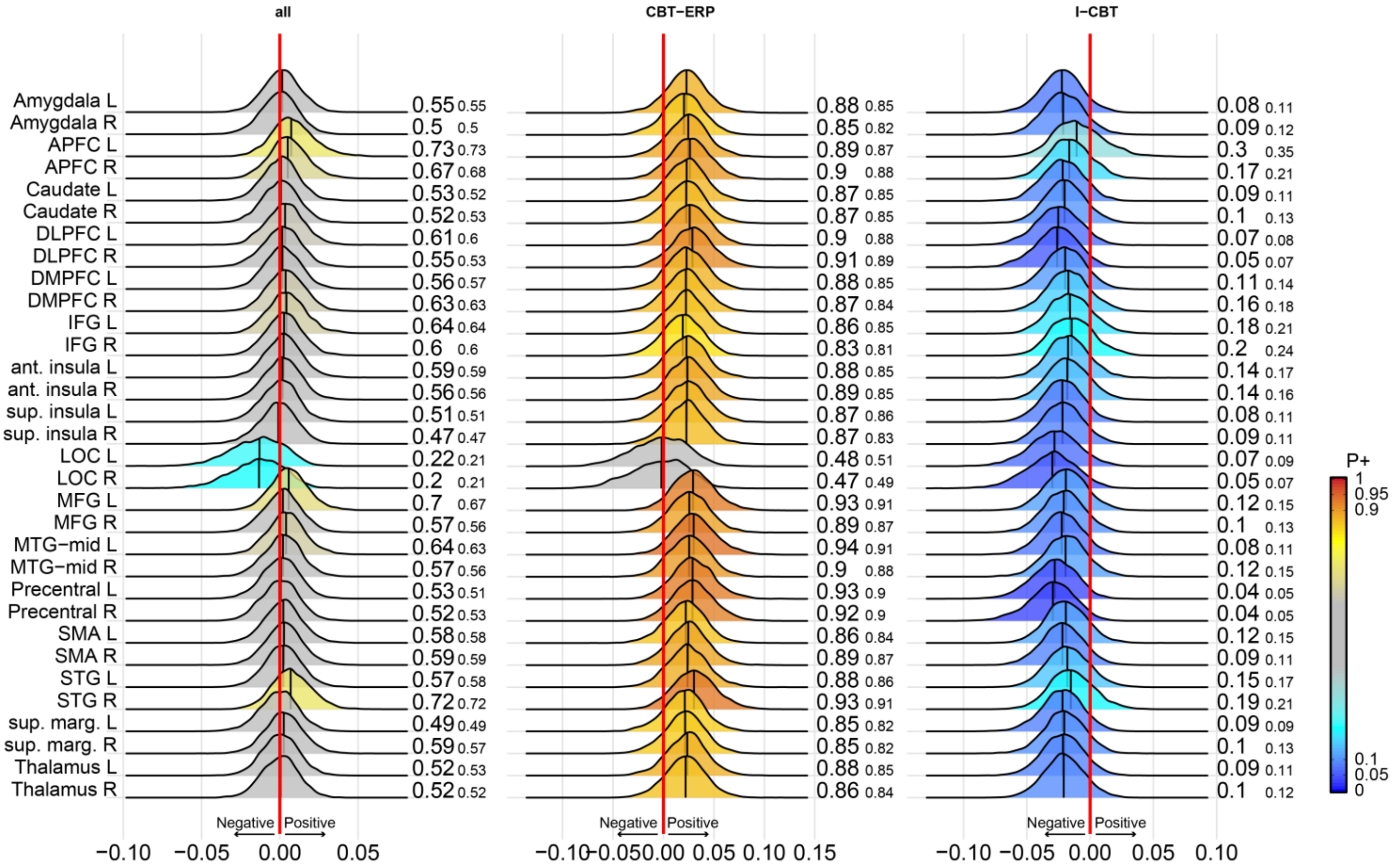

**Supplementary Figure 21 – Bayesian posterior distribution plots of the association between baseline task activation during the emotion contrast and change in OCD symptoms after treatment in the intention-to-treat (ITT) sample.**

The first column shows the association across both treatment groups. Column two and three show the associations in the CBT-ERP and I-CBT group, respectively. The posterior distribution communicates the credibility of an effect. Positive posterior probabilities (*P*+) are shown next to each distribution and color coded. *P*+ values ≥0.90 indicate moderate to very high credibility for a positive effect, P+ ≤0.10 indicate moderate to very high credibility for a negative effect. The smaller P+ values represent the posterior probabilities when adjusting for medication status. The meaning of the direction of effects are shown next to the red zero-effect line. See supplementary Table 1 for the definition of the abbreviated regions of interest. All analyses were adjusted for age and sex and baseline YBOCS score.

### Association between pre-treatment activation and symptom improvement – PP sample (N=65)

Contrast: EMOTION

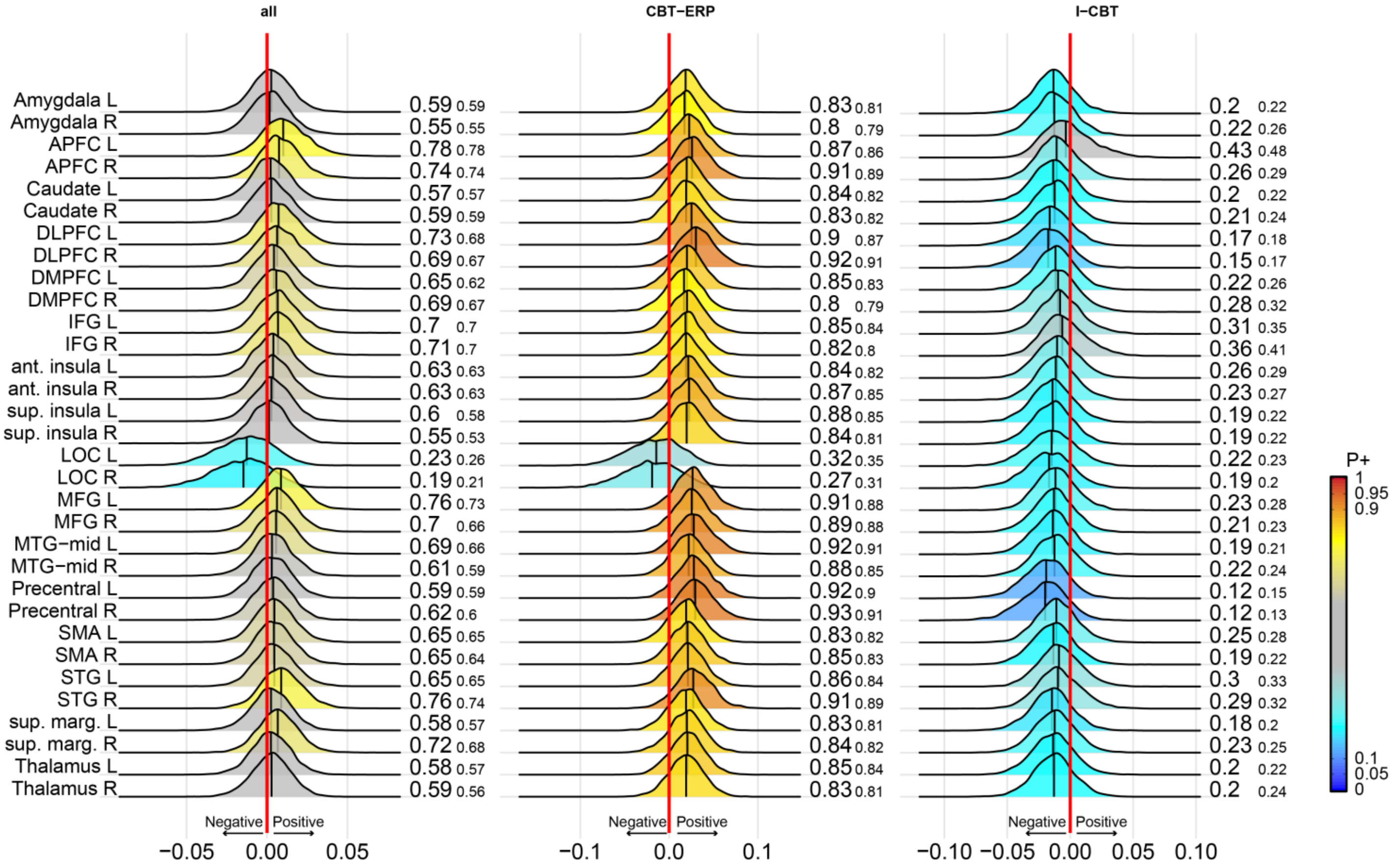

Supplementary Figure 22 – Bayesian posterior distribution plots of the association between baseline task activation during the emotion contrast and change in OCD symptoms after treatment in the Per Protocol (PP) sample.

The first column shows the association across both treatment groups. Column two and three show the associations in the CBT-ERP and I-CBT group, respectively. The posterior distribution communicates the credibility of an effect. Positive posterior probabilities (*P*+) are shown next to each distribution and color coded. *P*+ values ≥0.90 indicate moderate to very high credibility for a positive effect, P+ ≤0.10 indicate moderate to very high credibility for a negative effect. The smaller P+ values represent the posterior probabilities when adjusting for medication status. The meaning of the direction of effects are shown next to the red zero-effect line. See supplementary Table 1 for the definition of the abbreviated regions of interest. All analyses were adjusted for age and sex and baseline YBOCS score.

### Difference in pre-treatment activation between responders and non-responders – ITT sample (N=73)

Contrast: EMOTION

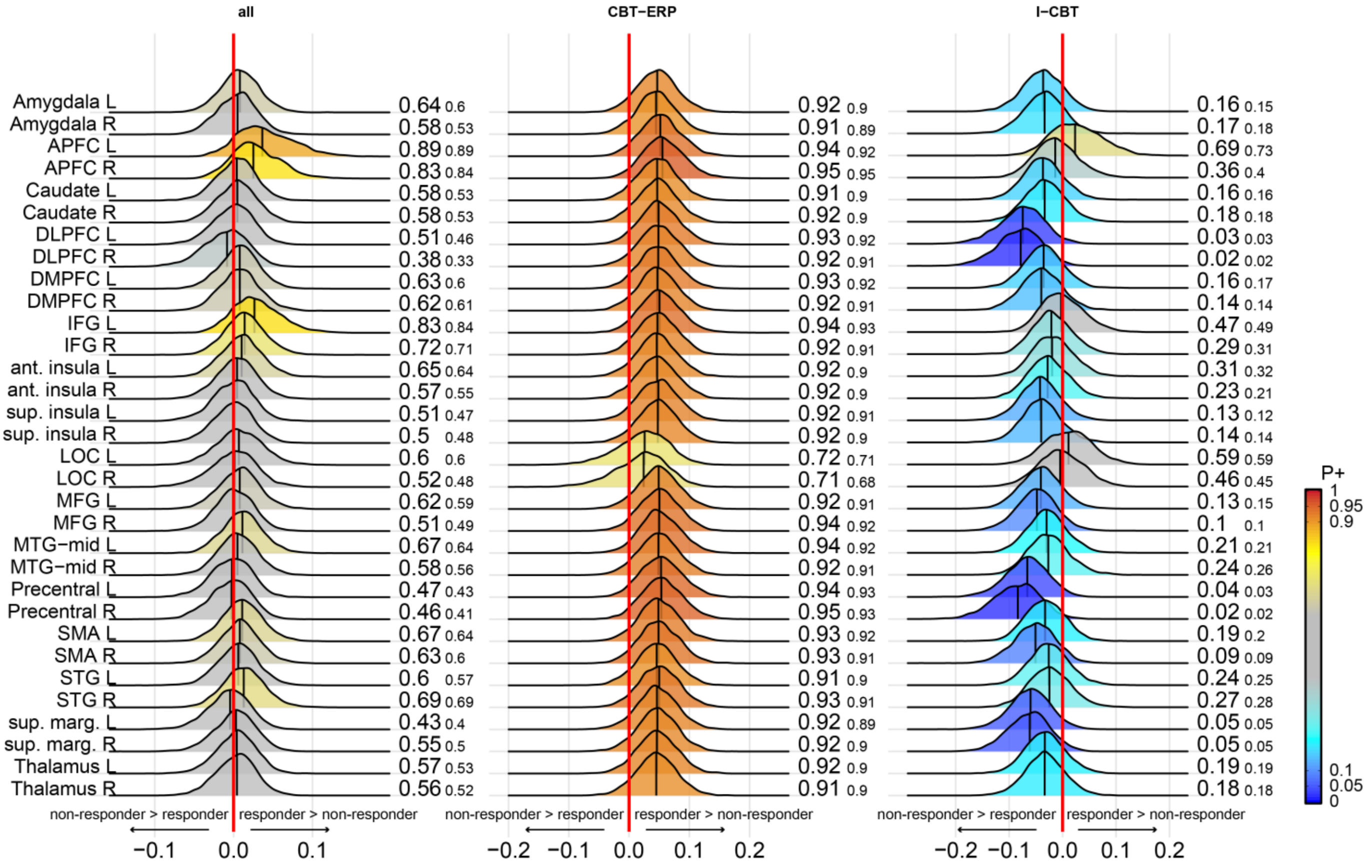

**Supplementary Figure 23 – Bayesian posterior distribution plots of the difference in baseline task activation during the emotion contrast between responders and non-responders in the intention-to-treat (ITT) sample.** The first column shows the difference between responders and non-responders across both treatment groups. Column two and three show the between-group difference in the CBT-ERP and I-CBT treatment conditions, respectively. The posterior distribution communicates the credibility of an effect. Positive posterior probabilities (*P*+) are shown next to each distribution and color coded. *P*+ values ≥0.90 indicate moderate to very high credibility for a positive effect, P+ ≤0.10 indicate moderate to very high credibility for a negative effect. The smaller P+ values represent the posterior probabilities when adjusting for medication status. The meaning of the direction of effects are shown next to the red zero-effect line. See supplementary Table 1 for the definition of the abbreviated regions of interest. All analyses were adjusted for age and sex.

### Difference in pre-treatment activation between responders and non-responders – PP sample (N=65)

Contrast: EMOTION

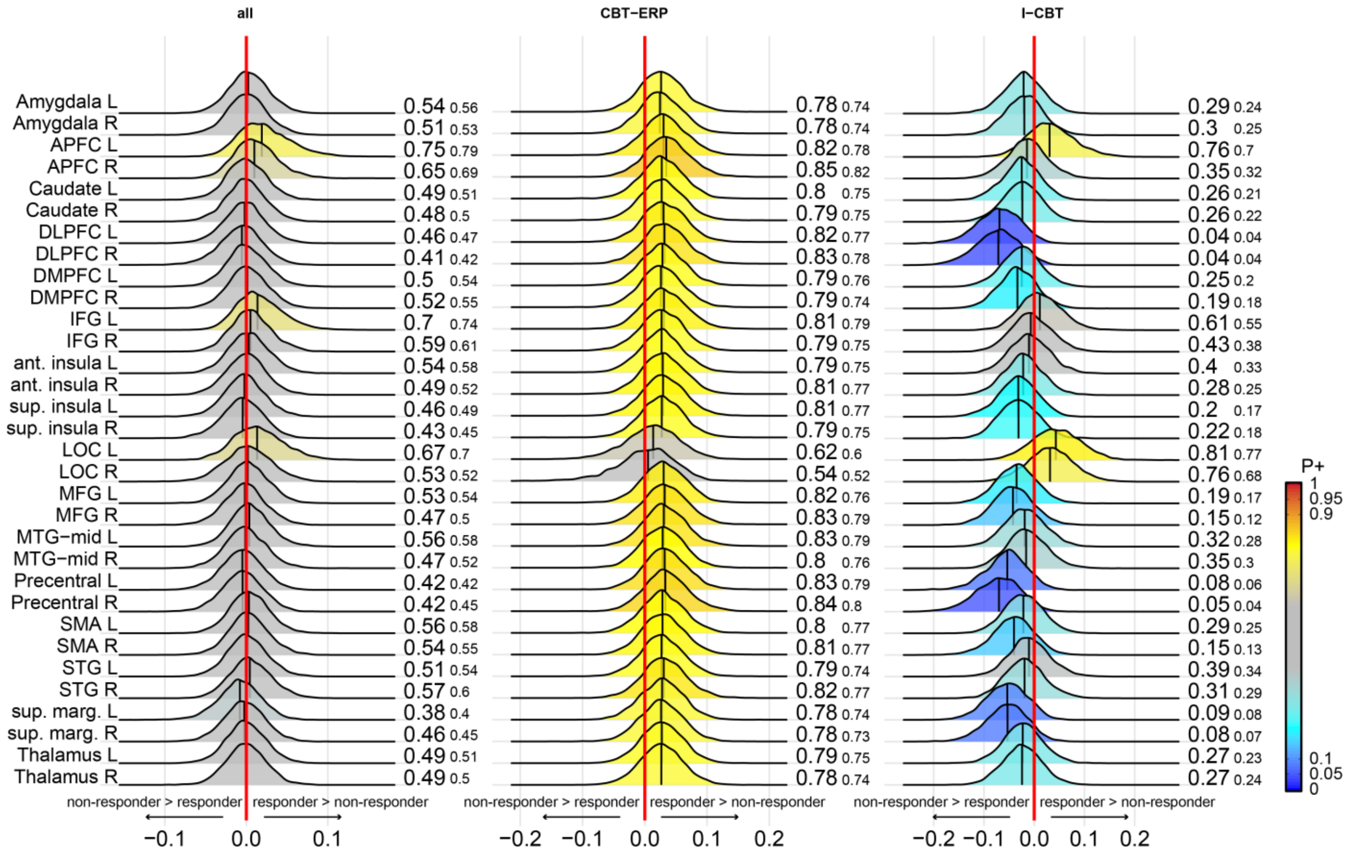

**Supplementary Figure 24 – Bayesian posterior distribution plots of the difference in baseline task activation during the OCD contrast between responders and non-responders in the Per Protocol (PP) sample.**
The first column shows the difference between responders and non-responders across both treatment groups. Column two and three show the between-group difference in the CBT-ERP and I-CBT treatment conditions, respectively. The posterior distribution communicates the credibility of an effect. Positive posterior probabilities (*P*+) are shown next to each distribution and color coded. *P*+ values ≥0.90 indicate moderate to very high credibility for a positive effect, P+ ≤0.10 indicate moderate to very high credibility for a negative effect. The smaller P+ values represent the posterior probabilities when adjusting for medication status. The meaning of the direction of effects are shown next to the red zero-effect line. See supplementary Table 1 for the definition of the abbreviated regions of interest. All analyses were adjusted for age and sex.

#### Whole-brain analyses

**Supplementary Table 13 - Associations between baseline task activation during the fear contrast and change in symptoms after treatment**

| Group, association | Region | Hemisphere | Cluster size (k) | MNI x | y | z | Peak Z |
|---|---|---|---|---|---|---|---|
| **All, positive** | OFC | L | 49 | -38 | 28 | -6 | 3.61 |
| | OFC/anterior insula | L | 35 | -42 | 16 | -16 | 3.41 |
| | Fusiform gyrus | L | 22 | -42 | -60 | -22 | 3.48 |
| | Hippocampus | L | 29 | -26 | -28 | -14 | 3.63 |
| | Cerebellum | R | 86 | 40 | -76 | -30 | 3.48 |
| | Cerebellum | R | 13 | 40 | -66 | -42 | 3.42 |
| | Cerebellum | R | 57 | 8 | -52 | -38 | 3.85 |
| | Cerebellum | R | 53 | 8 | -86 | -34 | 3.66 |
| | Cerebellum | L | 5 | -14 | -88 | -38 | 3.18 |
| | Cerebellum | L | 7 | -18 | -34 | -28 | 3.18 |
| **All, negative** | - | | | | | | |
| **CBT-ERP, positive** | vlPFC | L | 152 | -38 | 52 | -6 | 3.99 |
| | Superior insula | L | 87 | -32 | 6 | 14 | 3.86 |
| | Posterior cingulate cortex | L | 7 | -8 | -48 | 18 | 3.4 |
| | Angular gyrus | L | 38 | -42 | -66 | 46 | 3.37 |
| | Precuneus | L | 6 | -6 | -34 | 30 | 3.57 |
| | Inferior temporal gyrus | L | 10 | -42 | -16 | -30 | 3.43 |
| | Hippocampus | L | 105 | -24 | -28 | -18 | 4.73 |
| | Cerebellum | R | 140 | 30 | -60 | -40 | 3.99 |
| | Cerebellum | R | 87 | 6 | -48 | -40 | 3.72 |
| | Cerebellum | R | 20 | 8 | -86 | -34 | 3.5 |
| **CBT-ERP, negative** | Cuneus | R | 17 | 10 | -92 | 32 | 3.53 |
| **I-CBT, positive** | Cerebellum crus II | R | 6 | 14 | -78 | -42 | 3.54 |
| **I-CBT, negative** | - | | | | | | |

Results are reported at P<0.001, uncorrected for multiple comparisons. Abbreviations: OFC = orbitofrontal cortex, vlPFC = ventrolateral prefrontal cortex.

**Supplementary Table 14 - Associations between baseline task activation during the OCD contrast and change in symptoms after treatment**

| Group, association | Region | Hemisphere | Cluster size (k) | MNI x | y | z | Peak Z |
|---|---|---|---|---|---|---|---|
| **All, positive** | vlPFC | L | 5 | -20 | 62 | 10 | 3.24 |
| | vlPFC | L | 6 | -42 | 44 | -10 | 3.21 |
| | pre-SMA | L | 5 | -28 | 18 | 54 | 3.19 |
| | Angular gyrus | L | 37 | -42 | -62 | 28 | 3.5 |
| | Fusiform gyrus | R | 14 | 42 | -48 | 10 | 3.55 |
| **All, negative** | LOC | R | 23 | 30 | -88 | -12 | 3.58 |
| | LOC | R | 14 | 36 | -74 | -16 | 3.47 |
| | Fusiform gyrus | L | 8 | -52 | -68 | -6 | 3.32 |
| | Parahippocampal gyrus | R | 5 | 30 | -22 | -26 | 3.39 |
| **CBT-ERP, positive** | vlPFC | L | 31 | -38 | 52 | -6 | 3.81 |
| | WM/OFC | R | 8 | 30 | 36 | -4 | 3.65 |
| | WM/OFC | L | 17 | -22 | 50 | 2 | 3.48 |
| | Anterior insula | R | 11 | 28 | 22 | -4 | 3.36 |
| | Anterior insula | L | 8 | -28 | 16 | 2 | 3.29 |
| | Precuneus | L | 24 | -8 | -60 | 42 | 3.68 |
| **CBT-ERP, negative** | Lingual gyrus | L | 22 | -10 | -94 | -10 | 3.42 |
| | LOC | R | 13 | 28 | -86 | -18 | 3.34 |
| **I-CBT, positive** | Angular gyrus | L | 12 | -54 | -68 | 28 | 3.29 |
| **I-CBT, negative** | Amygdala | L | 7 | -32 | 0 | -24 | 3.29 |

Results are reported at P<0.001, uncorrected for multiple comparisons. Abbreviations: OFC = orbitofrontal cortex, vlPFC = ventrolateral prefrontal cortex, pre-SMA = pre-supplementary motor area, LOC = lateral occipital cortex.

### Whole-brain analyses

**Supplementary Table 15 - Associations between baseline task activation during the emotion contrast and change in symptoms after treatment**

| Group, association | Region | Hemisphere | Cluster size (k) | MNI x | y | z | Peak Z |
|---|---|---|---|---|---|---|---|
| **All, positive** | - | | | | | | |
| **All, negative** | - | | | | | | |
| **CBT-ERP, positive** | vlPFC | L | 77 | -38 | 52 | -6 | 4.12 |
| | Precentral gyrus | R | 12 | 22 | -16 | 68 | 3.41 |
| | Precuneus | L | 25 | -8 | -60 | 40 | 3.59 |
| | Hippocampus | L | 26 | -22 | -28 | -18 | 3.73 |
| | Cerebellum | R | 6 | 30 | -60 | -42 | 3.27 |
| | Cerebellum/brainstem | R | 61 | 10 | -50 | -42 | 3.97 |
| **CBT-ERP, negative** | Cuneus | R | 16 | 8 | -92 | 34 | 3.43 |
| **I-CBT, positive** | - | | | | | | |
| **I-CBT, negative** | SMA | L | 47 | -8 | -12 | 62 | 4.24 |
| | Precentral gyrus | L | 7 | -60 | -2 | 38 | 3.39 |
| | Postcentral gyrus | L | 12 | -32 | -38 | 72 | 3.4 |

Results are reported at P<0.001, uncorrected for multiple comparisons. Abbreviations: vlPFC = ventrolateral prefrontal cortex, SMA = supplementary motor area

### Pre and post-treatment group differences in activation

contrast: fear

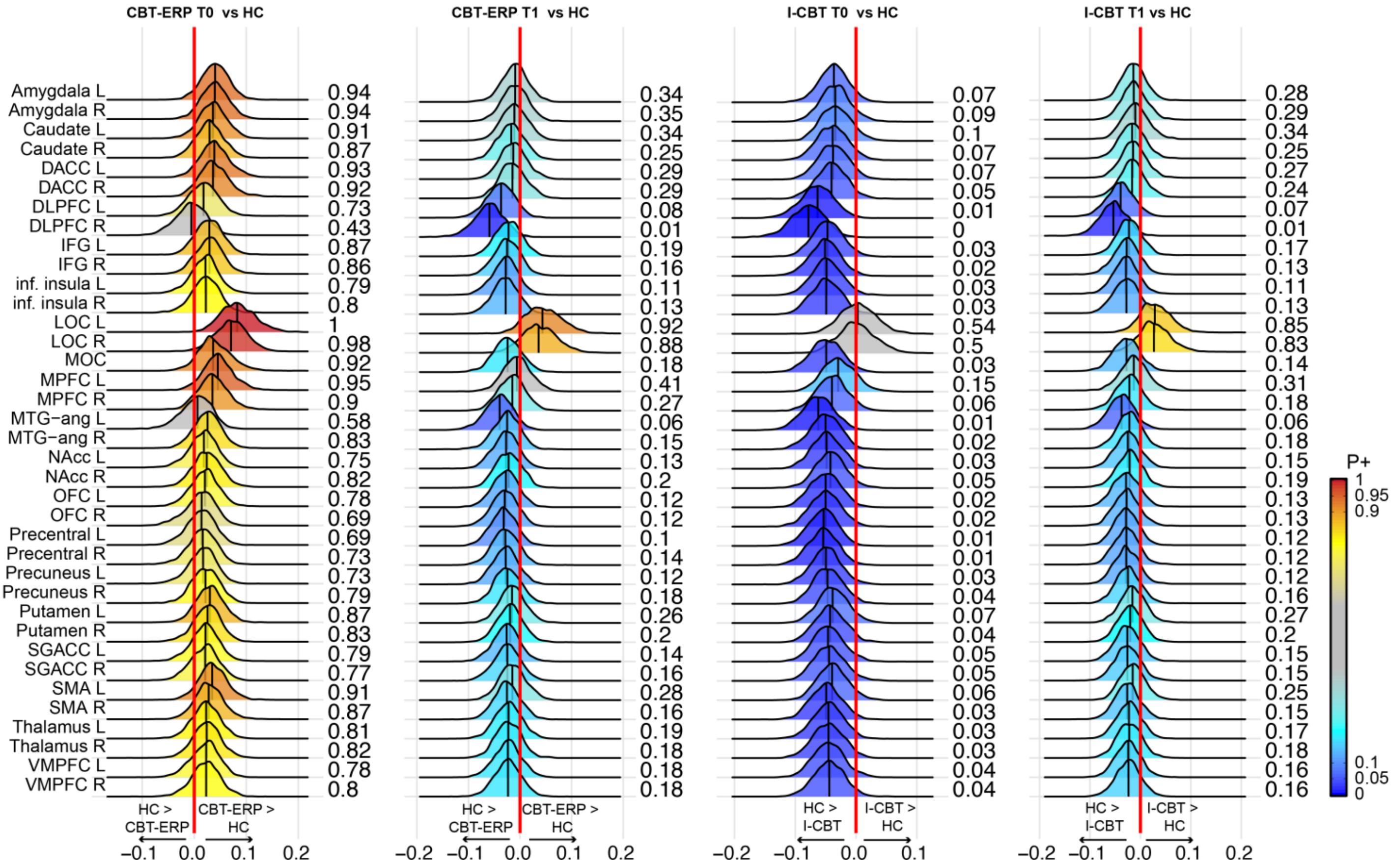

**Supplementary Figure 25 – pre (T0) and post-treatment (T1) differences in activation in the fear contrast in the CBT-ERP and I-CBT treatment groups relative to healthy controls.** The posterior distribution communicates the credibility of an effect. Positive posterior probabilities (*P*+) are shown next to each distribution and color coded. Note that the P+ values for the pre-treatment (T0) comparisons are slightly different than in supplementary figure 4 due to the stochastic nature of the Bayesian analyses. *P*+ values ≥0.90 indicate moderate to very high credibility for a positive effect, P+ ≤0.10 indicate moderate to very high credibility for a negative effect. The meaning of the direction of effects are shown next to the green zero-effect line. See supplementary Table 1 for the definition of the abbreviated regions of interest.

### Pre and post-treatment group differences in activation

contrast: ocd

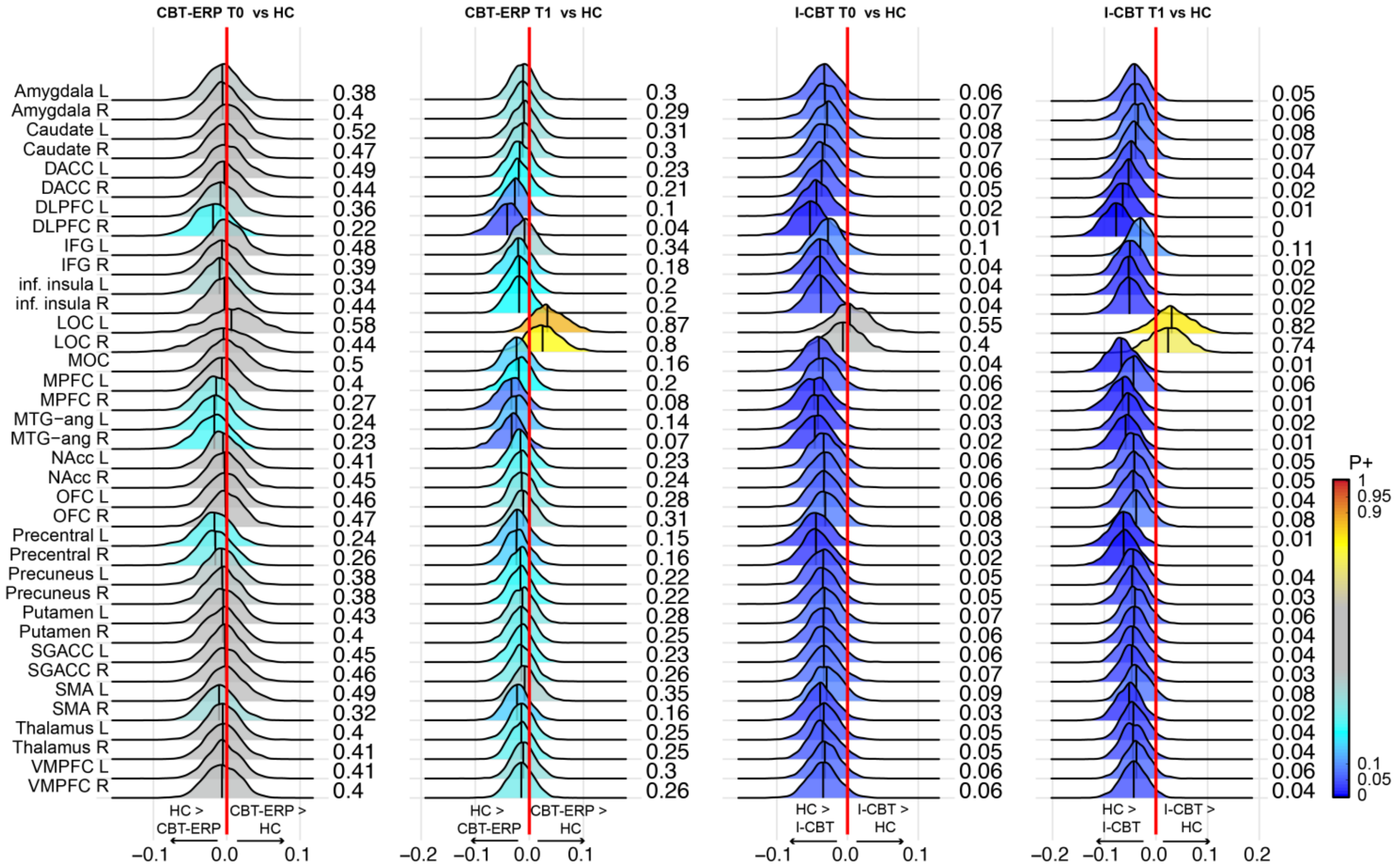

**Supplementary Figure 26 – pre (T0) and post-treatment (T1) differences in activation in the ocd contrast in the CBT-ERP and I-CBT treatment groups relative to healthy controls.** The posterior distribution communicates the credibility of an effect. Positive posterior probabilities (*P*+) are shown next to each distribution and color coded. Note that the P+ values for the pre-treatment (T0) comparisons are slightly different than in supplementary figure 5 due to the stochastic nature of the Bayesian analyses. *P*+ values ≥0.90 indicate moderate to very high credibility for a positive effect, P+ ≤0.10 indicate moderate to very high credibility for a negative effect. The meaning of the direction of effects are shown next to the green zero-effect line. See supplementary Table 1 for the definition of the abbreviated regions of interest.
